# Unifying human infectious disease models and real-time awareness of population- and subpopulation-level intervention effectiveness

**DOI:** 10.1101/2024.01.17.24301344

**Authors:** Rachel L. Seibel, Michael J. Tildesley, Edward M. Hill

## Abstract

During infectious disease outbreaks, humans often base their decision to adhere to an intervention strategy on individual choices and opinions. However, due to data limitations and inference challenges, infectious disease models usually omit these variables. We constructed a compartmental, deterministic Susceptible-Exposed-Infectious-Recovered (SEIR) disease model that includes a behavioural function with parameters influencing intervention uptake. The behavioural function accounted for an initial subpopulation opinion towards an intervention, their outbreak information awareness sensitivity and the extent they are swayed by the real-time intervention effectiveness information. Applying the model to vaccination uptake and three human pathogens - pandemic influenza, SARS-CoV-2 and Ebola virus - we explored through model simulation how these intervention adherence decision parameters and behavioural heterogeneity impacted epidemiological outcomes (cumulative cases and cumulative deaths). From our model simulations we found that in some pathogen systems, different types of outbreak information awareness at different outbreak stages may be more informative to an information-sensitive population and lead to less severe epidemic outcomes. Incorporating behavioural functions that modify infection control intervention adherence into epidemiological models can aid our understanding of adherence dynamics during outbreaks. Ultimately, by parameterising models with what we know about human behaviour towards vaccination adherence, such models can help assist decision makers during outbreaks.

## Introduction

Human behaviour is undoubtedly a driving force of pathogen spread during infectious disease outbreaks. Intervention adherence, government policy, misinformation, travel restrictions and alterations to contact patterns can all impact how the outbreak develops [1–3]. The COVID-19 pandemic provides a recent and notable example of the effects of behavioural heterogeneity. Misinformation played a large role in many countries and amongst online social networks, especially in the early stages of the pandemic and in individual-level decisions to vaccinate [4–6]. In an epidemiologically-ideal scenario lockdowns would have perfect adherence, vaccines would be available on the first day of an emerging disease outbreak and the infections would die out. However, this is not a reflection of reality. Our epidemiological reality is a reflection of the decisions and choices human populations make, which may be based on a plethora of factors such as susceptibility to severe infection, vulnerable relatives or close contacts, moral beliefs, intervention accessibility and cost of infection. Its complex nature often requires models of infectious disease dynamics to make assumptions about behaviour that can lead to appreciable differences between epidemic models and what we observe in reality [7].

Variation between individuals in their behavioural response to infections has been evident for several pathogens of human health concern. For influenza, since many individuals and age-groups are not susceptible to severe infection, behaviours that may be most pressing include lockdown fatigue and vaccination adherence [8–10]. In the context of Ebola outbreak resurgences in the Democratic Republic of the Congo (DRC) in recent years, especially 2018, Vinck *et al*. 2019 identified low trust in government institutions in the region and widespread belief in misinformation regarding Ebola virus [31]. Throughout the COVID-19 pandemic, individuals have expressed their opinions, virtually and in-person, on intervention strategies such as broad scale lockdown, mask mandates and vaccine campaigns [4, 12]. More generally, individuals can be influenced by ideas such as vaccine hesitancy, the beliefs of social contacts or personal vulnerability due to existing health concerns [13–15].

Given that intervention adherence is desirable, efforts towards influencing collective opinions are often a goal of public health officials [4, 16, 17]. However, collective opinions can also be in opposition to the intervention strategy. For instance, human behaviours and thought processes may vary drastically regarding vaccination campaigns compared with social distancing requirements [18]. Understanding how and when these opinions and subsequent behaviours shift is essential in developing impactful public health campaigns and interventions.

There is therefore a multitude of information that is being encountered amongst the population. A prior review by Funk *et al*. categorised the source of information into two types: ‘local’ information and ‘global’ information [19]. Local information corresponds to information originating from an individual’s social neighbourhood (subpopulation). Global information corresponds to publicly available information. We note the information awareness process for a disease outbreak is independent from the spatial awareness process for a disease outbreak (‘local spatial awareness’ can refer to awareness of disease spread in your neighbourhood, whereas ‘global spatial awareness’ can refer to awareness of disease spread over a broad spatial extent - e.g. regional, national and/or international). We stress that in this study when referring to ‘local’ or ‘global’ information, we are specifically referring to the information awareness process (not the spatial awareness process).

A valuable methodological development in infectious disease modelling would be reliably capturing real-time changes in opinions towards disease control strategies as well as real-time disease prevalence. It is anticipated these integrated epidemiological-behavioural dynamics can help enhance the robustness of modelling findings provided to decision makers in the public health sector [20–22]. Nonetheless, due to data limitations and inference challenges, behavioural dynamics are often omitted or, when key aspects are included, they are usually simplified [19, 23–26]. One such example is intervention adherence, such as vaccine uptake rate for a vaccination programme. Modellers may make the assumption of a fixed vaccine uptake rate, or allow for uptake rates to depend on health episode measures such as cases, hospitalisation or deaths. However, such assumptions omit other variables that may impact an individual’s decision to get vaccinated; their initial opinion on the intervention strategy, the cost to the individual to adopt the intervention and their awareness of the intervention’s effectiveness of disease control within their social contacts as well as the greater population [27]. As these processes directly impact decisions to adhere to public health intervention strategies, there is a need to develop mathematical models of infectious disease dynamics that explicitly incorporate such mechanisms.

In this study, we highlight three key human behaviours relevant to decisions to adhere to intervention strategies: (i) their initial preference towards the intervention (and perceived risk of infection); (ii) their tendency to react to information about the outbreak; and (iii) the extent they are swayed by real-time intervention effectiveness information (from both ‘local’ and ‘global’ information perspectives), where poor health outcomes in vaccinated individuals could have detrimental impacts on the rate of vaccine uptake amongst the population [28, 29]. We constructed a compartmental, deterministic SEIR-type disease model that explicitly featured the three aforementioned intervention adherence decision making considerations at the subpopulation level, grouping individuals by vaccination status and behavioural traits. Applied to vaccination uptake and three human pathogens - pandemic influenza, SARS-CoV-2 and Ebola virus - we explored through model simulation how these intervention adherence decision parameters and behavioural heterogeneity in the population impacted epidemiological outcomes (cumulative cases and cumulative deaths). Our simulation-based study revealed how the data stream informing the real-time perception of vaccine effectiveness (either cases- or deaths-based) that would result in lower public health burdens can differ between pathogens. Furthermore, there was notable sensitivity in outbreak size under different assumptions regarding the population split in behavioural traits. It is therefore important that consideration is given to behavioural heterogeneity to intervention adherence in the population, and the explicit factors that influence intervention adherence, to enable improved insights into potential epidemic impacts in future infectious disease outbreaks.

## Methods

Our methodological approach involved developing a deterministic model of infectious disease dynamics that compartmentalised the population (into subpopulations) by vaccination status and behavioural traits. We chose a deterministic modelling approach to allow us to explore a large parameter space with overall shorter computational times compared with a stochastic modelling framework, whilst still capturing epidemiological impacts of underlying subpopulation-level behavioural traits. We investigated the implications on infection dynamics of varying levels of vaccine opinion and information sensitivity, for ‘local’ (subpopulation-level) and ‘global’ (population-level/public) information awareness about vaccine effectiveness; this construction for the behavioural modifier captures the behavioural dynamic of a reduction in vaccine uptake caused by breakthrough infections and deaths in vaccinated individuals [30]. As a sensitivity analysis, we applied our model to three pathogens of public health concern: pandemic influenza, SARS-CoV-2 and Ebola. We selected these three pathogens as vaccines have been developed for each one, whilst they also exhibit distinctive epidemiological traits with regards to spreading potential in an immunologically naïve population (i.e. basic reproduction number) and infection fatality rate. It should be noted that we refer to pandemic influenza rather than seasonal influenza given our assumption of a vaccine-naïve population.

By exploring underlying disease parameters representative of three different human pathogens, we sought to capture the variability in epidemiological severity in different pathogen systems and behavioural structures. We felt this to be particularly pertinent as studies that have previously explored human behaviour during outbreaks often focus on a single pathogen system [4, 8, 31]. Our methodology comprised multiple aspects that we detail in turn: (i) the base mathematical model of infectious disease transmission and pathogen disease history; (ii) definition of our behavioural functions that mechanistically modulated the vaccine uptake rate; (iii) pathogen-specific model parameterisation; (iv) computational simulations to numerically evaluate the scenarios of interest.

### Mathematical model of pathogen disease history and transmission dynamics

#### Disease history

To simulate the infection dynamics and encapsulate the disease history of the three selected human pathogens (pandemic influenza, SARS-CoV-2, Ebola), our model foundation was a deterministic, compartmental Susceptible-Exposed-Infectious-Recovered (SEIR) model. Infected individuals had a latent period, with a duration of *σ*^−1^ days, followed by an infectious period, with a duration of *γ*^−1^ days. Infectious individuals could then die or recover, which in the absence of interventions were allocated by proportions *m* and (1−*m*) respectively. Each of these parameters were pathogen-specific. For the purposes of this study, we did not include demographic processes (births and natural deaths) in our model as the timescales of the simulated outbreaks were short (less than a decade). With the behavioural complexities being the focus of our study, to simplify the model we also chose not to include age-stratification. That being said, we acknowledge the inclusion of demographic processes and age-stratification as viable extensions of the model. We give further remarks on the potential implications of the inclusion of these processes and attributes in the Discussion.

#### Vaccination and behaviour stratification

We further stratified the population by two additional attributes. The first attribute was vaccination status, with *u* and *v* subscripts denoting unvaccinated and vaccinated classes, respectively. The second attribute was behavioural grouping, with *i* subscripts indicating a subgroup with unique behaviour-associated attributes—in our simulations, the subgroups represented vaccine-resistant, vaccine-hesitant and vaccine-accepting subpopulations. As a modelling simplification, we assumed no movement between behavioural subpopulations. We also assumed that no other control measures were used except for vaccination. These model assumptions enabled us to focus on the epidemiological impacts of vaccine beliefs and sensitivity to outbreak information within subgroups. We acknowledge these assumptions could be relaxed, with further remarks given in the Discussion.

#### Implementation of vaccination

Considering collectively disease status, vaccination status and behavioural group, within our model we defined the following unvaccinated compartments (visualised in (Fig. 1)): susceptible (*S*_*u,i*_), exposed (*E*_*u,i*_), infectious (*I*_*u,i*_), recovered (*R*_*u,i*_), pre-death (*PD*_*u,i*_) or deceased (*D*_*u,i*_). The vaccinated compartments were similarly defined: susceptible (*S*_*v,i*_), exposed (*E*_*v,i*_), infectious (*I*_*v,i*_), recovered (*R*_*v,i*_), pre-death (*PD*_*v,i*_) or deceased (*D*_*v,i*_). Only unvaccinated individuals in the susceptible, exposed, and recovered classes could move to the respective vaccinated classes. We made a decision to exclude infectious individuals from vaccination to reflect a scenario often included in disease models where infectious individuals are symptomatic and may either be in self-isolation or encouraged by public health officials to wait to vaccinate until they have recovered. Although, we should note that excluding infectious individuals from vaccination did not have an impact on the daily vaccination rates. Also note that the compartments corresponded to the absolute numbers in each disease and vaccination state.

**Figure 1.**
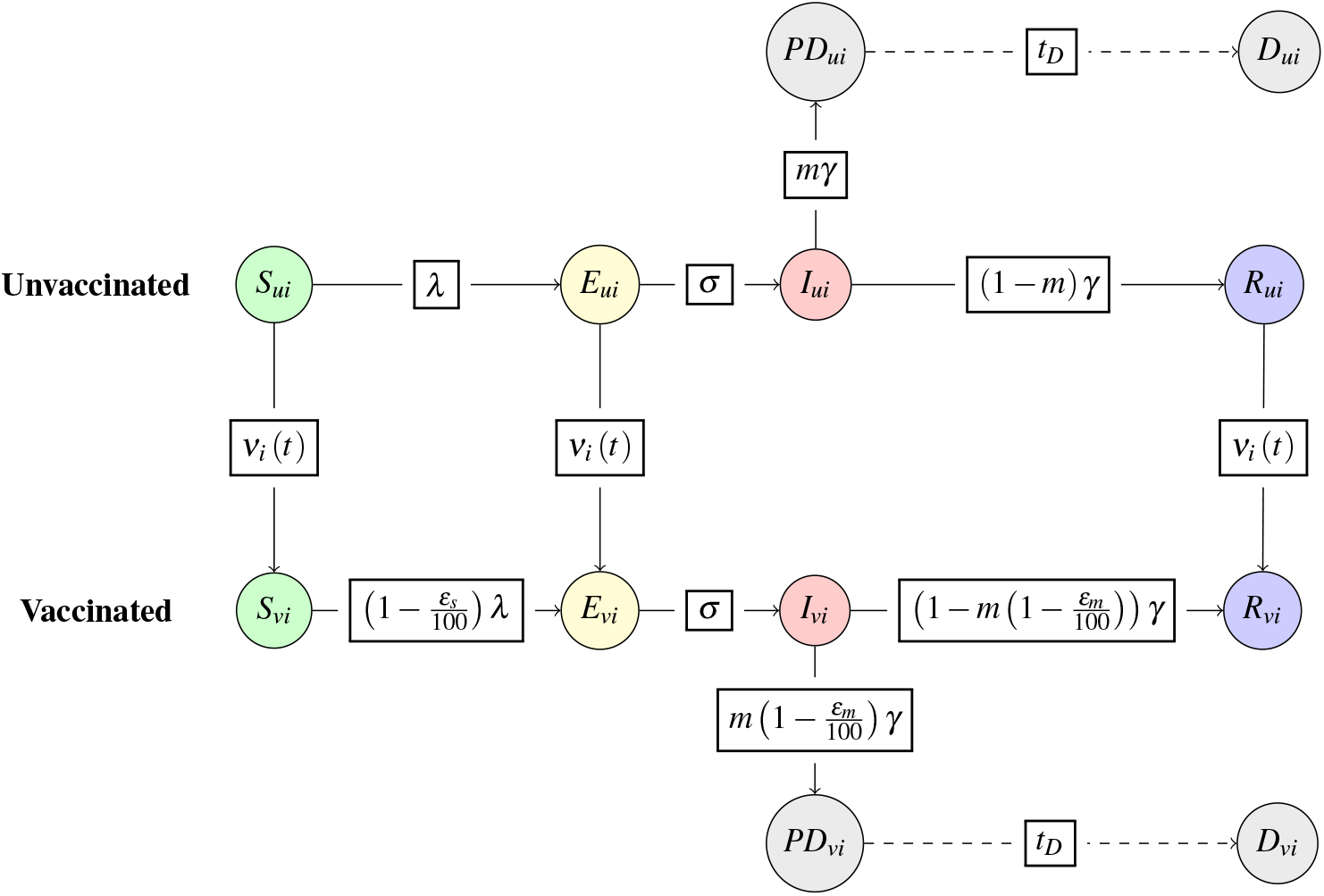
Representation of the underlying disease model with vaccination and time- and behaviour-dependent vaccine uptake, *ν* _*i*_ (*t*). Solid arrows show the flow of individuals between compartments. Dashed arrows indicates the time lags from hospitalisation to reported death, *t*_*D*_. Boxes contain the transition rates between compartments. The subscript *i* denotes the behavioural group, which in our case is determined by vaccine opinion *ρ*. The subscript *u* denotes unvaccinated individuals and the subscript *v* denotes vaccinated individuals. *λ* is the force of infection, *σ* is the rate of progression from the exposed state to the infectious state, *γ* is the rate of recovery from the infectious state and *m* is the infection fatality probability. *ε*_*s*_ represents the vaccine efficacy for infection blocking and *ε*_*m*_ represents the vaccine efficacy for reducing severe disease outcomes. *ν*_*i*_ (*t*) represents the daily vaccine uptake rate. More information about the model equations and parameter estimates can be found in Equation (1) and Table 1.

For intervention-adherent individuals who received the vaccination, we assumed a dual ‘leaky’ vaccination action of (i) being infection blocking and, (ii) reducing severe outcomes conditional on being infected. For the purposes of this study, we also made simplifying assumptions that the vaccine did not reduce disease transmissibility and that the vaccine had the same percentage effectiveness for each action, *ε* ∈ [0, 100] and that the effects were gained instantaneously once vaccinated (i.e. there was no delay in the relevant level of protection being induced post the vaccine being administered). Although we do not explore scenarios in which the vaccine efficacies of infection blocking and reduction of severe disease have different values to one another, it is important to separate these actions in the case that real-world estimates are used to parameterise this model. Such parameterisation is plausible, with SARS-CoV-2 being one example pathogen where there has been reporting of vaccine effectiveness estimates for infection blocking and preventing severe outcomes (e.g. hospitalisations) [32]. Similarly, influenza vaccination has been found to reduce disease severity even without an appreciable infection blocking action [33]. On each new day of the outbreak, the daily vaccine uptake rate, *ν*_*i*_ (*t*), was updated according to the implementation of the behavioural function. We ceased vaccination when the number of individuals eligible for vaccination was close to zero (below 10) for the purposes of reducing the time duration of simulations. See Table 1 for a summary of parameter notation.

**Table 1.** Disease parameter estimates for pandemic influenza, SARS-CoV-2 and Ebola. Parameter values without references were assumptions made for the purposes of this study. We derived *β* from estimates of *R*_0_ and *γ*. Units associated with the parameters are given in parentheses in the parameter description – parameter with no units specified are dimensionless.

| Parameter | Description | Estimate |  |  |
| --- | --- | --- | --- | --- |
|  |  | Pandemic influenza | SARS-CoV-2 | Ebola |
| $R_0$ | Basic reproduction number | 1.5 [34] | 3 [35] | 2 [36] |
| $\beta$ | Transmission rate (day<sup>-1</sup>) | 0.3 | 0.43 | 0.29 |
| $\sigma$ | Rate of progression to infectious disease (day<sup>-1</sup>) | 0.5 [37] | 0.2 [38] | 0.5 [36] |
| $\gamma$ | Rate of recovery or death (day<sup>-1</sup>) | 0.2 [37] | 0.14 [38] | 0.14 [36] |
| $m$ | Probability of unvaccinated death, instead of recovery, due to infection | 1x10<sup>-4</sup> [39] | 6.38x10<sup>-3</sup> [40] | 3.9x10<sup>-1</sup> [41] |

**Table 2.** Behavioural function *ν* _*i*_ (*t*) parameter descriptions. Further information about the values used for each parameter can be found in Table 3 and the formulations of *θ* (*t*) can be found in Equations (6) and (7).

|  | Parameter description | Range |
| --- | --- | --- |
| $v(t)$ | Daily vaccine uptake rate | [0,0.02] |
| $v_0$ | Baseline vaccine uptake rate | 0.005 |
| $\rho$ | Vaccine opinion | [0,2] |
| $\alpha$ | Information sensitivity | [0,2] |
| $\theta(t)$ | Outbreak information | [0,1] |
| $\mu$ | Memory window (days) | (1, Full history] |

#### Mathematical model equations

Under these modelling assumptions, the dynamics were governed by a system of ordinary differential equations (ODEs):

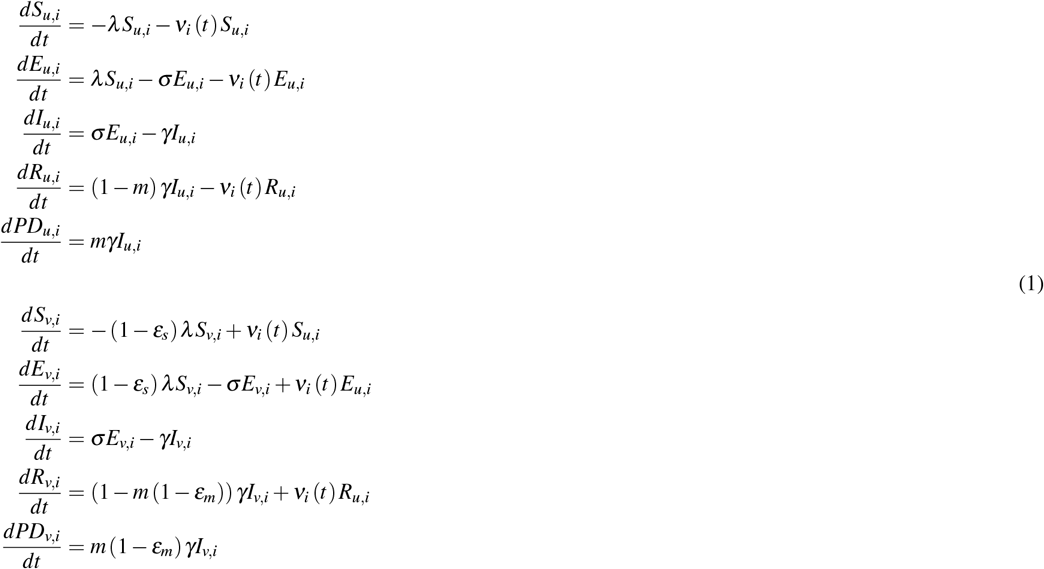

The force of infection, *λ*, was defined as:

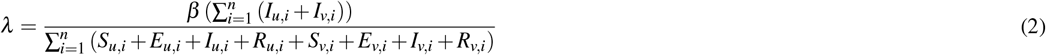

where *β* was the transmission rate for the given pathogen. Here we assumed vaccination status did not impact the rate of transmission. The denominator corresponded to the number of individuals who were alive and not hospitalised at time *t* in the simulated outbreak.

From the time point of entering the pre-death compartments, *PD*_*u,i*_ and *PD*_*v,i*_, there was a time lag, *t*_*D*_, of seven days until the individuals were considered deceased (*D*_*u,i*_, *D*_*v,i*_). This parameterisation was chosen as it reflected a plausible duration from severe disease to reporting of deaths for all three pathogens being studied given values from the scientific literature [42–44]. The total number of unvaccinated individuals in each behavioural subpopulation was *N*_*u,i*_ = *S*_*u,i*_ + *E*_*u,i*_ + *I*_*u,i*_ + *R*_*u,i*_ + *PD*_*u,i*_ + *D*_*u,i*_ and the total number of vaccinated individuals in each behavioural subpopulation was *N*_*v,i*_ = *S*_*v,i*_ + *E*_*v,i*_ + *I*_*v,i*_ + *R*_*v,i*_ + *PD*_*v,i*_ + *D*_*v,i*_. The rate of vaccine uptake, *ν*_*i*_ (*t*), depended on the behavioural function, with further details given in the following subsection.

### Implementation of the behavioural function

In our model, we considered human behaviour related to vaccination by scaling a baseline daily vaccine uptake rate. We assumed a baseline daily vaccine uptake, *ν*_0_, of 0.005 individuals per day, which was modified according to four factors (each detailed below): vaccine opinion, information sharing, outbreak information awareness and memory window.

#### Vaccine opinion and information sharing

We outline here two parameters that correspond to key aspects of human behaviour that can introduce heterogeneity into vaccine uptake. The first was vaccine opinion, *ρ*, corresponding to the initial opinion individuals had prior to the onset of the outbreak. The second was information sensitivity, *α*, which accounted for the sensitivity individuals had to information throughout the simulated outbreak.

We examined two population types in our study: homogeneous and heterogeneous. For scenarios where the population was homogeneous, everyone in the population has the same values for vaccine opinion, *ρ*, and information sensitivity, *α*. For scenarios where the population was heterogeneous, the heterogeneity was with respect to vaccine opinion, *ρ*. The population was split into three subpopulations with different levels of *ρ* to represent vaccine-resistant (*ρ*_*i*_ = 0), vaccine-hesitant (*ρ*_*i*_ = 1) and vaccine-accepting groups (*ρ*_*i*_ = 2). We assumed that everyone had the same information sensitivity *α*, however, this variable could be explored in further simulation studies.

For the purposes of our study, we examined vaccine opinions (*ρ*) and information sensitivities (*α*) ranging from 0 to 2. We chose a fine resolution of these parameters in homogeneous scenario 1 and later selected a representative subset to explore in the remaining scenarios.

#### Outbreak information and memory window

As we were interested in the potentially negative behavioural response on vaccination uptake to vaccinated individuals suffering infection and serious health episodes, recall that our selected dependency for vaccine behaviour modification was a measure of real-time vaccine effectiveness. We had four forms for this outbreak information awareness dependency, whose computation would require data on new cases or new deaths for both unvaccinated and vaccinated individuals (Eqs. (4) and (5)).

These expressions incorporated a population-level memory window, *µ*, which we defined as the amount of time (days) prior to time *t* from which outbreak information, *θ*, was computed. On each new day of a simulation, the daily vaccine uptake rate, *ν*_*i*_ (*t*), was updated by considering the outbreak information, *θ* (*t*), of interest. In our case, we explored the number of new cases or new deaths within the memory window, time *t*− 1 −*µ* to time *t*− 1. For simplicity, we refer to time *t* −1− *µ* as *t*_*µ*_. In our model we included a time lag, *t*_*D*_, of seven days until hospitalised individuals were considered deceased. This time delay is a key difference between the two types of outbreak information we explored, with deaths being reported at least one week later than cases.

We then calculated the outbreak information for the respective unvaccinated and vaccinated subpopulations. The equations for calculating new cases for each unvaccinated and vaccinated subpopulation, *i*, were given, first, by a set of ODEs for tracking vaccination status at the time of infection:

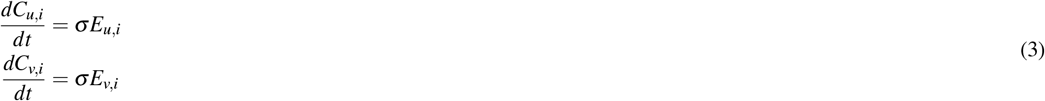

then, by the following equations to determine new cases by vaccination status:

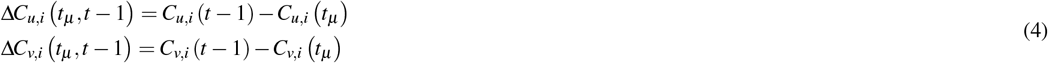

The equations for calculating new deaths for each unvaccinated and vaccinated subpopulation, *i*, were given by:

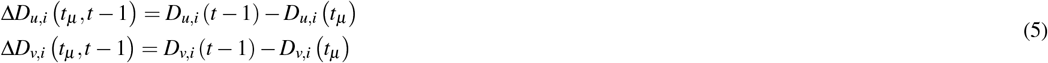

We defined our local outbreak information awareness equations, *θ*_*LC,i*_ and *θ*_*LD,i*_, to correspond to local (subpopulation-level) cases or deaths in unvaccinated individuals relative to cases or deaths in all individuals in the subpopulation:

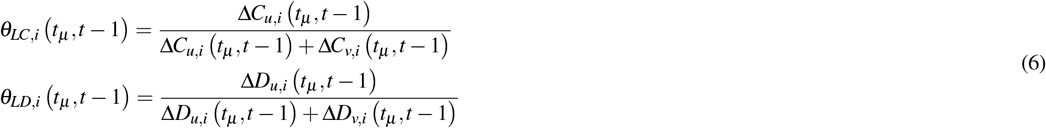

Similarly, we defined our global outbreak information awareness equations, *θ*_*GC*_ and *θ*_*GD*_, to correspond to global (population-level) cases or deaths in unvaccinated individuals relative to cases or deaths in all individuals in the population:

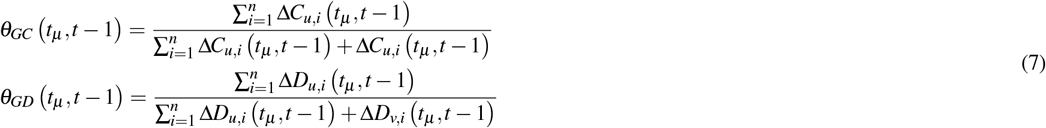

#### Modification of vaccine uptake rate due to behavioural attributes

We modified the daily vaccine uptake rate, *ν*_*i*_ (*t*), by the behavioural function, which accounted for a baseline vaccine uptake rate, *ν*_0_, vaccine opinion, *ρ*_*i*_, information sensitivity, *α*, outbreak information awareness, *θ*_*i*_ *t*_*µ*_, *t* − 1 and memory window duration, *µ*:

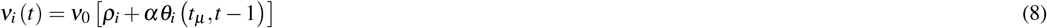

The equation above represents the behavioural function *ν*_*i*_ (*t*) for a behaviourally-heterogeneous population (Eq. (8)). It should be noted that information sensitivity *α* is not notated with a subscript *i* — although we do not vary information sensitivity within simulations for the purposes of this study, *α* could be varied in further studies. We also note that *θ*_*i*_ in the equation can be replaced by *θ*_*LC,i*_, *θ*_*LD,i*_, *θ*_*GC*_ or *θ*_*GD*_.

By design, the outbreak information equations described in the preceding ‘Outbreak information and memory window’ section result in a decrease in the daily vaccine uptake rate for non-fully effective vaccines. This embodies a situation where, due to vaccine effectiveness being less than 100%, breakthrough infections and deaths in vaccinated individuals arise that cause a reduction in the daily vaccine uptake rate from its baseline value. For a fully effective intervention (effectiveness of 100%), note that the daily vaccine uptake rate would be unchanged by the outbreak information.

For behaviourally-homogeneous populations, we have a simplified form of the vaccination uptake rate (Eq. (9)):

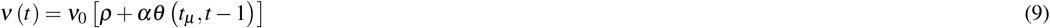

Since the equation above is specific to behaviourally-homogeneous populations, only global outbreak information awareness is explored leaving *θ* to be replaced by *θ*_*GC*_ or *θ*_*GD*_.

Based on the range of behavioural parameters we considered in our simulations, our behavioural function *ν*_*i*_ (*t*) produced daily vaccine uptake rates ranging from 0 to 0.02, or 0% to 2% of the subpopulation being vaccinated per day (Fig. 2). Between 14 December 2020 and 07 April 2021 in the UK, the daily share of the population receiving a COVID-19 vaccine dose ranged from approximately 0.1% to 0.9% [45]. Both extremes of this range of daily vaccine uptake rates are included within our parameter space.

**Figure 2.**
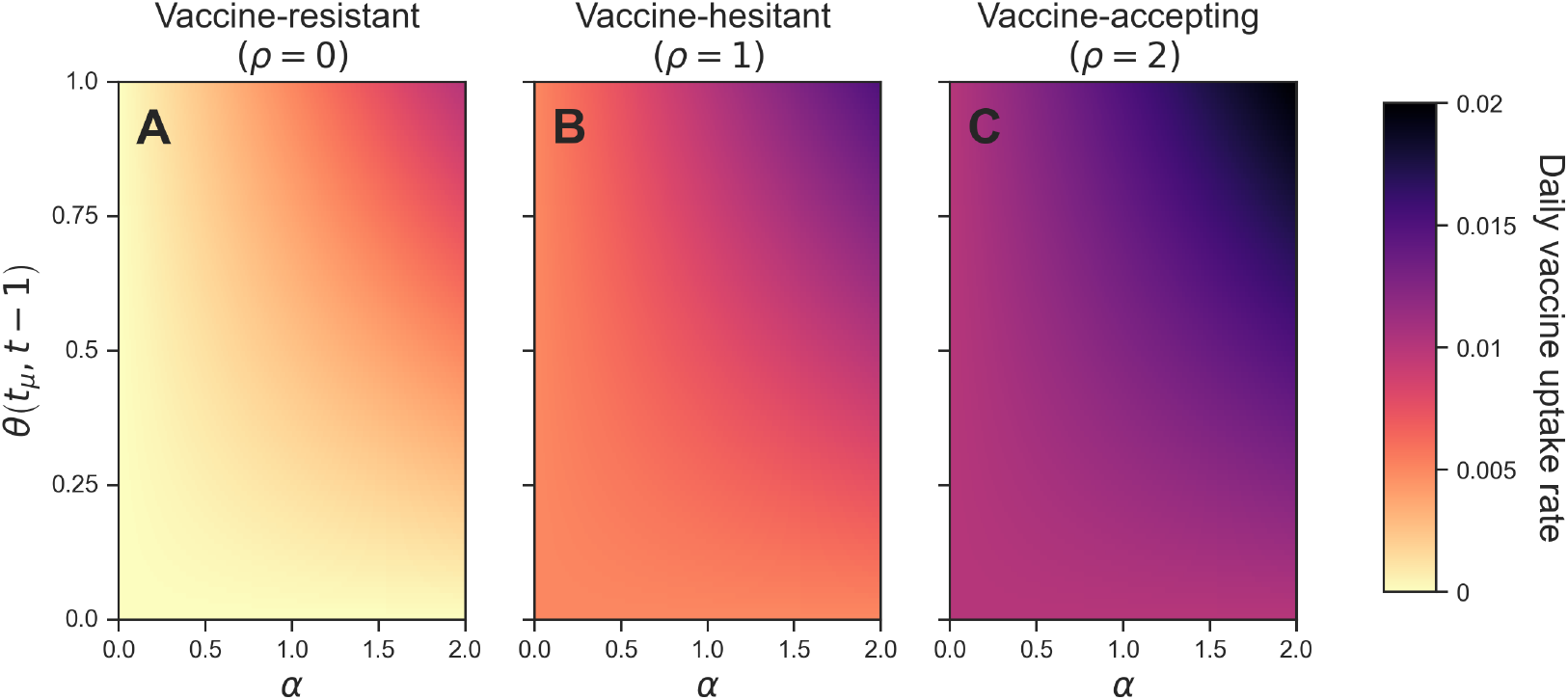
Dependency of the daily vaccine uptake rate on the behavioural function *ν* _*i*_ (*t*). We display three examples at different levels of vaccine opinion, *ρ*_*i*_, which we use to define our three behavioural groups in our heterogeneous simulations: vaccine-resistant (*ρ* = 0); **(B)** vaccine-hesitant (*ρ* = 1); and **(C)** vaccine-accepting (*ρ* = 2). The daily vaccine uptake rate (colour) is shown for different values of information sensitivity, *α*, and outbreak information, *θ*_*i*_ *t*_*µ*_, *t* − 1. The outbreak information is calculated using the equations in Eqs. (6) and (7).

### Pathogen-specific model parameterisation

To disentangle how incorporating behaviour into our model might impact systems of varying transmission potential and rate of infection fatality, we parameterised the model for three distinct human respiratory pathogen systems that differed in terms of these attributes: pandemic influenza, SARS-CoV-2 and Ebola. We based the disease parameter values, including the basic reproduction number, *R*_0_, latent and infectious periods and infection fatality risks on estimates from the existing scientific literature (Table 1). Pandemic influenza has an *R*_0_ of approximately 1.5 [34], with relatively short latent and infectious periods of 2 and 5 days [37] (Table 1). In contrast, the SARS-CoV-2 wild-type variant has an estimated *R*_0_ of approximately 3 [35] with longer latent and infectious periods (relative to our parameter estimates for pandemic influenza) of 5 and 7 days [38] (Table 1). Lastly, Ebola has an *R*_0_ around 2 with latent and infectious periods of 2 and 7 days [36]. Compared with the other two pathogens, Ebola has a much larger probability of death due to infection (*m*) at 0.39 [41], with probability of death due to infection with SARS-CoV-2 at 6.38×10^−3^ [40] and pandemic influenza at 1×10^−4^ [39].

### Simulation overview

For all model simulations we used an overall population size of 100,000 individuals. We initialised infection with one unvaccinated infectious individual on day 0. For the heterogeneous simulations described below, the one initial infection was distributed according to the proportion assigned to each behavioural subpopulation. We ran the simulations until the number of active infections was fewer than one. Each sub-analysis had a bespoke simulation set, summarised in Table 3 and with further details below. We wrote the model code in Python 3.11.7, with the model code available at https://github.com/rachelseibel/outbreak_information_model.

**Table 3.**
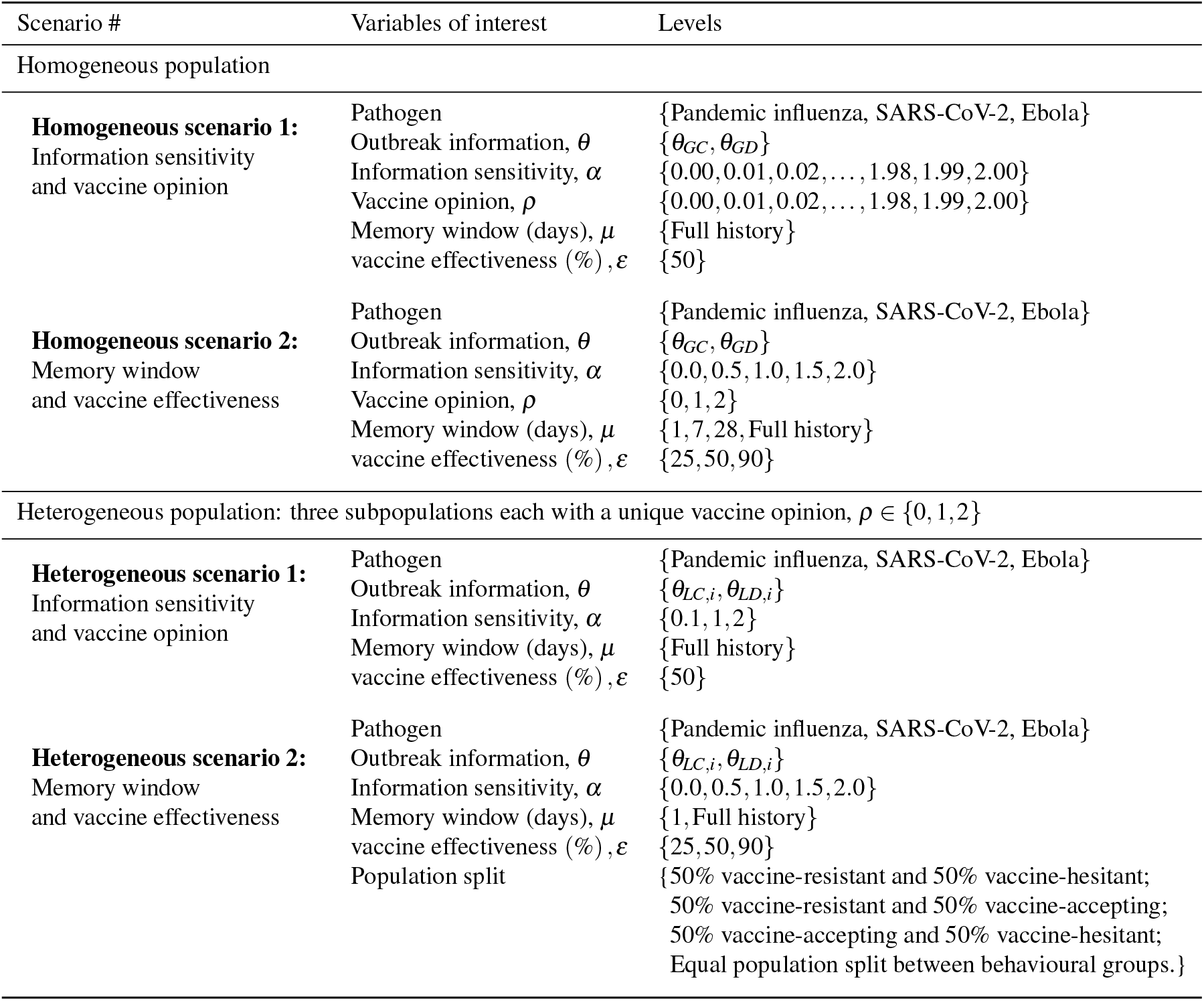
Simulation outline specifying levels of behavioural variables for four scenarios. For each scenario, we specify the population type (homogeneous or heterogeneous), the variables of interest and the levels we simulated of these variables. Each scenario simulation can be thought of as a unique combination of the variables of interest and their respective levels. It should be emphasised that the value for vaccine effectiveness, *ε*, is the same for the two different vaccine actions – infection blocking (*ε*_*s*_) and reduction of severe disease (*ε*_*m*_). More information regarding the simulation parameters and variables can be found in Tables 1 and 2 and Eqs. (6) and (7).

| Scenario # | Variables of interest | Levels |
| --- | --- | --- |
| Homogeneous population |  |  |
| **Homogeneous scenario 1:**<br>Information sensitivity<br>and vaccine opinion | Pathogen<br>Outbreak information, $\theta$<br>Information sensitivity, $\alpha$<br>Vaccine opinion, $\rho$<br>Memory window (days), $\mu$<br>vaccine effectiveness (%), $\epsilon$ | {Pandemic influenza, SARS-CoV-2, Ebola}<br>{$\theta_{GC}, \theta_{GD}$}<br>{0.00, 0.01, 0.02, ..., 1.98, 1.99, 2.00}<br>{0.00, 0.01, 0.02, ..., 1.98, 1.99, 2.00}<br>{Full history}<br>{50} |
| **Homogeneous scenario 2:**<br>Memory window<br>and vaccine effectiveness | Pathogen<br>Outbreak information, $\theta$<br>Information sensitivity, $\alpha$<br>Vaccine opinion, $\rho$<br>Memory window (days), $\mu$<br>vaccine effectiveness (%), $\epsilon$ | {Pandemic influenza, SARS-CoV-2, Ebola}<br>{$\theta_{GC}, \theta_{GD}$}<br>{0.0, 0.5, 1.0, 1.5, 2.0}<br>{0, 1, 2}<br>{1, 7, 28, Full history}<br>{25, 50, 90} |
| Heterogeneous population: three subpopulations each with a unique vaccine opinion, $\rho \in \{0, 1, 2\}$ | | |
| **Heterogeneous scenario 1:**<br>Information sensitivity<br>and vaccine opinion | Pathogen<br>Outbreak information, $\theta$<br>Information sensitivity, $\alpha$<br>Memory window (days), $\mu$<br>vaccine effectiveness (%), $\epsilon$ | {Pandemic influenza, SARS-CoV-2, Ebola}<br>{$\theta_{LC,i}, \theta_{LD,i}$}<br>{0.1, 1, 2}<br>{Full history}<br>{50} |
| **Heterogeneous scenario 2:**<br>Memory window<br>and vaccine effectiveness | Pathogen<br>Outbreak information, $\theta$<br>Information sensitivity, $\alpha$<br>Memory window (days), $\mu$<br>vaccine effectiveness (%), $\epsilon$<br>Population split | {Pandemic influenza, SARS-CoV-2, Ebola}<br>{$\theta_{LC,i}, \theta_{LD,i}$}<br>{0.0, 0.5, 1.0, 1.5, 2.0}<br>{1, Full history}<br>{25, 50, 90}<br>{50% vaccine-resistant and 50% vaccine-hesitant;<br>50% vaccine-resistant and 50% vaccine-accepting;<br>50% vaccine-accepting and 50% vaccine-hesitant;<br>Equal population split between behavioural groups.} |

#### Homogeneous population scenarios

In these two homogeneous scenarios, within each simulation the entire population had the same behavioural parameter values. We considered the sensitivity of epidemiological outcomes for 6 unique combinations of pathogen (pandemic influenza, SARS-CoV-2 and Ebola) and outbreak information (global cases, global deaths) - we refer to each of these combinations as a ‘batch’. We expand below on the behavioural parameter values considered under each scenario.

##### Homogeneous scenario 1: Influence of vaccine opinion and information sensitivity in a homogeneous population

In our initial analysis, for each of the six batches (combination of pathogen and outbreak information) we considered the sensitivity of cumulative cases, cumulative deaths and epidemic duration to vaccine opinion and information sensitivity. We performed 40,401 simulations per batch, one simulation for each combination of vaccine opinion, *ρ*, ranging from 0 to 2 (with an increment of 0.01), and information sensitivity, *α*, ranging from 0 to 2 (with an increment of 0.01). This resulted in a total of 242,406 simulations.

##### Homogeneous scenario 2: Influence of memory window and vaccine effectiveness in a homogeneous population

We next considered the sensitivity of cumulative cases and cumulative deaths to memory window and vaccine effectiveness, across different levels of pathogen, outbreak information, vaccine opinion and information sensitivity. Within each batch, we had 15 combinations of vaccine opinion (*ρ*∈ *{*0, 1, 2}), information sensitivity (*α* ∈{0, 0.5, 1, 1.5, 2}). We selected these parameter values to reasonably span the range of parameter space when considering the two parameters. Furthermore, to then assess sensitivity of modelled epidemiological outcomes to memory window and vaccine effectiveness, for each of these 15 combinations we also considered 12 combinations of memory window length (*µ* ∈ {1 day, 7 day, 28 day, full outbreak history}) and vaccine effectiveness (*ε* ∈ {25%, 50%, 90%}). This gave a total of 1,080 simulations for this scenario.

#### Heterogeneous population scenarios

In these two heterogeneous scenarios, within each simulation the population was split between three subpopulations that each had a unique vaccine opinion to represent vaccine-resistant (*ρ* = 0), vaccine-hesitant (*ρ* = 1) and vaccine-accepting (*ρ* = 2) subpopulations. We stratified group occupancy to a resolution of 5% - a subjective choice that ensured the total required computational time for running the collection of scenarios was manageable, whilst providing a resolution that would be capable of revealing trends between vaccine opinion group composition and epidemiological outcomes. Unique combinations of occupancy (population split) across the three groups that summed to unity resulted in 231 heterogeneous vaccine opinion group configurations. We considered the sensitivity of epidemiological outcomes for each of 6 combinations of pathogen (pandemic influenza, SARS-CoV-2 and Ebola) and outbreak information (local cases, local deaths). Overall, we considered 1,386 unique combinations of pathogen, outbreak information and population split. We expand below on the behavioural parameter values considered under each scenario.

##### Heterogeneous scenario 1: Influence of vaccine opinion and information sensitivity in a heterogeneous population

In this scenario, we considered the effect of information sensitivity on cumulative cases and cumulative deaths across different levels of pathogen, outbreak information and population split. For each of the 1,386 combinations of pathogen, outbreak information and population split, we explored sensitivity of epidemiological outcomes to 3 specific information sensitivity values, with *α* ∈ {0, 1, 2}. We therefore carried out a total of 4,158 simulations for this scenario.

##### Heterogeneous scenario 2: Influence of memory window and vaccine effectiveness in a heterogeneous population

We lastly considered how memory window and vaccine effectiveness impacted cumulative cases and cumulative deaths across 120 different combinations of pathogen, outbreak information, population split and information sensitivity (*α* ∈{0, 0.5, 1, 1.5, 2}). We specifically explored four population splits or behavioural configurations of interest: (i) 50% vaccine-resistant and 50% vaccine-hesitant; (ii) 50% vaccine-resistant and 50% vaccine-accepting; (iii) 50% vaccine-hesitant and 50% vaccine-accepting; and (iv) equal population split between vaccine-resistant, vaccine-hesitant and vaccine-accepting groups. We then considered six combinations of memory window length (*µ*∈{1 day, full outbreak history}) and vaccine effectiveness (*ε* ∈{25%, 50%, 90%}). This gave a total of 720 simulations for this scenario. In the homogeneous scenarios, we found a weak effect of memory window across the 1-day, 7-day, 28-day and full outbreak history values, therefore, we decided to present the extremes of this range for this heterogeneous scenario.

## Results

### Homogeneous scenario 1: Preference for a cases- or deaths-driven behavioural reaction for improved epidemiological outcomes are pathogen-dependent

We first studied the influence of vaccine opinion (*ρ*) and information sensitivity (*α*) in a homogeneous population on cumulative cases, cumulative deaths and epidemic duration. These analyses were done with consideration to our three selected human pathogens: pandemic influenza, SARS-CoV-2 and Ebola, and two types of global outbreak information: global cases (*θ*_*GC*_) and global deaths (*θ*_*GD*_).

Across all pathogens and types of global outbreak information, cumulative cases and cumulative deaths decreased as vaccine opinion and information sensitivity increased (Figures 3 and 4). In other words, cumulative epidemiological metrics were worse when behavioural parameters were turned off. When outbreak information was based on global cases, cumulative cases ranged from approximately 72 to 94,000 cases across all pathogens whilst cumulative deaths ranged from approximately 0.005 to 35,000 deaths. When outbreak information was based on global deaths, cumulative cases ranged from approximately 91 to 94,000 cases across all pathogens whilst cumulative deaths ranged from approximately 0.006 to 35,000 deaths. For SARS-CoV-2 and Ebola, the epidemic duration in days increased as vaccine opinion and information sensitivity increased. For pandemic influenza, epidemic duration did not follow a linear relationship with the behavioural variables of interest. Instead, very low and high levels (0-0.1, 1.7-2) of vaccine opinion and information sensitivity led to shorter epidemic durations (less than 300 days) whilst mid-range levels (0.1-1.7) of vaccine opinion and information sensitivity led to longer epidemic durations (300-980 days) (Figures 3 and 4). The low levels of vaccine opinion led to short outbreaks due to rapid infection spread, whilst high levels of vaccine opinion led to outbreaks which were controlled quickly.

**Figure 3.**
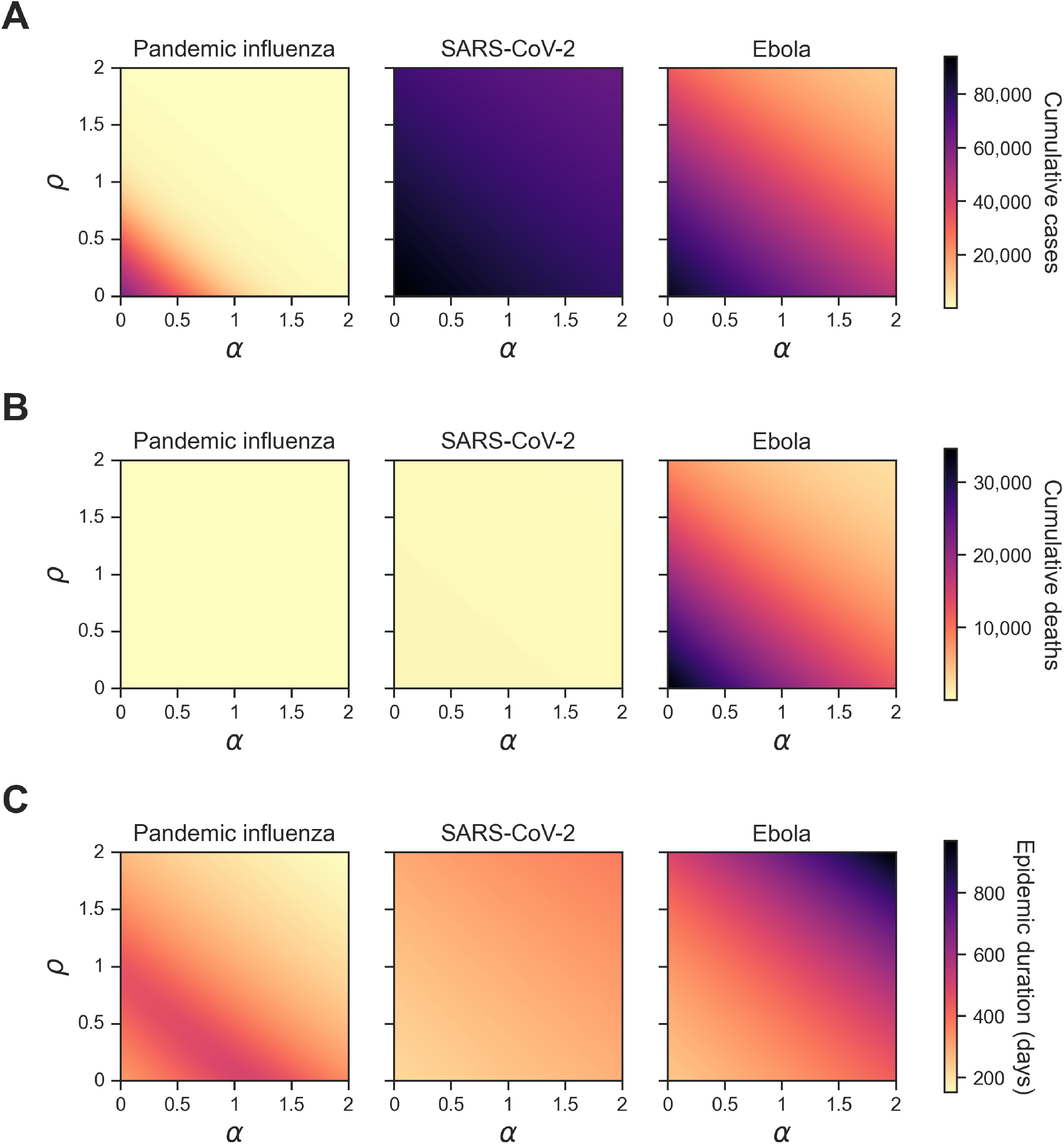
Homogeneous scenario 1. Summary epidemiological metrics by pathogen when outbreak information was based on global cases (*θ* _*GC*_). Each panel shows a given epidemiological statistic across different levels of information sensitivity (*α*) (x-axis, ranging from 0 to 2) and different levels of vaccine opinion (*ρ*) (y-axis, ranging from 0 to 2). Each column corresponds to a different pathogen: pandemic influenza (column one), SARS-CoV-2 (column two) and Ebola (column three). Each row displays one of the three summary epidemiological measures with a shared colour bar: **(A)** cumulative cases; cumulative deaths; **(C)** epidemic duration (days). Darker shading corresponds to higher values for each epidemiological metric. The cumulative outcomes vary between pathogen systems, with pandemic influenza being particularly distinct from the other two pathogens. The corresponding plot for outbreak information based on global deaths (*θ*_*G****D***_) can be found in Figure S1.

**Figure 4.**
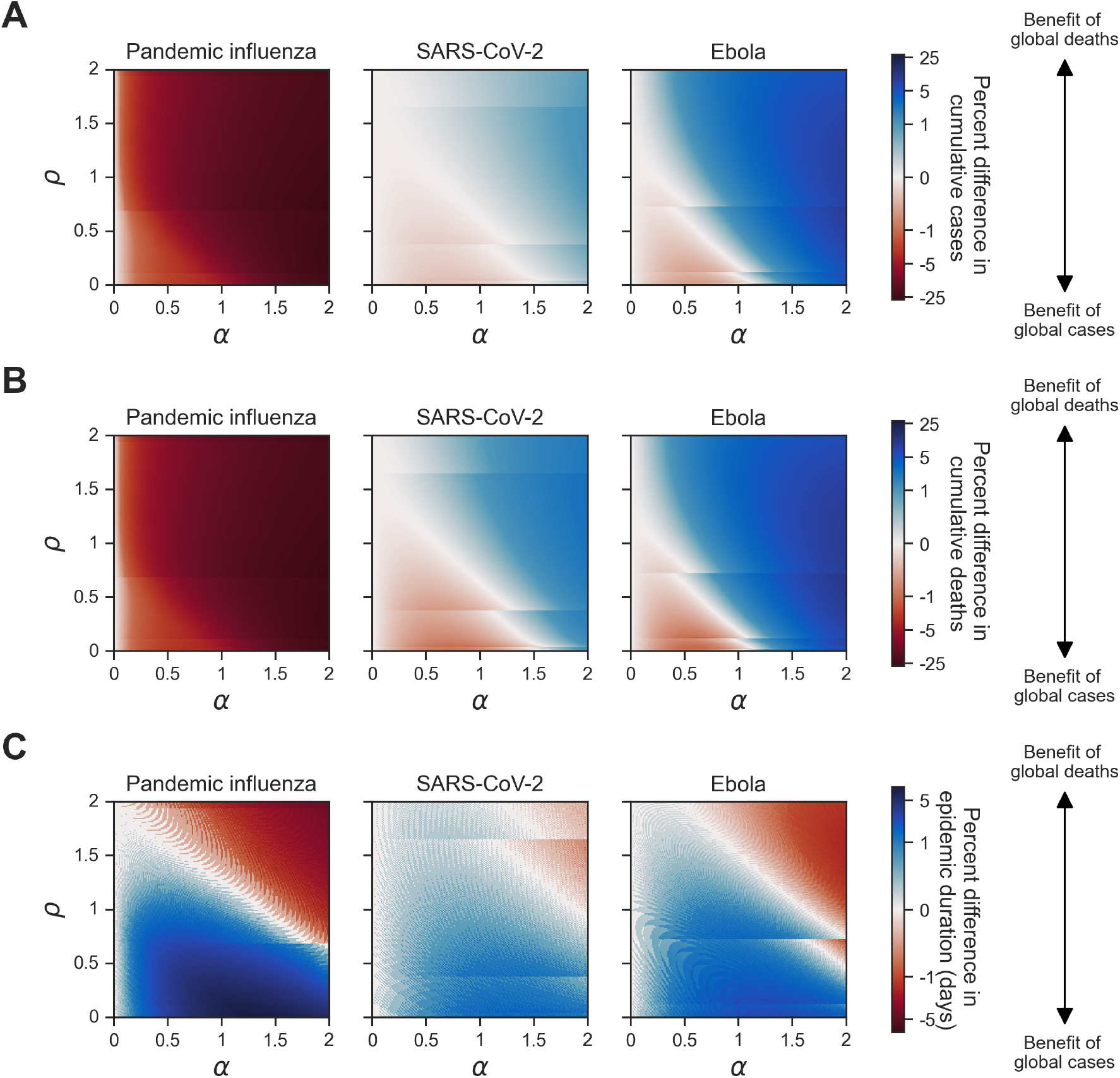
Homogeneous scenario 1. Percent difference in summary epidemiological metrics between simulations with outbreak information based on global cases (*θ* _*GC*_) and global deaths (*θ* _*GD*_). Each panel shows a given epidemiological statistic across different levels of information sensitivity (*α*) (x-axis, ranging from 0 to 2) and vaccine opinion (*ρ*) (y-axis, ranging from 0 to 2). Each column corresponds to a different pathogen: pandemic influenza (column one), SARS-CoV-2 (column two) and Ebola (column three). Each row shows the percent difference between simulations where the global cases (*θ*_*GC*_) and global deaths (*θ*_*GD*_) behavioural functions were used: **(A)** cumulative cases; **(B)** cumulative deaths; **(C)** epidemic duration (days). The shared colour bar is on a log scale. Red shading corresponds to scenarios where outbreak information based on global cases led to lower values in the corresponding epidemiological metric compared with global cases. In other words, red shading corresponds to a benefit of global cases and blue shading corresponds to a benefit of global deaths as outbreak information (*θ*). The striations at certain values of *ρ* are a numerical artefact arising due to precision of the vaccine uptake rate; improving upon the variable precision would lead to longer computational times (Equation (9)) causing slight shifts in behaviour at early outbreak stages.

We then considered the percent difference in cumulative outbreak measures between simulations with outbreak information based on global cases (*θ*_*GC*_) when compared to simulations with outbreak information based on global deaths (*θ*_*GD*_). Percent difference was calculated by the following equation:

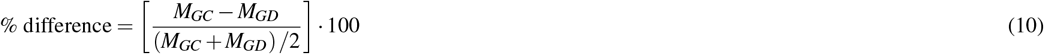

where *M* was the epidemiological metric of interest for the respective types of outbreak information.

For cumulative cases and cumulative deaths, pandemic influenza showed patterns distinct from SARS-CoV-2 and Ebola; for pandemic influenza the percent difference in cumulative cases and cumulative deaths was negative across all levels of vaccine opinion and information sensitivity, which showed a benefit of outbreak information based on global cases (*θ*_*GC*_) (Figure 4A,B). These percent differences were also quite large compared with the other two pathogens because the values of cumulative cases and deaths were extremely small (cumulative deaths were less than 1). For SARS-CoV-2 and Ebola, the percent difference in cumulative cases and cumulative deaths was negative for low levels of vaccine opinion and information sensitivity (0-1.5) and positive for high levels of vaccine opinion and information sensitivity (1.6-2). Therefore, for low levels of vaccine opinion and information sensitivity, we observed a benefit of outbreak information based on global cases (*θ*_*GC*_). Similarly, for high levels of vaccine opinion and information sensitivity, we observed a benefit of outbreak information based on global deaths (*θ*_*GD*_) (Figure 4A,B). For all pathogens, the percent differences in epidemic duration were positive (i.e. a benefit of outbreak information based on global cases (*θ*_*GC*_)) for low levels of vaccine opinion and information sensitivity (0-1.5) and negative (i.e. a benefit of outbreak information based on global deaths (*θ*_*GD*_)) for high levels of vaccine opinion and information sensitivity (1.5-2) (Figure 4C).

Given that the cumulative outbreak measures differed in pattern by pathogen, we examined temporal dynamics showing the cumulative cases, cumulative deaths and cumulative vaccinations across time in days for a vaccine opinion (*ρ*) of 2 and information sensitivity (*α*) of 2 (Figure 5). These behavioural parameters were highlighted since the greatest differences between outbreak information types fell in this parameter combination. For pandemic influenza, cumulative cases and cumulative deaths were consistently lower when outbreak information was based on global cases (*θ*_*GC*_) compared with global deaths (*θ*_*GD*_) (Figure 5A,B). This contrasted with SARS-CoV-2 and Ebola, where cumulative cases and cumulative deaths were instead lower for global cases (*θ*_*GC*_) at earlier time points, but higher at later time points compared with global deaths (*θ*_*GD*_). Across all pathogens, due to reported deaths being a lagged measure compared with reported cases, having outbreak information based on global deaths resulted in cumulative vaccinations also being lagged at the start of the outbreak relative to cumulative vaccinations under outbreak information based on global cases (Figure 5C).

**Figure 5.**
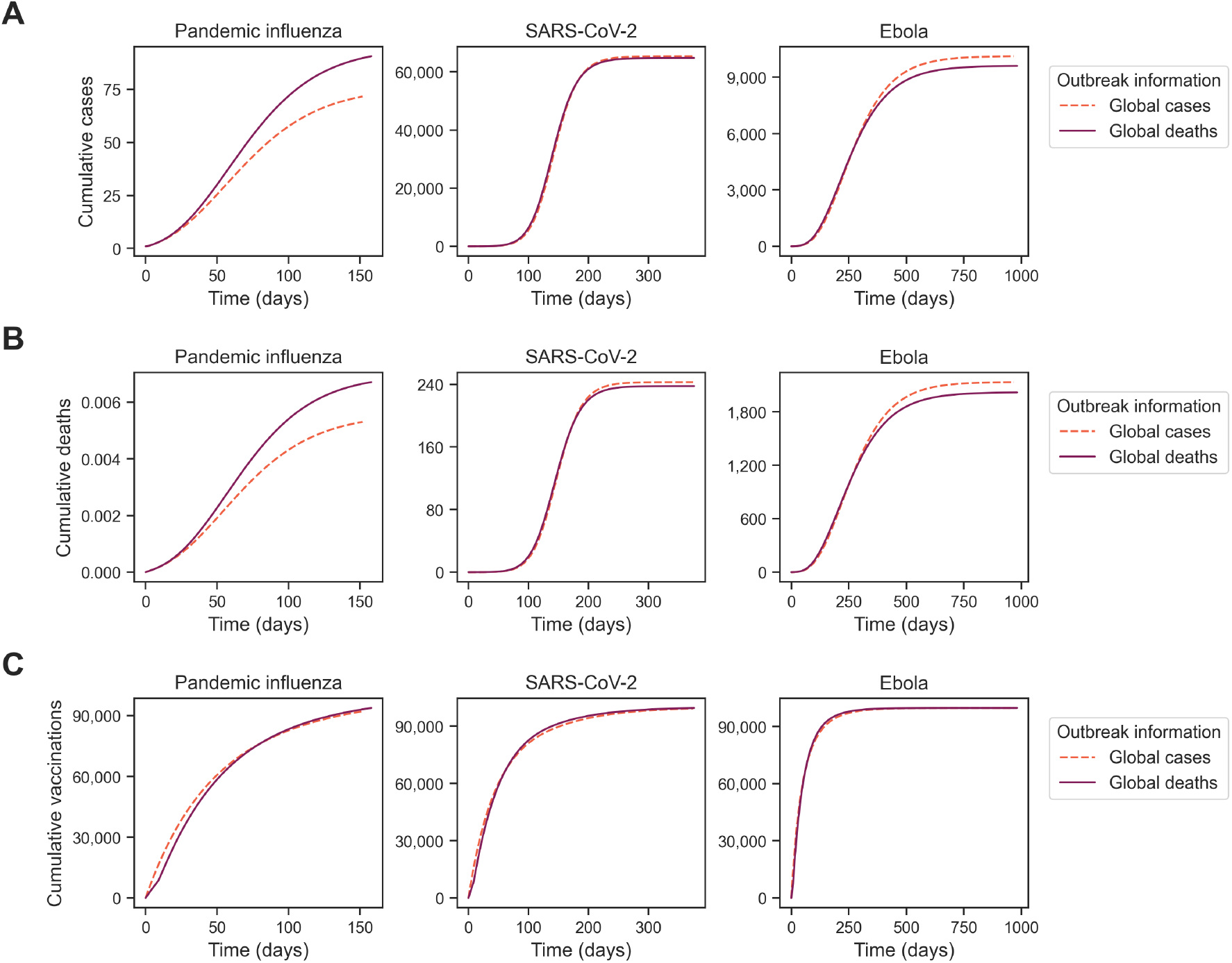
Homogeneous scenario 1. Temporal outbreak measures between simulations with outbreak information based on global cases (*θ* _*GC*_) and global deaths (*θ* _*GD*_). For a vaccine opinion (*ρ*) of 2 and information sensitivity (*α*) of 2, we display our epidemiological metrics across time (x-axis): **(A)** cumulative cases, **(B)** cumulative deaths and **(C)** cumulative vaccinations. Each line indicates a different level of outbreak information (*θ*): global cases (pink dashed line) and global deaths (purple solid line). Note the differences in cumulative metrics through time by the underlying outbreak information considered.

### Homogeneous scenario 2: Preference for a cases- or deaths-driven behavioural reaction for improved epidemiological outcomes are vaccine effectiveness-dependent

We next considered the sensitivity of cumulative cases and cumulative deaths to memory window and vaccine effectiveness in a behaviourally homogeneous population. The important takeaway from these simulations was that outbreak information preference was sensitive to vaccine effectiveness alongside the underlying pathogen-specific parameters identified in the homogeneous scenario 1 simulations.

For all pathogens at a full history memory window (*µ*), we found that cumulative cases decreased as vaccine effectiveness (*ε*) increased (Figure 6A). At a vaccine effectiveness of 25%, the percent difference in cumulative cases between global cases and global deaths was small (0-2%) across all pathogens (Figure 6B). For pandemic influenza, the percent difference in cumulative cases between global cases and global deaths was negative (benefit of global cases) for a vaccine effectiveness of 50% (ranging from 0% to -26%) and 90% (ranging from 0% to -61%). For SARS-CoV-2 and Ebola at a vaccine effectiveness of 50%, the percent difference in cumulative cases was negative («-1%) when vaccine opinion was 0 and information sensitivity was 1 (benefit of global cases) and zero or positive (0-8%) for all other combinations of vaccine opinion and information sensitivity. Across all pathogens at a vaccine effectiveness of 90%, the percent difference in cumulative cases was zero or negative: 0 to -61% for pandemic influenza, 0 to -39% for SARS-CoV-2 and 0 to -65% for Ebola. The trends in epidemiological metrics were similar when considering cumulative deaths (Figures S2 and S4).

**Figure 6.**
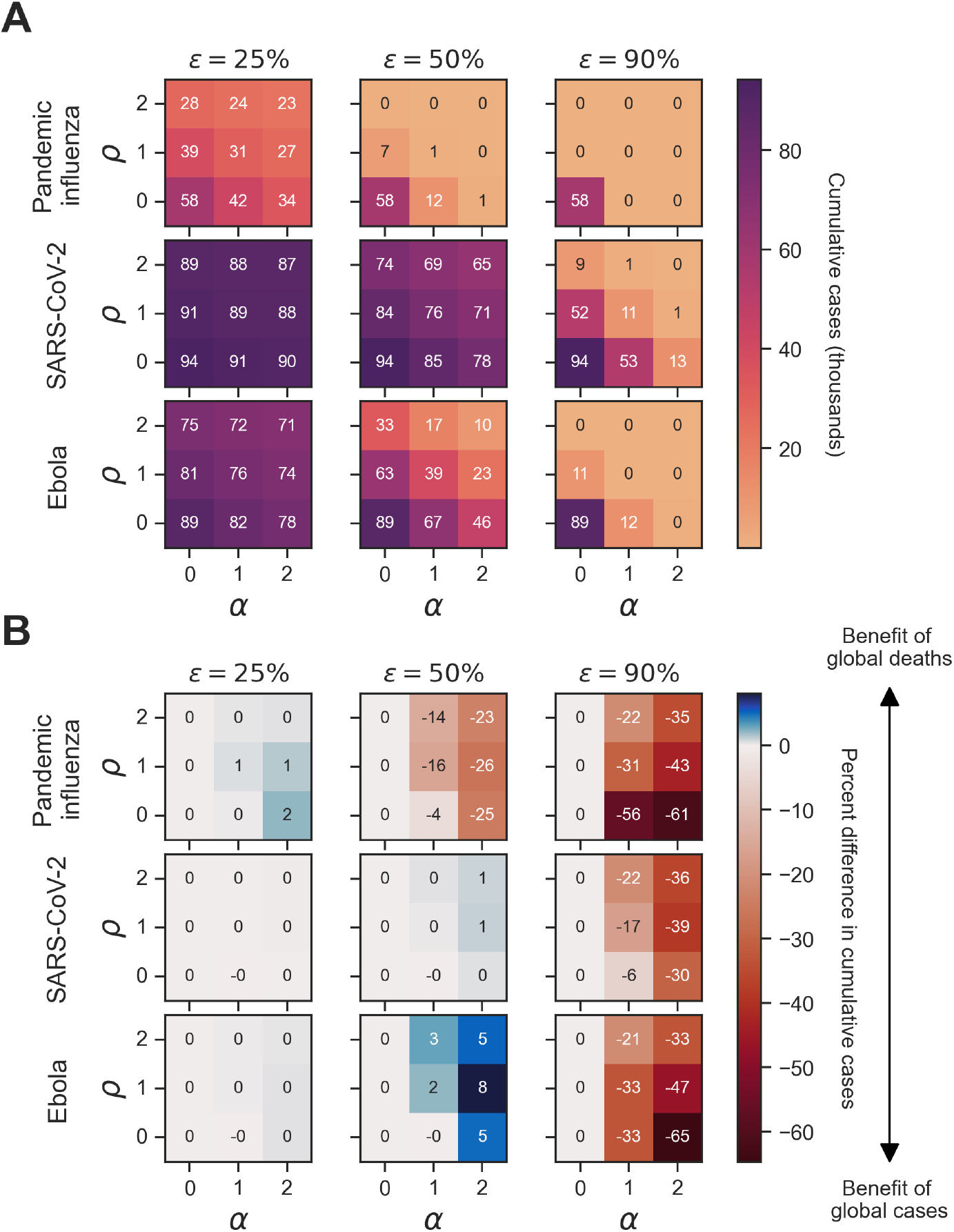
Homogeneous scenario 2. Sensitivity of cumulative cases to vaccine effectiveness across pathogens and type of outbreak information for a full history memory window. For each panel, cumulative cases are presented across the three pathogens of interest (pandemic influenza, SARS-CoV-2, Ebola) alongside three vaccine efficacies (*ε* ∈ 25%, 50%, 90%) of interest. Panel **(A)** shows simulations where the outbreak information was global cases as well as the corresponding cumulative cases in thousands for each unique combination of pathogen, vaccine effectiveness (*ε*), vaccine opinion (*ρ*) and information sensitivity (*α*). Dark purple hues correspond to more cumulative cases whilst light orange hues correspond to fewer cumulative cases. Panel **(B)** shows the percent difference in cumulative cases between simulations where the outbreak information was global cases (*θ*_*GC*_) and global deaths (*θ*_*GD*_) for each unique combination of pathogen, vaccine effectiveness (*ε*), vaccine opinion (*ρ*) and information sensitivity (*α*). Blue hues correspond to positive percent differences in cumulative cases (representing a benefit of global deaths) whilst red hues correspond to negative percent differences in cumulative cases (representing a benefit of global cases).

We then considered a memory window (*µ*) of 1 day and found that the benefits of outbreak information types remained the same across different levels of vaccine opinion and information sensitivity. However, the magnitude of the percent differences in cumulative epidemiological metrics differed. For pandemic influenza at a vaccine effectiveness of 90%, the percent difference in cumulative cases between global cases and global deaths ranged from 0 to -89% compared with 0 to -61% at a full history memory window (Figure 7B). Despite this variation, the cumulative case numbers are below 500 for both memory windows and therefore the percent differences in cumulative cases are not greatly meaningful. The same is true for 7-day and 28-day memory window values (Figures S6 and S7).

**Figure 7.**
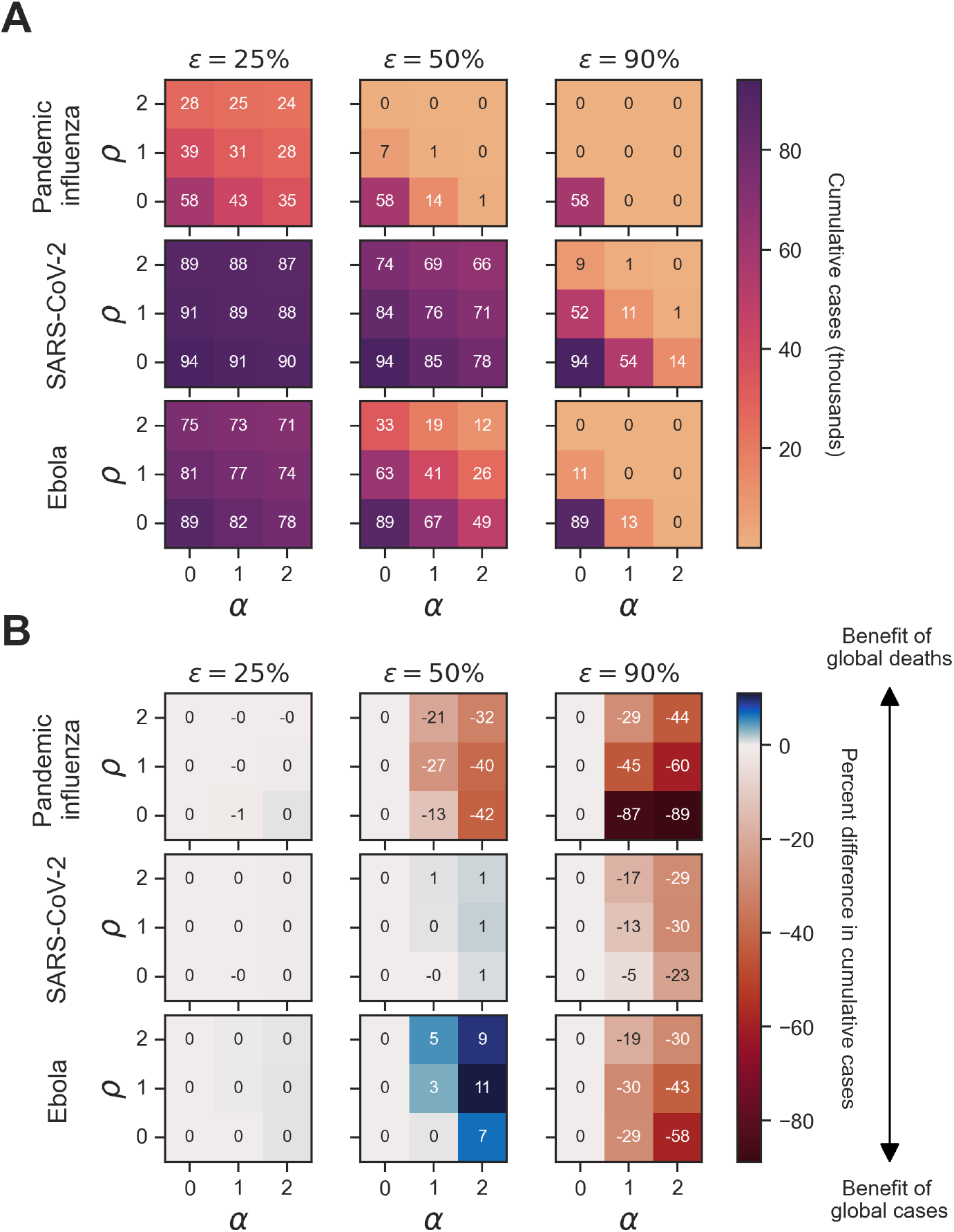
Homogeneous scenario 2. Sensitivity of vaccine effectiveness across pathogens and type of outbreak information for a 1-day memory window. For each panel, cumulative cases are presented across the three pathogens of interest (pandemic influenza, SARS-CoV-2, Ebola) alongside three vaccine efficacies (*ε* ∈ 25%, 50%, 90%) of interest. Panel shows simulations where the outbreak information was global cases as well as the corresponding cumulative cases in thousands for each unique combination of pathogen, vaccine effectiveness (*ε*), vaccine opinion (*ρ*) and information sensitivity (*α*). Dark purple hues correspond to more cumulative cases whilst light orange hues correspond to fewer cumulative cases. Panel **(B)** shows the percent difference in cumulative cases between simulations where the outbreak information was global cases (*θ*_*GC*_) and global deaths (*θ*_*GD*_) for each unique combination of pathogen, vaccine effectiveness (*ε*), vaccine opinion (*ρ*) and information sensitivity (*α*). Blue hues correspond to positive percent differences in cumulative cases (representing a benefit of global deaths) whilst red hues correspond to negative percent differences in cumulative cases (representing a benefit of global cases).

However, for Ebola at a vaccine effectiveness of 50% and global cases as outbreak information, a 1-day memory window (23,000 cases) led to approximately 3,000 more cases compared with a full-history memory window (26,000 cases) (Figures 6 and 7). The percent difference in cumulative cases between global cases and global deaths increased from 8 to 11% for Ebola with the 1-day memory window compared with the full-history memory window (Figure 7).

Inspecting the sensitivity of cumulative cases to vaccine effectiveness (*ε*) in a behaviourally homogeneous population, with global cases as outbreak information, for all pathogens we found that cumulative cases decreased as vaccine effectiveness (*ε*) increased (Figure 8). With respect to increasing vaccine effectiveness (*ε*), cumulative cases decreased more rapidly for pandemic influenza and decreased slowest for SARS-CoV-2. The trends in epidemiological metrics were similar when outbreak information was based on global deaths (*θ*_*GD*_) (Figure 8).

**Figure 8.**
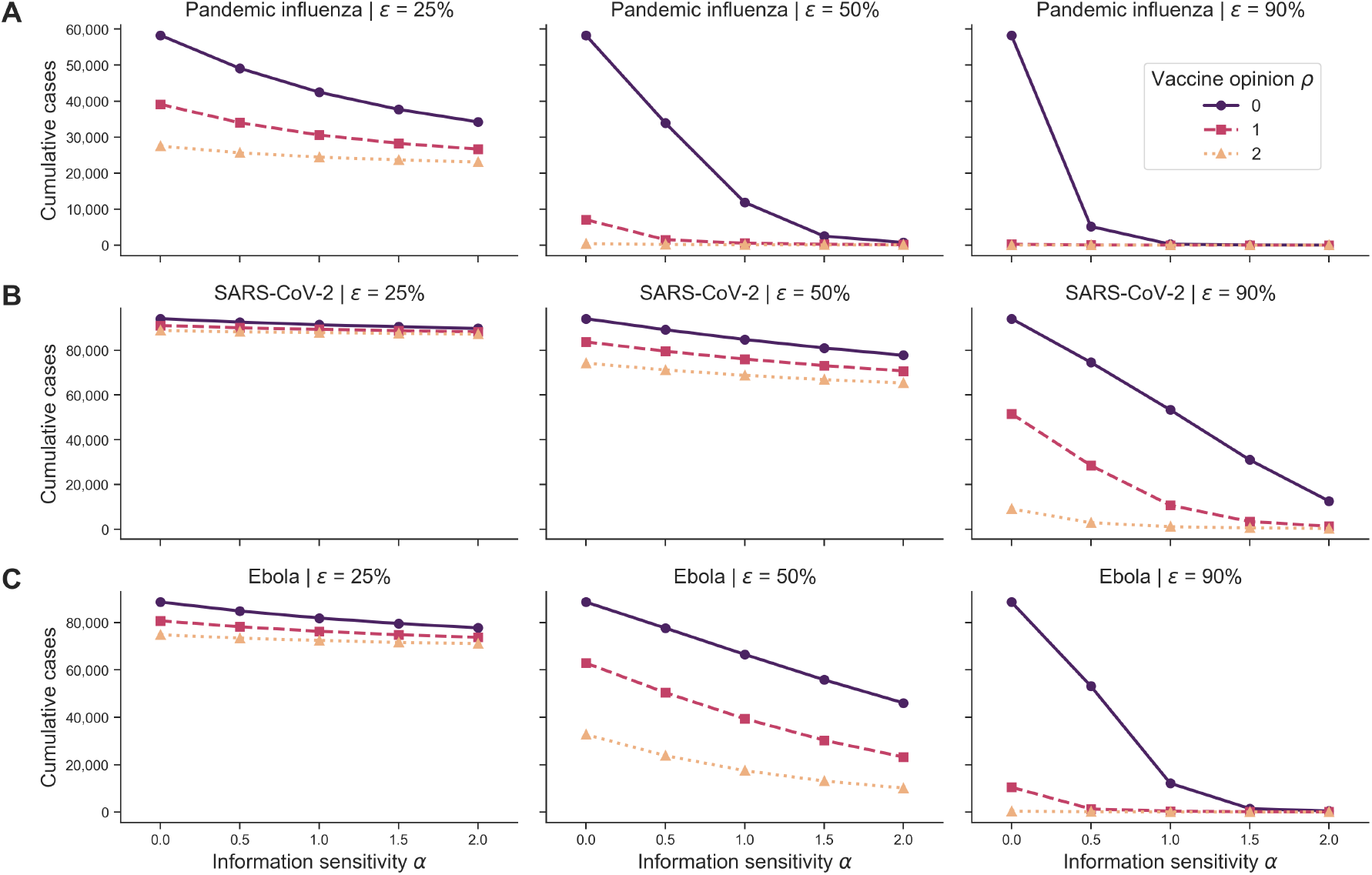
Homogeneous scenario 2. Cumulative cases across pathogen systems and vaccine effectiveness for homogeneous behavioural configurations and outbreak information based on global cases. Each row shows cumulative cases for a given pathogen system: **(A)** pandemic influenza, **(B)** SARS-CoV-2 and **(C)** Ebola. Each column shows a different vaccine effectiveness (*ε*): 25% (first column), 50% (second column) and 90% (third column). Different line types, colours and markers indicate different homogeneous behavioural configurations: vaccine-resistant (*ρ* = 0, purple solid line with circle markers), vaccine-hesitant (*ρ* = 1, pink dashed line with square markers) and vaccine-accepting (*ρ* = 2, orange dotted line with triangle markers). The memory window (*µ*) was fixed at a full history and outbreak information was based on global cases (*θ*_*GC*_).

### Heterogeneous scenario 1: Preference for a cases- or deaths-driven behavioural reaction for improved epidemiological outcomes are pathogen-dependent

To relax our previous assumption of the population being homogeneous with respect to both behavioural-associated parameters *α* and *ρ*, we studied the impact of population splits with multiple levels of vaccine opinion (*ρ*). Similar to the homogeneous scenario 1 outcomes, we found that pandemic influenza exhibited patterns distinct from SARS-CoV-2 and Ebola. We also found that behavioural configuration was important in explaining variations in outbreak severity. For all pathogens, cumulative cases decreased as vaccine opinion and information sensitivity increased (Figure 9).

**Figure 9.**
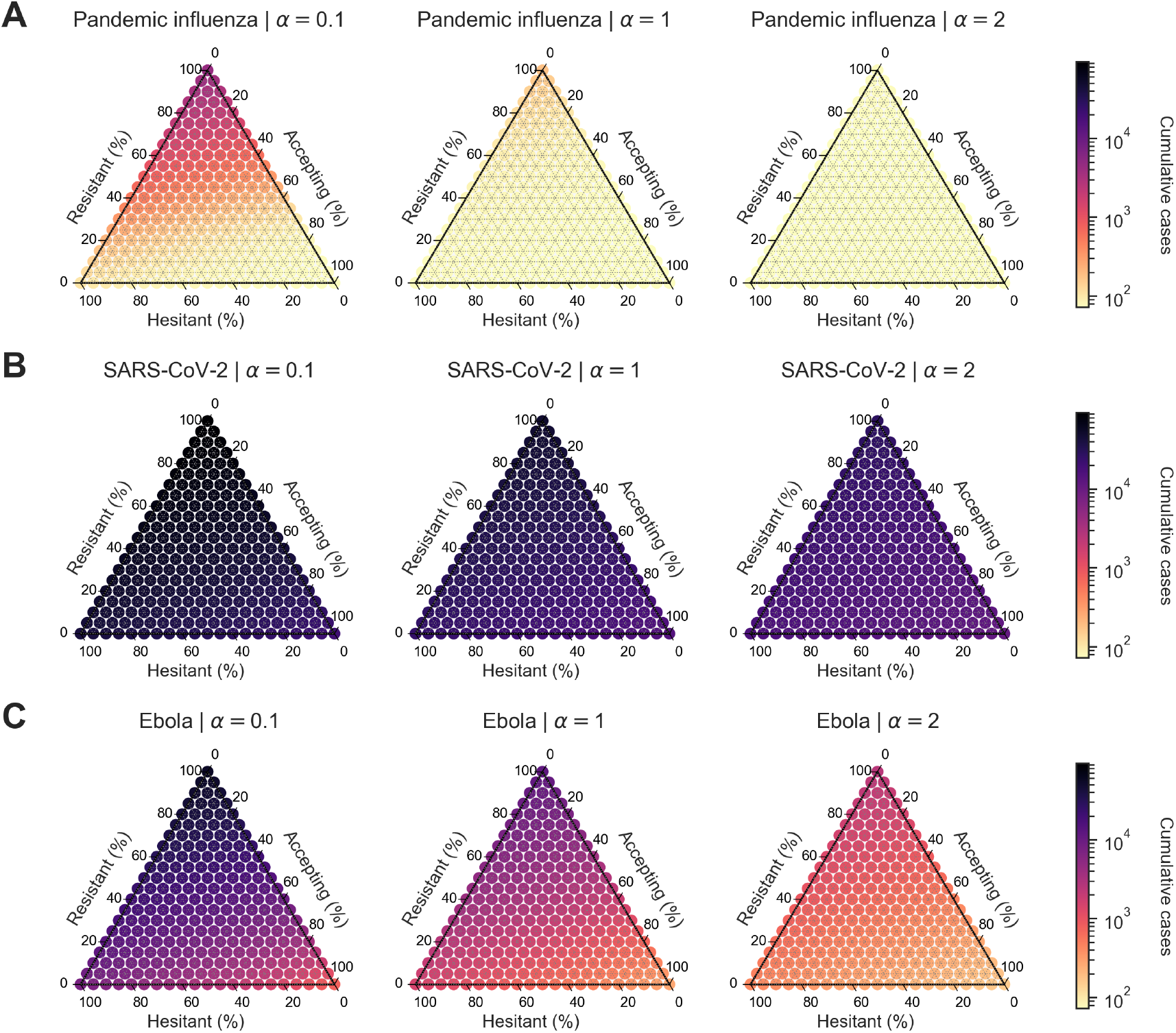
Heterogeneous scenario 1. Cumulative cases across pathogen systems and information sensitivity, with outbreak information based on local cases and vaccine effectiveness of 50%. For each panel, the ternary plot axes show the percentage of the population assigned to each of three behavioural groups: vaccine-resistant (*ρ* = 0), vaccine-hesitant (*ρ* = 1) and vaccine-accepting (*ρ* = 2). Each row shows cumulative cases for a given pathogen: **(A)** pandemic influenza, **(B)** SARS-CoV-2 *α* = 1 (second column), *α* = 2 (third column). Darker colour hues indicate more severe outcomes in terms of cumulative cases. SARS-CoV-2 shows less variable outcomes across information sensitivity compared with the other two pathogens.

We then considered the percent difference in cumulative cases between simulations with outbreak information based on local cases (*θ*_*LC*_) when compared with simulations with outbreak information based on local deaths (*θ*_*LD*_). For pandemic influenza, the percent difference in cumulative cases was negative across all levels of information sensitivity and behavioural configuration (ranging from -0.5% to -37.0%), which showed a benefit of outbreak information based on local cases (*θ*_*LC*_) (Figure 10A). For

**Figure 10.**
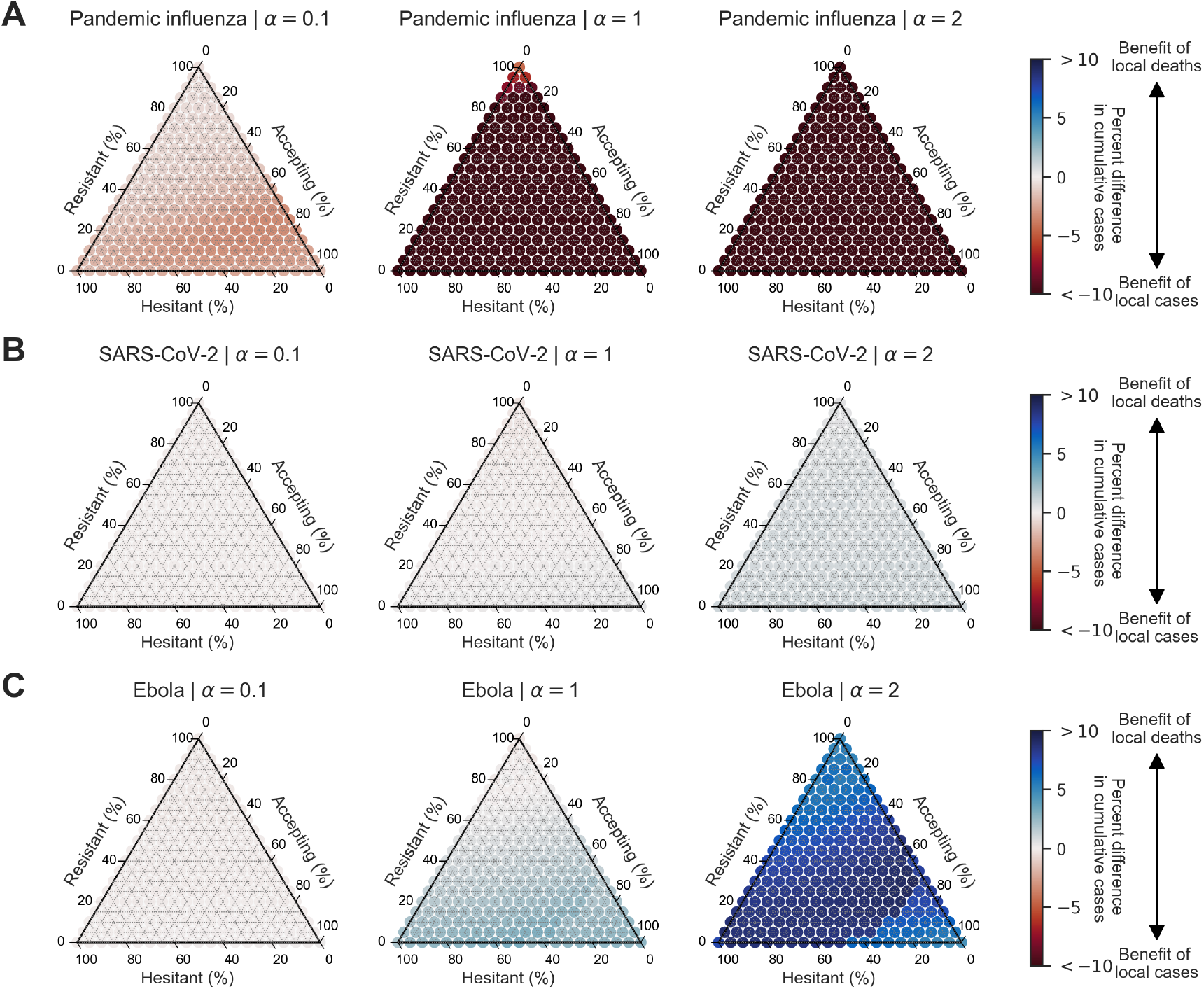
Heterogeneous scenario 1. Percent difference in cumulative cases between local cases and local deaths across pathogen systems and information sensitivity with a vaccine effectiveness of 50%. For each panel, the ternary plot axes show the percentage of the population assigned to each of three behavioural groups: vaccine-resistant (*ρ* = 0), vaccine-hesitant (*ρ* = 1) and vaccine-accepting (*ρ* = 2). Each row shows percent difference in cumulative cases between outbreak information scenarios based on local cases and local deaths for a given pathogen: **(A)** pandemic influenza, **(B)** SARS-CoV-2, **(C)** Ebola. Each column corresponds to a different level of information sensitivity: *α* = 0.1 (first column), *α* = 1 (second column), *α* = 2 (third column). Darker colour hues indicate more severe outcomes in terms of percent difference in cumulative cases.

SARS-CoV-2 and Ebola, there were only slight differences in cumulative cases for mostly resistant populations and positive differences for mostly accepting populations (SARS-CoV-2: ranging from -0.3% to 1.0%; Ebola: ranging from -0.3% to 9.3%). For resistant populations, this indicated no appreciable difference between the two information systems for Ebola and SARS-CoV-2 (with the exception of Ebola when *a* = 2, where local deaths was more informative for the population). For mostly accepting populations, this indicated a benefit of outbreak information based on local deaths (*θ*_*LD*_) (Figure 10B,C).

### Heterogeneous scenario 2: Preference for a cases- or deaths-driven behavioural reaction for improved epidemiological outcomes are vaccine effectiveness-dependent

Our last scenario considered the sensitivity of cumulative cases to memory window (*µ*) and vaccine effectiveness (*ε*) in a behaviourally heterogeneous population with respect to vaccine opinion (*ρ*), across different pathogens, outbreak information types (*θ*) and information sensitivities (*α*).

For all pathogens, we found that cumulative cases decreased as vaccine effectiveness (*ε*) and information sensitivity (*α*) increased in behaviourally heterogeneous populations (Figure 11). Cumulative cases for pandemic influenza decreased most rapidly as vaccine effectiveness (*ε*) increased compared with SARS-CoV-2 and Ebola. Cumulative cases for SARS-CoV-2 decreased most slowly as vaccine effectiveness (*ε*) increased. Across all pathogens, the resistant/hesitant configuration led to the most cumulative cases, followed by the resistant/accepting configuration, the accepting/hesitant configuration and lastly by the equally-split configuration (Figure 11). Overall, SARS-CoV-2 scenarios had the highest cumulative cases (approximately 95,000 cases), followed by Ebola (approximately 85,000 cases) and then pandemic influenza (approximately 50,000 cases). The trends in epidemiological metrics were similar when outbreak information was based on local deaths (*θ*_*LD*_) (Figure S40) as well as when considering cumulative deaths (Figures S38 and S41).

**Figure 11.**
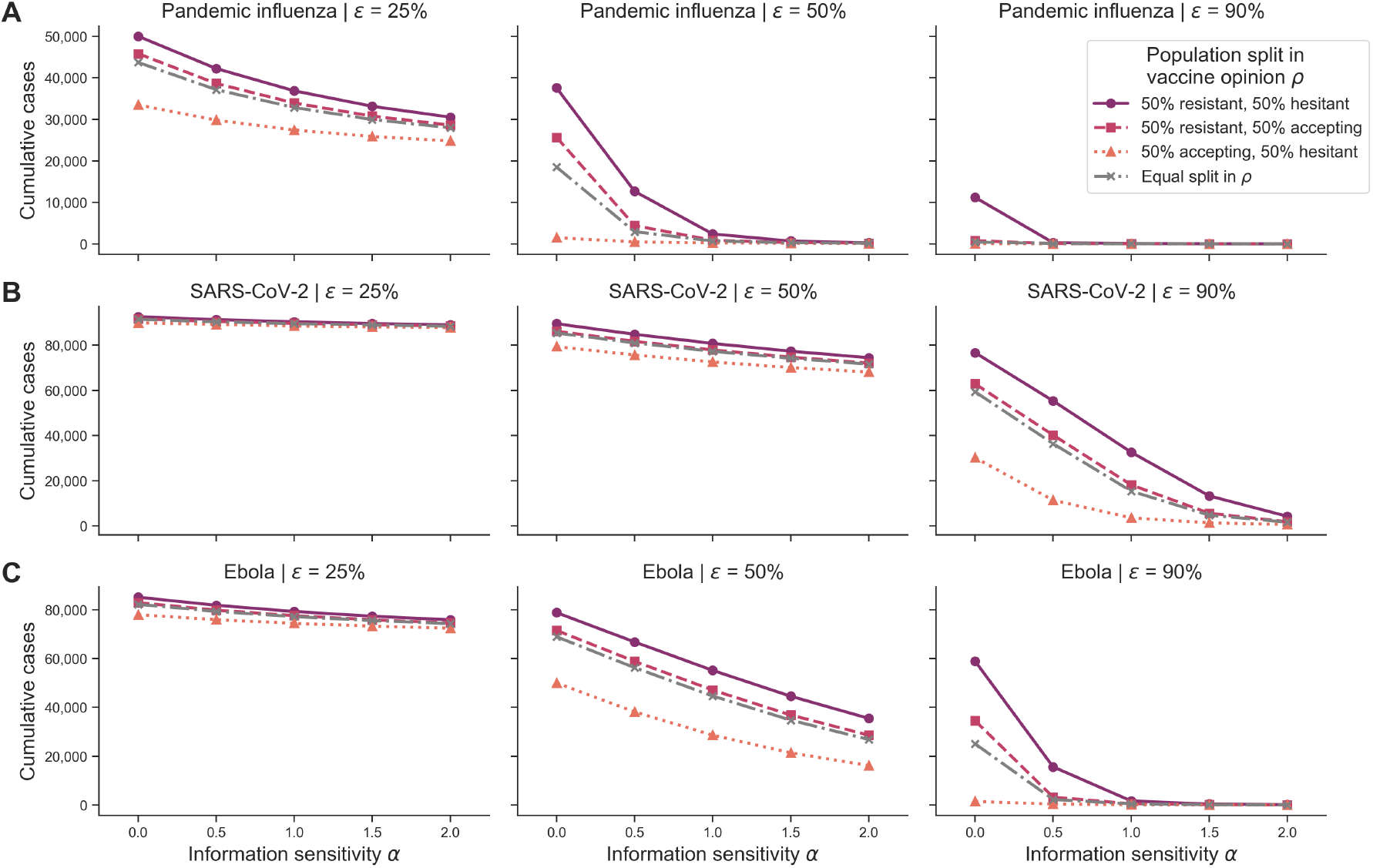
Heterogeneous scenario 2. Cumulative cases across pathogen systems, vaccine effectiveness for heterogeneous behavioural configurations with local cases as outbreak information. Each row shows cumulative cases for a given pathogen system: **(A)** pandemic influenza, **(B)** SARS-CoV-2 and **(C)** Ebola. Each column shows a different vaccine effectiveness (*ε*): 25% (first column), 50% (second column) and 90% (third column). Different line types, colours and markers indicate different mixed behavioural configurations: 50% resistant (*ρ* = 0) and 50% hesitant (*ρ* = 1) (solid purple line with circle markers); 50% resistant (*ρ* = 0) and 50% accepting (*ρ* = 2) (dashed pink line with square markers); 50% accepting (*ρ* = 2) and 50% hesitant (*ρ* = 1) (orange dotted line with triangle markers); and equal split in vaccine opinion (*ρ* ∈ 0, 1, 2, grey dashed-dotted line with x markers). The memory window (*µ*) was fixed at the full history.

In terms of epidemic duration, across all pathogens and outbreak information types for a vaccine effectiveness of 50% and full-history memory window, the accepting/hesitant configurations led to the longest epidemic durations (> 600 days for pandemic influenza), followed by the equal split configuration, then the resistant/accepting configuration and lastly the resistant/hesitant configuration (approximately 225 days for SARS-CoV-2) (Figures S39 and S42). As vaccine effectiveness increased, some behavioural configurations resulted in longer epidemics for mid-range levels of information sensitivity (0.5-1.5). Taking SARS-CoV-2 for instance, with the equal split configuration and a 50% vaccine effectiveness, epidemic duration was longest at an information sensitivity of 1 (approximately 320 days) and shortest at an information sensitivity of 2 (approximately 260 days) (Figure S39B).

We then considered the temporal dynamics of cases for different levels of vaccine effectiveness (*ε*), pathogen system and four behavioural configurations of interest: 50% resistant (*ρ* = 0) and 50% accepting (*ρ* = 2); 50% resistant (*ρ* = 0) and 50% hesitant (*ρ* = 1); 50% accepting (*ρ* = 2) and 50% hesitant (*ρ* = 1); and equal split in vaccine opinion (*ρ* ∈ 0, 1, 2).

For pandemic influenza and across all 50% split configurations, we found that the least accepting subpopulation contributed to more cumulative cases as the outbreak progressed (Figure 12A). For SARS-CoV-2 and Ebola, the least accepting subpopulation contributed to more cumulative cases between 30 and 150-200 days into the outbreak (54% of cumulative cases), but this contribution decreased at later outbreak stages (52% of cumulative cases) (Figure 12B,C). Overall, behavioural configuration had little impact on the percent contribution of each subpopulation to cumulative cases for pandemic influenza and SARS-CoV-2. For Ebola, the 50% accepting, 50% hesitant configuration led to the most even contribution of each subpopulation to cumulative cases by the end of the outbreak (Figure 12C). For the equal split configurations, there was very little variation in the vaccine-hesitant subpopulation contribution to cumulative cases through time, while the magnitudes of contribution to cases in the vaccine-resistant and vaccine-accepting subpopulations were similar to those in the 50% split configurations (Figure 12D).

**Figure 12.**
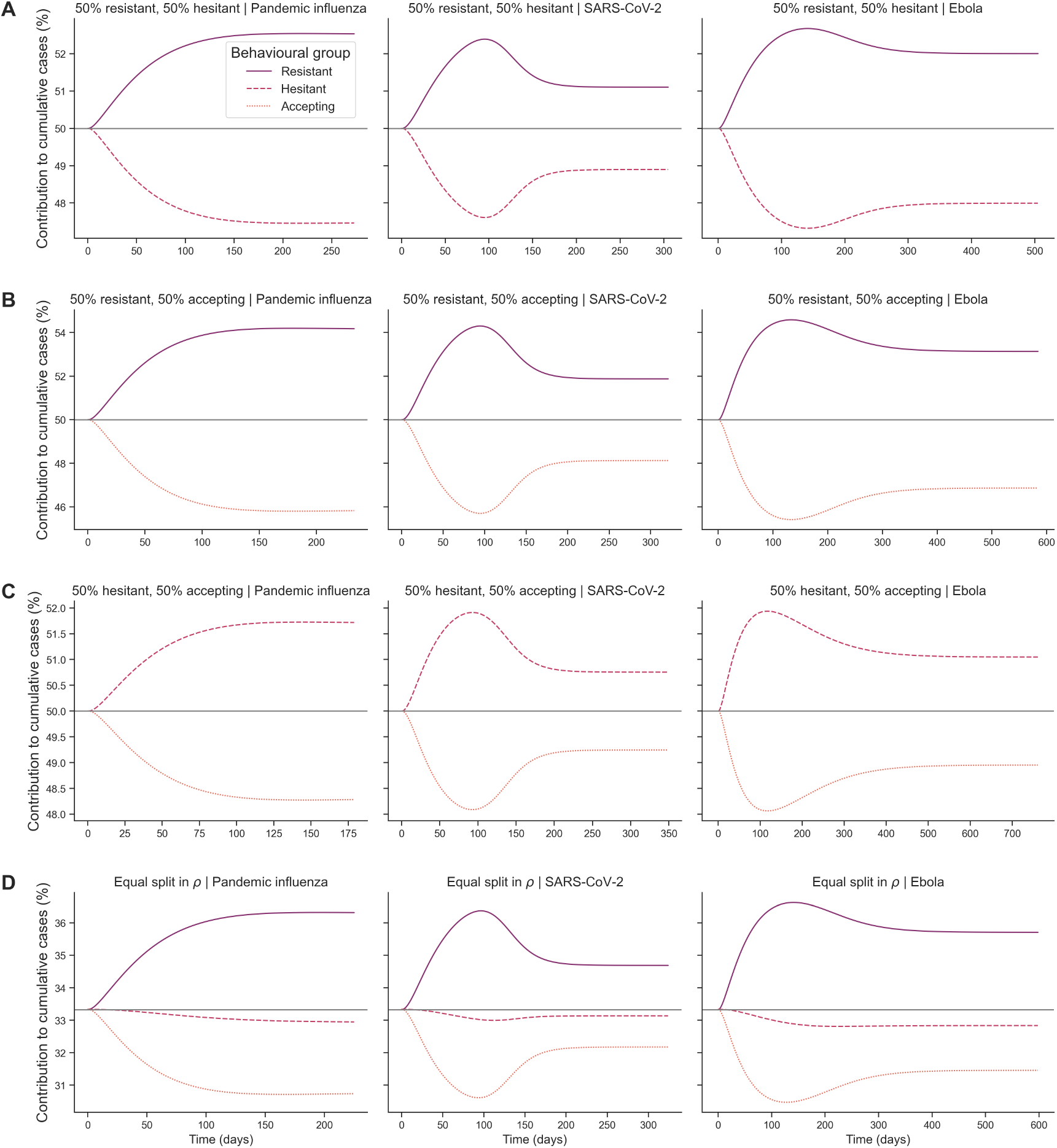
Heterogeneous scenario 2. Subpopulation-level percent contribution to cumulative cases over time across pathogen systems for heterogeneous behavioural configurations and outbreak information based on local cases. Each row shows percent contribution for each subpopulation towards cumulative cases across time in days for a unique behavioural configuration: **(A)** 50% resistant (*ρ* = 0), 50% accepting (*ρ* = 2); **(B)** 50% resistant (*ρ* = 0), 50% hesitant (*ρ* = 1); **(C)** 50% accepting (*ρ* = 2), 50% hesitant (*ρ* = 1); and **(D)** equal split in vaccine opinion (*ρ*∈ 0, 1, 2). Each column represents a different pathogen system: pandemic influenza (first column), SARS-CoV-2 (second column) and Ebola (third column). For each behavioural configuration, lines are shown for each behavioural subpopulation: vaccine-resistant (*ρ* = 0), vaccine-hesitant (*ρ* = 1) and vaccine-accepting (*ρ* = 2). The grey line represents the expected contribution of each subpopulation if no differences are present. Information sensitivity (*α*) was fixed at 2, memory window (*µ*) was fixed at the full history and vaccine effectiveness (*ε*) was fixed at 50%.

## Discussion

Mathematical models of infectious disease dynamics can contribute to public health response efforts against infectious disease outbreaks. Nonetheless, heterogeneity in human behaviour has typically not been considered in such models. Despite the numerous studies that explore human behaviour during outbreaks in sociological and psychological contexts, studies that examine how human behaviour affects disease dynamics is a growing area of interest, particularly as a result of the COVID-19 pandemic [19, 28, 58]. By improving existing disease models by considering heterogeneity in human behaviour, modellers can therefore help better inform public health officials and policymakers [19, 20, 46].

To contribute to this existing literature, we have presented a SEIR-type disease model that incorporates a subpopulation-level intervention adherence behavioural function modifier. The behavioural modifier considered an initial intervention opinion, real-time response to local and global outbreak information - in our case capturing a reduction in vaccine uptake as a result of imperfect protection resulting in breakthrough infections and deaths in vaccinated individuals - and sensitivity to such outbreak information. Using vaccination as an example intervention application and three different pathogens of public health concern (pandemic influenza, SARS-CoV-2 and Ebola), through computational simulation we have shown how subpopulation-level behavioural heterogeneity can result in disparate epidemic impacts on public health. In the current literature, behavioural parameters related to vaccine uptake are not generally explored in mathematical models. We found that these behavioural elements are necessary to explore in future models of infectious disease, since epidemic outcomes vary drastically under different conditions within our parameter space of interest.

We found that differences in preference towards outbreak information were pathogen-specific. Consequently, in some pathogen systems, outbreak information types at different outbreak stages may be more informative to an information-sensitive population and lead to less severe epidemic outcomes. It has been found that local spread of disease awareness during an outbreak can stop a disease from spreading [47]. Similarly, we found that sensitivity to local outbreak information can lead to mild outbreaks, however, this less-severe outcome is dependent on the pathogen system and the behavioural configuration of the population. As emphasised by Funk *et al*. 2010, information individuals use in decision-making during outbreaks may be based on disease prevalence or information independent of prevalence, such as prior beliefs about vaccination [19]. We have aimed to capture these two types of information in this study by considering subpopulation-level vaccine opinion, information sensitivity and type of outbreak information (i.e. local cases, local deaths).

In this study, outbreak information preference was sensitive to vaccine effectiveness, demonstrating the importance of considering human behaviour during outbreaks in the context of the perceived effectiveness of the intervention. In situations where individuals may perceive low infection risk, their perceived risk of adhering to the intervention may play a more significant role in decision making [48].

During an emerging infectious disease outbreak, it is important to consider how real-time outbreak information is disseminated to the public and the heterogeneity that may exist in data reporting by various sources (e.g. government websites, social media, news media) [21, 49, 50].

We also found that behavioural configuration was important in explaining variations in outbreak severity, drawing attention to the relevance of behavioural heterogeneity when planning in the public health sector. Whilst our study was an exploratory modelling investigation into how epidemiological impacts depended on vaccine beliefs and sensitivity to outbreak information amongst the population, prospectively the behavioural elements of the model can be parameterised from behavioural data. Information can be garnered from historical outbreaks, such as the 2018-2019 Ebola outbreak in DR Congo where the spread of misinformation has been studied [31]. Unifying the novel components of our model structure with this model of misinformation spread would be an interesting direction to explore, for instance. We also recommend reflecting on the plausibility of applying such models as part of real-time response efforts, which will require timely availability of relevant data on behavioural characteristics. There may be groups of people underrepresented in a given data set given limitations in sampling and data privacy regulations, requiring further validation [16]. We therefore encourage reflection on the forms of data collected during the COVID-19 pandemic and development of appropriate data management procedures to assist data availability.

In psychology, Protection Motivation Theory (PMT), the Extended Parallel Processing Model (EPPM) and the COM-B model provide valuable frameworks for understanding how individuals process outbreak information and make behavioural decisions. PMT suggests that behavioural responses are shaped by threat appraisal (perceived severity and vulnerability) and coping appraisal (perceived efficacy and response costs) [51]. Considering this framework, our model represented threat appraisal through people’s sensitivity to different types of outbreak information appears to be influenced by pathogen characteristics (e.g., susceptibility and mortality). Additionally, coping appraisal is represented in our model through the daily vaccination rate resulting from the underlying behavioural traits. Similarly, the EPPM suggests that individuals respond to fear-based messaging based on their perceived efficacy to act [52]. If both threat perception (how severe and personally relevant the risk is) and efficacy perception (confidence in one’s ability to respond effectively) are high, fear appeals can promote protective behaviours. However, if threat perception is high but efficacy perception is low, people may enter fear control, where they avoid the threat rather than taking action (e.g., denial, fatalism, or disengagement). Although we considered threat perception in our model through outbreak information and epidemiological parameters, it would be interesting to include efficacy perception in further studies. Lastly, the COM-B model is a behaviour change framework that proposes three necessary components for any behaviour to occur: capability, opportunity and motivation [**?**]. In our model, motivation (e.g., beliefs, habits) is represented by vaccine opinion and information sensitivity. Although we do not explicitly include capability (e.g., knowledge, ability) or opportunity (e.g., support, resources), these theory components could be included to examine differences in capability and resource-limited scenarios. The integration of these theoretical perspectives strengthens the interdisciplinary relevance of our model by linking epidemiological and behavioural factors to established psychological mechanisms. Future research could explore these links further, using experimental approaches to assess how threat and coping appraisals influence information processing and behavioural intentions in the context of emerging infectious diseases.

In addition, although we aimed to capture many aspects of disease spread and behaviour, the following points are limitations of the model framework and addressing these are future avenues of research: (i) we considered one form of behavioural function and outbreak information input; (ii) subpopulation-level behavioural traits were representative of social groups and subpopulation sizes were fixed during the simulated outbreak; (iii) we did not include spatial heterogeneity, demographic processes or age-structure; and (iv) vaccination impacted susceptibility to infection and disease severity, had no capacity constraints and was a standalone intervention.

First, we recommend expanding upon the behavioural function, *ν*_*i*_ (*t*), by incorporating additional variables or different function types. Additional variables could include differentiating between individual cost of infection and cost of adherence. The outbreak information can also be generalised to account for different epidemiological metrics, such as a dependency on disease incidence and/or prevalence. It is also the case that opinions of individuals on interventions can change over the course of infectious disease outbreaks due to new scientific findings, government regulations or changes in perceived risk [53]. One possibility for incorporating such temporal dependencies is the use of objective and cost functions to demonstrate individual-based or government-based choices over time (a commonly used strategy in opinion dynamics) [20, 54].

Second, we assumed that subpopulation sizes were fixed during the simulated outbreaks, as opposed to allowing movement between subpopulations during the outbreak, and that the three vaccine opinion groups (resistant, hesitant, accepting) were representative of social groups. Allowing for individuals to change their preexisting beliefs on the intervention strategy based on dynamics such as conformity and new information presented in the media would be a reasonable direction to explore. When considering local outbreak information, the vaccine uptake rate for a given subpopulation accounted for outbreak information within the subpopulation alone. By incorporating local and global information concurrently, we anticipate more variable outcomes in outbreak severity.

Thirdly, the model did not account for spatial heterogeneity, demographic processes or age-structure. Vaccine-related behaviour, and infection intervention-related behaviour mode generally, can be highly correlated with spatial location, especially in social networks with few close contacts or in areas with a high representation of susceptible groups. Spatial variations in behavioural traits would be important when considering long-distance dispersal of pathogens [55, 56] and for disease outbreaks occurring in active conflict zones that impact the ability to enact infection control strategies [36]. We chose not to include demographic processes given the relatively short time scales (a few years) of the simulated outbreaks. If considering the possibility of multiple variants of a pathogen and waning immunity, then due consideration should be given to population-level processes that can alter the immunity structure. With regard to age-structure, the pathogens that we considered tend to have disproportionate susceptibility to severe infection in young children, the elderly and individuals with preexisting medical conditions. Given the similarity across the three pathogens of interest, we anticipate the inclusion of age-structure would not qualitatively alter our findings.

Lastly, we assumed that vaccination was infection blocking and reduced disease severity, had no capacity constraints and was a standalone intervention. We chose not to include reducing transmissibility as an action of the vaccine to simplify the model. However, this vaccine action could be explored in a future extension of the model. In response to the COVID-19 pandemic it has become more common for vaccination modelling studies to consider the multiple protective actions of vaccination; for example, a study by Keeling *et al*. [3] partitioned the action of a COVID-19 vaccine into five elements: protection against infection; protection against symptomatic disease; protection against requiring hospital admission; protection against death; and the reduction in onward transmission for vaccinated individuals who do become infected. We also made the simplifying assumption that resources were always available regardless of vaccine uptake, but could consider intervention availability to demonstrate situations with limited public health and hospital resources [57]. Exploring human behaviour in the context of varied resources available during outbreaks would be essential to investigate likely differences in disease and behaviour dynamics in resource-limited scenarios. Vaccination also served as a sole intervention, in order to focus on the epidemiological impacts of vaccine beliefs and sensitivity to outbreak information within subgroups. For the purposes of this work, we considered vaccination as the intervention strategy used to combat disease spread. However, it is important to recognise that other intervention strategies are often used in conjunction such as social distancing and mask-usage in various geographic and social contexts [21, 58]. For instance, antivirals have been used to combat pandemic influenza [59]. For SARS-CoV-2, non-pharmaceutical interventions such as self-quarantining or disinfecting frequently-used surfaces were used [4]. Ebola is transmitted differently than the previous two pathogens, which have made safe burials, contact tracing and case management reasonable measures to combat disease [36]. The inclusion of such intervention strategies in conjunction with vaccination would provide further insights into pathogen-specific preferences towards outbreak information in resource-limited scenarios.

In conclusion, this work contributes to the existing literature by encapsulating awareness of population and subpopulation-level intervention effectiveness in real-time within a human infectious disease model. We demonstrated the need for exploring different behavioural functions, with variability exhibited in epidemic impacts given different behavioural assumptions. Public health officials should consider expanding upon current data collections to include behavioural insights into individual opinions on vaccination (vaccine opinion) and propensity to vaccinate given new information from the government or social circles (outbreak information sensitivity). Our findings suggest that sensitivity to outbreak information is influenced by pathogen characteristics, particularly transmissibility (*R*_0_) and severity (fatality rate). For an emerging infectious disease, these characteristics may not be immediately known with precision, but early estimates could provide a basis for forecasting public response. One possible approach is to use initial epidemiological parameters - such as estimated *R*_0_ and infection fatality rate - to anticipate whether case-based or death-based information is likely to be more effective in communicating risk, although these relationships will likely evolve as more information is gathered across pathogen systems. Additionally, real-time public sentiment analysis (e.g. through surveys or social media monitoring) could help refine this understanding as more data become available. With these data available, disease modellers can use such data within model frameworks such as the one we have presented in this study, enabling us to better understand behavioural implications on epidemic outcomes. We encourage researchers to continue enhancing the body of work in the behavioural epidemiology field, which will be integral in combating future infectious disease outbreaks.

## Acknowledgements

RLS and MJT were supported by the Engineering and Physical Sciences Research Council through the MathSys CDT (grant number EP/S022244/1). MJT and EMH are linked with the JUNIPER partnership (MRC grant no MR/X018598/1) and would like to acknowledge their help and support. EMH is affiliated to the National Institute for Health and Care Research Health Protection Research Unit (NIHR HPRU) in Gastrointestinal Infections at University of Liverpool (PB-PG-NIHR-200910), in partnership with the UK Health Security Agency (UKHSA), in collaboration with the University of Warwick (EMH is based at The University of Liverpool). The views expressed are those of the author(s) and not necessarily those of the NIHR, the Department of Health and Social Care or the UK Health Security Agency. The research was funded by The Pandemic Institute, formed of seven founding partners: The University of Liverpool, Liverpool School of Tropical Medicine, Liverpool John Moores University, Liverpool City Council, Liverpool City Region Combined Authority, Liverpool University Hospital Foundation Trust, and Knowledge Quarter Liverpool (EMH is based at The University of Liverpool). The views expressed are those of the author(s) and not necessarily those of The Pandemic Institute. For the purpose of open access, the authors have applied a Creative Commons Attribution (CC BY) licence to any Author Accepted Manuscript version arising from this submission.

## Author contributions statement

**Rachel L. Seibel:** Data curation, Formal analysis, Investigation, Methodology, Software, Validation, Visualisation, Writing - Original Draft, Writing - Review & Editing.

**Michael J. Tildesley:** Conceptualisation, Methodology, Software, Supervision, Visualisation, Writing - Original Draft, Writing

- Review & Editing.

**Edward M. Hill:** Conceptualisation, Methodology, Software, Supervision, Visualisation, Writing - Original Draft, Writing - Review & Editing.

## Additional information

## Data availability

All data utilised in this study are publicly available, with relevant references and data repositories provided.

## Code availability

The code repository for the study is available at: https://github.com/rachelseibel/outbreak_information_model

Archived code associated with this version of the study: https://doi.org/10.5281/zenodo.15024878

## References

1. Klepac P, Kucharski AJ, Conlan AJK, Kissler S, Tang M, Fry H, Gog JR. 2020 Contacts in context: Large-scale setting-specific social mixing matrices from the BBC Pandemic project. medRxiv. (10.1101/2020.02.16.20023754)

2. Gimma A, Munday JD, Wong KLM, Coletti P, van Zandvoort K, Prem K, Klepac P, Rubin GJ, Funk S, Edmunds WJ, Jarvis CI. 2022 Changes in social contacts in England during the COVID-19 pandemic between March 2020 and March 2021 as measured by the CoMix survey: A repeated cross-sectional study. PLOS Medicine 19, e1003907. (10.1371/journal.pmed.1003907)

3. Keeling MJ, Dyson L, Tildesley MJ, Hill EM, Moore S. 2022 Comparison of the 2021 COVID-19 roadmap projections against public health data in England. Nature communications 13, 4924. (10.1038/s41467-022-31991-0)

4. West R, Michie S, Rubin GJ, Amlôt R. 2020 Applying principles of behaviour change to reduce SARS-CoV-2 transmission. Nature human behaviour 4, 451–459.

5. Rocha YM, Moura GAD, Desidério GA, Oliveira CHD, Lourenço FD, Deadame L, Nicolete F. 2021 The impact of fake news on social media and its influence on health during the COVID-19 pandemic: A systematic review. Journal of Public Health: From Theory to Practice 31. (10.1007/s10389-021-01658-z/Published)

6. Nowak BM, Miedziarek C, Pełczynśki S, Rzymski P. 2021 Misinformation, fears and adherence to preventive measures during the early phase of COVID-19 pandemic: A cross-sectional study in Poland. International Journal of Environmental Research and Public Health 18. (10.3390/ijerph182212266)

7. Ferguson N. 2007 Capturing human behaviour. Nature 446, 733. (10.1038/446733a)

8. Kok G, Jonkers R, Gelissen R, Meertens R, Schaalma H, de Zwart O. 2010 Behavioural intentions in response to an influenza pandemic. BMC Public Health 10, 174. (10.1186/1471-2458-10-174)

9. Balinska M, Rizzo C. 2009 Behavioural responses to influenza pandemics. PLOS Currents 1, RRN1037. (10.1371/currents.RRN1037)

10. Van D, McLaws ML, Crimmins J, MacIntyre CR, Seale H. 2010 University life and pandemic influenza: Attitudes and intended behaviour of staff and students towards pandemic (H1N1) 2009. BMC Public Health 10, 130. (10.1186/1471-2458-10-130)

11. Vinck P, Pham PN, Bindu KK, Bedford J, Nilles EJ. 2019 Institutional trust and misinformation in the response to the 2018–19 Ebola outbreak in North Kivu, DR Congo: A population-based survey. The Lancet Infectious Diseases 19, 529–536. (10.1016/S1473-3099(19)30063-5)

12. Brooks-Pollock E, Northstone K, Pellis L, Scarabel F, Thomas A, Nixon E, Matthews DA, Bowyer V, Garcia MP, Steves CJ et al. 2023 Voluntary risk mitigation behaviour can reduce impact of SARS-CoV-2: a real-time modelling study of the January 2022 Omicron wave in England. BMC medicine 21, 25. (10.1186/s12916-022-02714-5)

13. Verelst F, Willem L, Beutels P. 2016 Behavioural change models for infectious disease transmission: a systematic review (2010–2015). Journal of The Royal Society Interface 13, 20160820. (10.1098/rsif.2016.0820)

14. Jackson T, Steed L, Pedruzzi R, Beyene K, Chan AHY. 2022 Editorial: COVID-19 and behavioral sciences. Frontiers in Public Health 9. (10.3389/fpubh.2021.830797)

15. Liu Y, Wu B. 2022 Coevolution of vaccination behavior and perceived vaccination risk can lead to a stag-hunt-like game. Physical Review E 106, 034308. (10.1103/PhysRevE.106.034308)

16. Buckee C, Noor A, Sattenspiel L. 2021 Thinking clearly about social aspects of infectious disease transmission. Nature 595, 205–213. (10.1038/s41586-021-03694-x)

17. Peralta AF, Kertész J, Iñiguez G. 2022 Opinion dynamics in social networks: From models to data. arXiv p. 2201.01322. (10.48550/arXiv.2201.01322)

18. Michie S, van Stralen MM, West R. 2011 The behaviour change wheel: A new method for characterising and designing behaviour change interventions. Implementation Science 6, 42. (10.1186/1748-5908-6-42)

19. Funk S, Salathe M, Jansen VAA. 2010 Modelling the influence of human behaviour on the spread of infectious diseases: A review. Journal of The Royal Society Interface 7, 1247–1256. (10.1098/rsif.2010.0142)

20. Shea K, Tildesley MJ, Runge MC, Fonnesbeck CJ, Ferrari MJ. 2014 Adaptive management and the value of information: Learning via intervention in epidemiology. PLOS Biology 12. (10.1371/journal.pbio.1001970)

21. Nita Bharti. 2021 Linking human behaviors and infectious disease. Proceedings of the National Academy of Sciences of the United States of America 118. (10.1073/pnas.2005241118)

22. Smaldino PE, Jones JH. 2021 Coupled dynamics of behaviour and disease contagion among antagonistic groups. Evolutionary Human Sciences 3, e28. (10.1017/ehs.2021.22)

23. Bedson J, Skrip LA, Pedi D, Abramowitz S, Carter S, Jalloh MF, Funk S, Gobat N, Giles-Vernick T, Chowell G, De Almeida JR, Elessawi R, Scarpino SV, Hammond RA, Briand S, Epstein JM, Hébert-Dufresne L, Althouse BM. 2021 A review and agenda for integrated disease models including social and behavioural factors. Nature Human Behaviour 5, 834–846. (10.1038/s41562-021-01136-2)

24. Verelst F, Willem L, Beutels P. 2016 Behavioural change models for infectious disease transmission: A systematic review (2010–2015). Journal of The Royal Society Interface 13, 20160820. (10.1098/rsif.2016.0820)

25. Kiss IZ. 2013 Incorporating human behaviour in epidemic dynamics: A modelling perspective. In Manfredi P, D’Onofrio A, editors, Modeling the Interplay Between Human Behavior and the Spread of Infectious Diseases, pp. 125–137. New York, NY: Springer. (10.1007/978-1-4614-5474-8_8)

26. Weston D, Hauck K, Amlôt R. 2018 Infection prevention behaviour and infectious disease modelling: A review of the literature and recommendations for the future. BMC Public Health 18, 336. (10.1186/s12889-018-5223-1)

27. Haensch A, Dragovic N, Börgers C, Boghosian B. 2022 COVID-19 vaccine hesitancy and mega-influencers. arXiv p. 2202.00630. (10.48550/arXiv.2202.00630)

28. Troiano G, Nardi A. 2021 Vaccine hesitancy in the era of COVID-19. Public Health 194, 245–251. (10.1016/j.puhe.2021.02.025)

29. Robertson E, Reeve KS, Niedzwiedz CL, Moore J, Blake M, Green M, Katikireddi SV, Benzeval MJ. 2021 Predictors of COVID-19 vaccine hesitancy in the UK household longitudinal study. Brain, Behavior, and Immunity 94, 41–50. (10.1016/j.bbi.2021.03.008)

30. Lipsitch M, Krammer F, Regev-Yochay G, Lustig Y, Balicer RD. 2022 SARS-CoV-2 breakthrough infections in vaccinated individuals: measurement, causes and impact. Nature Reviews Immunology 22, 57–65. (10.1038/s41577-021-00662-4)

31. Vinck P, Pham PN, Bindu KK, Bedford J, Nilles EJ. 2019 Institutional trust and misinformation in the response to the 2018–19 Ebola outbreak in North Kivu, DR Congo: A population-based survey. The Lancet Infectious Diseases 19, 529–536. (10.1016/S1473-3099(19)30063-5)

32. Wu N, Joyal-Desmarais K, Ribeiro PAB, Vieira AM, Stojanovic J, Sanuade C, Yip D, Bacon SL. 2023 Long-term effectiveness of COVID-19 vaccines against infections, hospitalisations, and mortality in adults: findings from a rapid living systematic evidence synthesis and meta-analysis up to December, 2022. The Lancet Respiratory Medicine 11, 439–452. (10.1016/S2213-2600(23)00015-2)

33. Pere G, Arantxa R, Núria S, Nuria T, Mireia J, Ana M, A CJ, Cristina R, Angela D, on Surveillance of Severe Influenza Hospitalized Cases in Catalonia. TWG. 2018 Influenza vaccine effectiveness in reducing severe outcomes over six influenza seasons, a case-case analysis, Spain, 2010/11 to 2015/16.. Euro Surveill 23. (10.2807/1560-7917.ES.2018.23.43.1700732)

34. Biggerstaff M, Cauchemez S, Reed C, Gambhir M, Finelli L. 2014 Estimates of the reproduction number for seasonal, pandemic, and zoonotic influenza: A systematic review of the literature. BMC Infectious Diseases 14. (10.1186/1471-2334-14-480)

35. D’Arienzo M, Coniglio A. 2020 Assessment of the SARS-CoV-2 basic reproduction number, R0, based on the early phase of COVID-19 outbreak in Italy. Biosafety and Health 2, 57–59. (10.1016/j.bsheal.2020.03.004)

36. Wong ZSY, Bui CM, Chughtai AA, Macintyre CR. 2017 A systematic review of early modelling studies of Ebola virus disease in West Africa. Epidemiology and Infection 145, 1069–1094. (10.1017/S0950268817000164)

37. Edlund S, Kaufman J, Lessler J, Douglas J, Bromberg M, Kaufman Z, Bassal R, Chodick G, Marom R, Shalev V, Mesika Y, Ram R, Leventhal A. 2011 Comparing three basic models for seasonal influenza. Epidemics 3, 135–142. (10.1016/j.epidem.2011.04.002)

38. Johansson MA, Quandelacy TM, Kada S, Prasad PV, Steele M, Brooks JT, Slayton RB, Biggerstaff M, Butler JC. 2021 SARS-CoV-2 transmission from people without COVID-19 symptoms. JAMA Network Open 4. (10.1001/jamanetworkopen.2020.35057)

39. Wong JY, Wu P, Nishiura H, Goldstein E, Lau EHY, Yang L, Chuang SK, Tsang T, Peiris JSM, Wu JT, Cowling BJ. 2013 Infection fatality risk of the pandemic A(H1N1)2009 virus in Hong Kong. American Journal of Epidemiology 177, 834–840. (10.1093/aje/kws314)

40. COVID-19 Forecasting Team. 2022 Variation in the COVID-19 infection–fatality ratio by age, time, and geography during the pre-vaccine era: A systematic analysis. The Lancet 399, 1469–1488. (10.1016/S0140-6736(21)02867-1)

41. Althaus CL, Low N, Musa EO, Shuaib F, Gsteiger S. 2015 Ebola virus disease outbreak in Nigeria: Transmission dynamics and rapid control. Epidemics 11, 80–84.

42. Campbell A, Rodin R, Kropp R, Mao Y, Hong Z, Vachon J, Spika J, Pelletier L. 2010 Risk of severe outcomes among patients admitted to hospital with pandemic (H1N1) influenza. Canadian Medical Association Journal 182.

43. Faes C, Abrams S, Van Beckhoven D, Meyfroidt G, Vlieghe E, Hens N, on COVID-19 Hospital Surveillance BCG. 2020 Time between symptom onset, hospitalisation and recovery or death: Statistical analysis of Belgian COVID-19 patients. International Journal of Environmental Research and Public Health 17. (10.3390/ijerph17207560)

44. Ji YJ, Duan XZ, Gao XD, Li L, Li C, Ji D, Li WG, Wang LF, Meng YH, Yang X, Ling BF, Gu ML, Jiang T, Koroma SKM, Bangalie J, Duan HJ. 2016 Clinical presentations and outcomes of patients with Ebola virus disease in Freetown, Sierra Leone. Infectious Diseases of Poverty 5.

45. Mathieu E, Ritchie H, Ortiz-Ospina E, Roser M, Hasell J, Appel C, Giattino C, Rodés-Guirao L. 2021 A global database of COVID-19 vaccinations. Nature Human Behaviour 5, 947–953. (10.1038/s41562-021-01122-8)

46. Funk S, Bansal S, Bauch CT, Eames KT, Edmunds WJ, Galvani AP, Klepac P. 2015 Nine challenges in incorporating the dynamics of behaviour in infectious diseases models. Epidemics 10, 21–25. (10.1016/j.epidem.2014.09.005)

47. Funk S, Gilad E, Watkins C, Jansen VAA. 2009 The spread of awareness and its impact on epidemic outbreaks. Proceedings of the National Academy of Sciences 106, 6872–6877. (10.1073/pnas.0810762106)

48. Manfredi P, D’Onofrio A, editors. 2013 Modeling the Interplay Between Human Behavior and the Spread of Infectious Diseases. New York, NY: Springer New York. (10.1007/978-1-4614-5474-8)

49. Lawson B, Lugo-Ocando J. 2022 Political communication, press coverage and public interpretation of public health statistics during the coronavirus pandemic in the UK. European Journal of Communication 37, 646–662. (10.1177/02673231221099407)

50. Polonsky JA, Baidjoe A, Kamvar ZN, Cori A, Durski K, John Edmunds W, Eggo RM, Funk S, Kaiser L, Keating P, Le Polain De Waroux O, Marks M, Moraga P, Morgan O, Nouvellet P, Ratnayake R, Roberts CH, Whitworth J, Jombart T. 2019 Outbreak analytics: A developing data science for informing the response to emerging pathogens. Philosophical Transactions of the Royal Society B: Biological Sciences 374. (10.1098/rstb.2018.0276)

51. Rogers RW. 1983 Cognitive and physiological processes in fear appeals and attitude change: a revised theory of protection motivation. In Social Psychophysiology,. New York, NY: Guilford Press.

52. Witte K. 1992 Putting the fear back into fear appeals: The extended parallel process model. Communication Monographs 59, 329–349. (10.1080/03637759209376276)

53. Wajid S, Samreen S, Sales I, Bawazeer G, Mahmoud MA, Aljohani MA. 2022 What has changed in the behaviors of the public after the COVID-19 pandemic? A cross-sectional study from the Saudi community perspective. Frontiers in Public Health 10, 723229. (10.3389/fpubh.2022.723229)

54. Probert WJ, Shea K, Fonnesbeck CJ, Runge MC, Carpenter TE, Dürr S, Garner MG, Harvey N, Stevenson MA, Webb CT, Werkman M, Tildesley MJ, Ferrari MJ. 2016 Decision-making for foot-and-mouth disease control: Objectives matter. Epidemics 15, 10–19. (10.1016/j.epidem.2015.11.002)

55. Keeling MJ, Woolhouse MEJ, Shaw DJ, Matthews L, Chase-Topping M, Haydon DT, Cornell SJ, Kappey J, Wilesmith J, Grenfell BT. 2001 Dynamics of the 2001 UK foot and mouth epidemic: Stochastic dispersal in a heterogeneous landscape. Science 294, 813–817. (10.1126/science.1065973)

56. Severns PM, Sackett KE, Farber DH, Mundt CC. 2019 Consequences of long-distance dispersal for epidemic spread: Patterns, scaling, and mitigation. Plant Disease 103, 177–191. (10.1094/PDIS-03-18-0505-FE)

57. Coelho FC, Codeço CT. 2009 Dynamic modeling of vaccinating behavior as a function of individual beliefs. PLOS Computational Biology 5. (10.1371/journal.pcbi.1000425)

58. de Bruin M, Suk JE, Baggio M, Blomquist SE, Falcon M, Forjaz MJ, Godoy-Ramirez K, Leurs M, Rodriguez-Blazquez C, Romay-Barja M, Uiters E, Kinsman J. 2022 Behavioural insights and the evolving COVID-19 pandemic. Eurosurveillance 27, pii=2100615. (10.2807/1560-7917.ES.2022.27.18.2100615)

59. Donaldson LJ, Rutter PD, Ellis BM, Greaves FEC, Mytton OT, Pebody RG, Yardley IE. 2009 Mortality from pandemic A/H1N1 2009 influenza in England: Public health surveillance study. BMJ 339, b5213–b5213. (10.1136/bmj.b5213)

