## Supplementary material for "Unifying human infectious disease models and real-time awareness of population- and subpopulation-level intervention effectiveness": Seibel_etal_2025_SI.pdf

### S1 Additional homogeneous population scenario results

#### S1.1 Homogeneous scenario 1

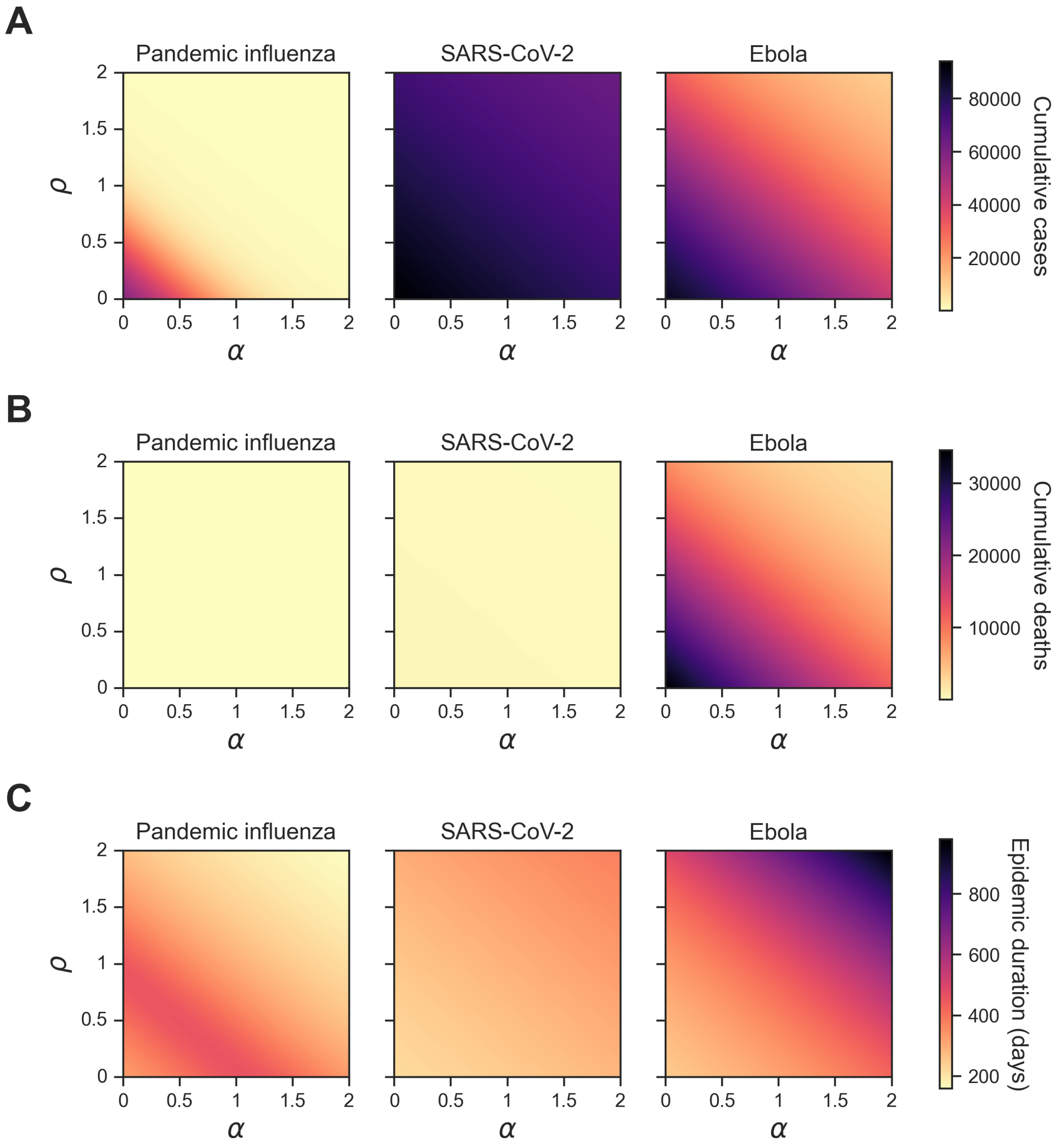

**Figure S1. Homogeneous scenario 1. Summary epidemiological metrics by pathogen when outbreak information was based on global deaths ($\boldsymbol{\theta}_{GD}$).** Each panel shows a given epidemiological statistic across different levels of information sensitivity ($\alpha$) (x-axis, ranging from 0 to 2) and different levels of vaccine opinion ($\rho$) (y-axis, ranging from 0 to 2). Each column corresponds to a different respiratory pathogen: pandemic influenza (column one), SARS-CoV-2 (column two) and Ebola (column three). Each row displays one of the three summary epidemiological measures with a shared colour bar: **(A)** cumulative cases; **(B)** cumulative deaths; **(C)** epidemic duration (days). Darker shading corresponds to higher values for each epidemiological metric.

#### S1.2 Homogeneous scenario 2

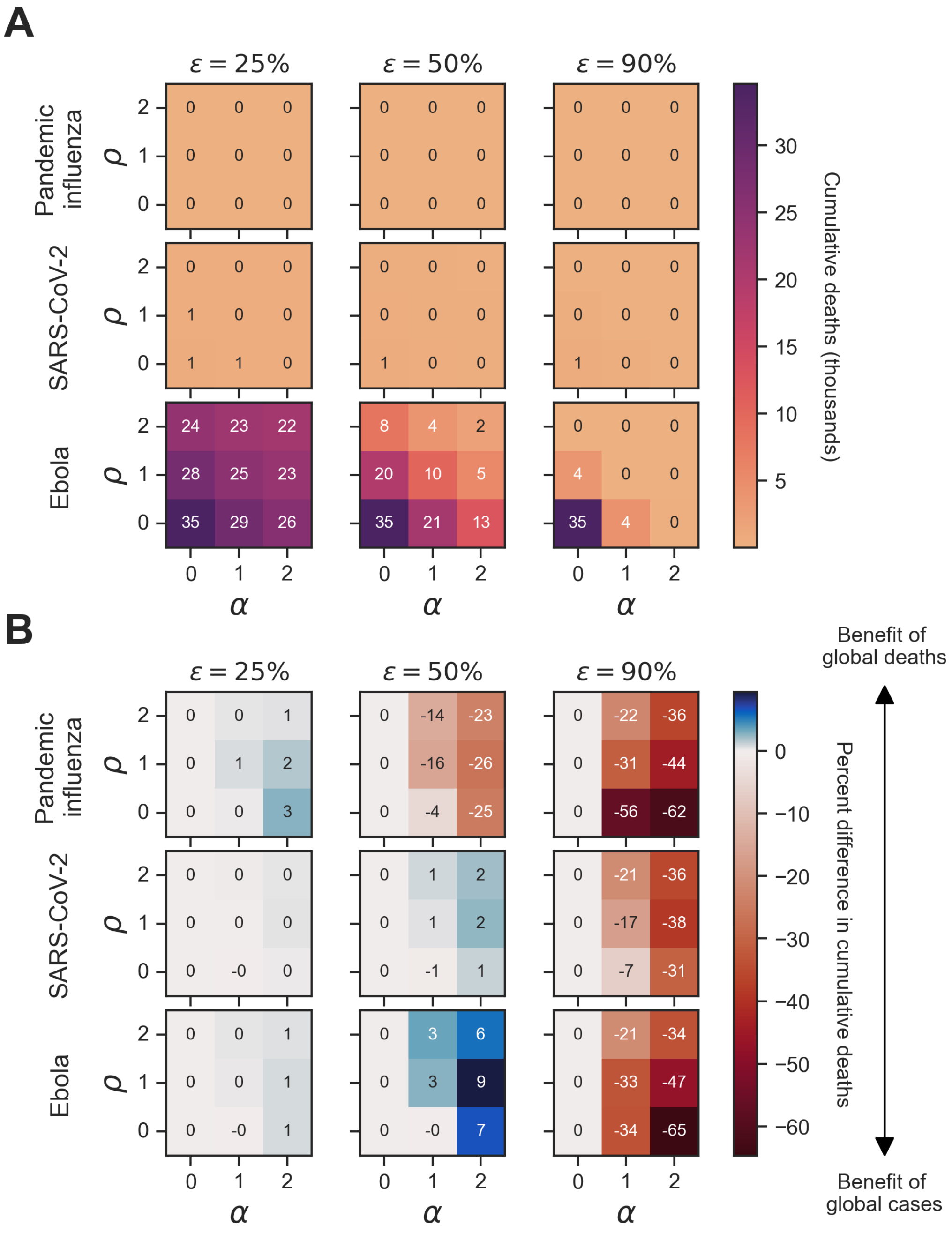

**Figure S2. Homogeneous scenario 2. Sensitivity of cumulative deaths to vaccine efficacy across pathogens and type of outbreak information for a full history memory window.** We present cumulative deaths across the three pathogens of interest (pandemic influenza, SARS-CoV-2, Ebola) alongside three vaccine efficacies ($\varepsilon \in 25\%, 50\%, 90\%$) of interest. Panel **(A)** shows simulations where the outbreak information was global cases as well as the corresponding cumulative deaths for each unique combination of pathogen, vaccine efficacy ($\varepsilon$), vaccine opinion ($\rho$) and information sensitivity ($\alpha$). Dark purple hues correspond to more cumulative cases whilst light orange hues correspond to fewer cumulative deaths. Panel **(B)** shows the percent difference in cumulative deaths between simulations where the outbreak information was global cases ($\theta_{GC}$) and global deaths ($\theta_{GD}$) for each unique combination of pathogen, vaccine efficacy ($\varepsilon$), vaccine opinion ($\rho$) and information sensitivity ($\alpha$). Blue hues correspond to positive percent differences in cumulative deaths (representing a benefit of global deaths) whilst red hues correspond to negative percent differences in cumulative deaths (representing a benefit of global cases).

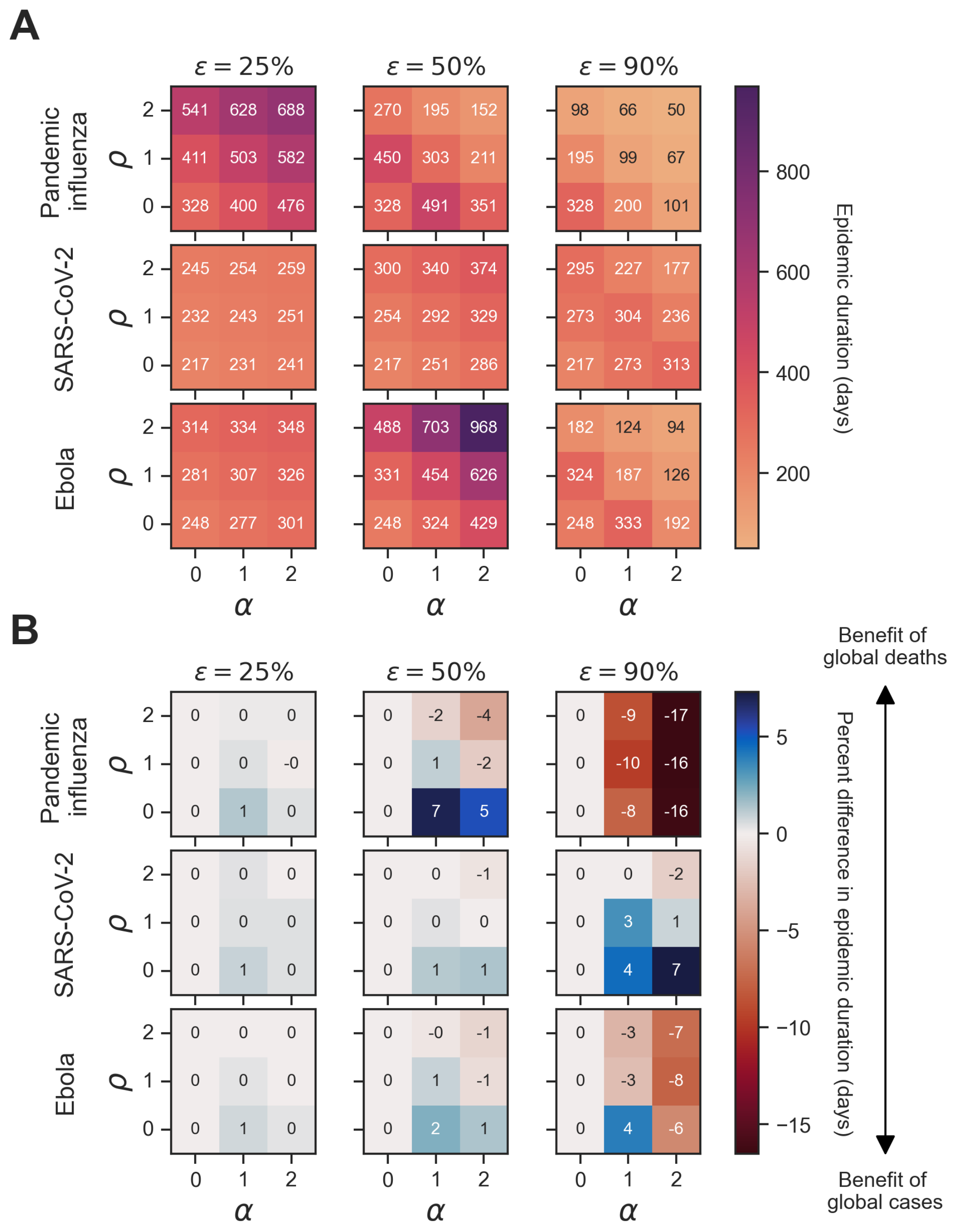

**Figure S3. Homogeneous scenario 2. Sensitivity of epidemic duration in days to vaccine efficacy across pathogens and type of outbreak information for a full history memory window.** For each panel, epidemic durations are presented across the three pathogens of interest (pandemic influenza, SARS-CoV-2, Ebola) alongside three vaccine efficacies ($\varepsilon \in 25\%, 50\%, 90\%$) of interest. Panel **(A)** shows simulations where the outbreak information was global cases as well as the corresponding epidemic duration for each unique combination of pathogen, vaccine efficacy ($\varepsilon$), vaccine opinion ($\rho$) and information sensitivity ($\alpha$). Dark purple hues correspond to longer epidemic durations whilst light orange hues correspond to shorter epidemic durations. Panel **(B)** shows the percent difference in epidemic duration between simulations where the outbreak information was global cases ($\theta_{GC}$) and global deaths ($\theta_{GD}$) for each unique combination of pathogen, vaccine efficacy ($\varepsilon$), vaccine opinion ($\rho$) and information sensitivity ($\alpha$). Blue hues correspond to positive percent differences in epidemic duration (representing a benefit of global deaths) whilst red hues correspond to negative percent differences in epidemic duration (representing a benefit of global cases).

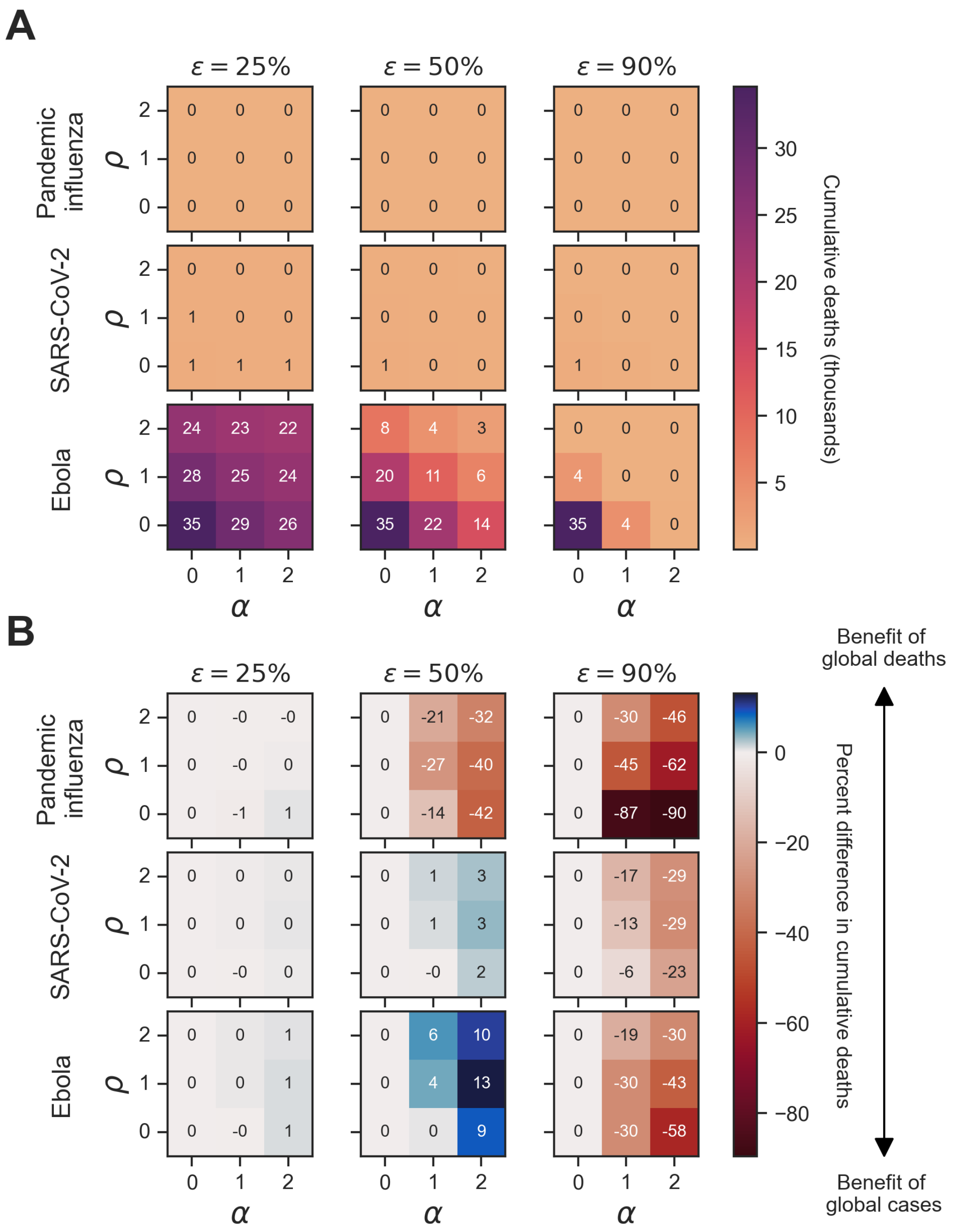

**Figure S4. Homogeneous scenario 2. Sensitivity of cumulative deaths to vaccine efficacy across pathogens and type of outbreak information for a 1-day memory window.** For each panel, cumulative deaths are presented across the three pathogens of interest (pandemic influenza, SARS-CoV-2, Ebola) alongside three vaccine efficacies ($\varepsilon \in 25\%, 50\%, 90\%$) of interest. Panel **(A)** shows simulations where the outbreak information was global cases as well as the corresponding cumulative deaths for each unique combination of pathogen, vaccine efficacy ($\varepsilon$), vaccine opinion ($\rho$) and information sensitivity ($\alpha$). Dark purple hues correspond to more cumulative deaths whilst light orange hues correspond to fewer cumulative deaths. Panel **(B)** shows the percent difference in cumulative deaths between simulations where the outbreak information was global cases ($\theta_{GC}$) and global deaths ($\theta_{GD}$) for each unique combination of pathogen, vaccine efficacy ($\varepsilon$), vaccine opinion ($\rho$) and information sensitivity ($\alpha$). Blue hues correspond to positive percent differences in cumulative deaths (representing a benefit of global deaths) whilst red hues correspond to negative percent differences in cumulative deaths (representing a benefit of global cases).

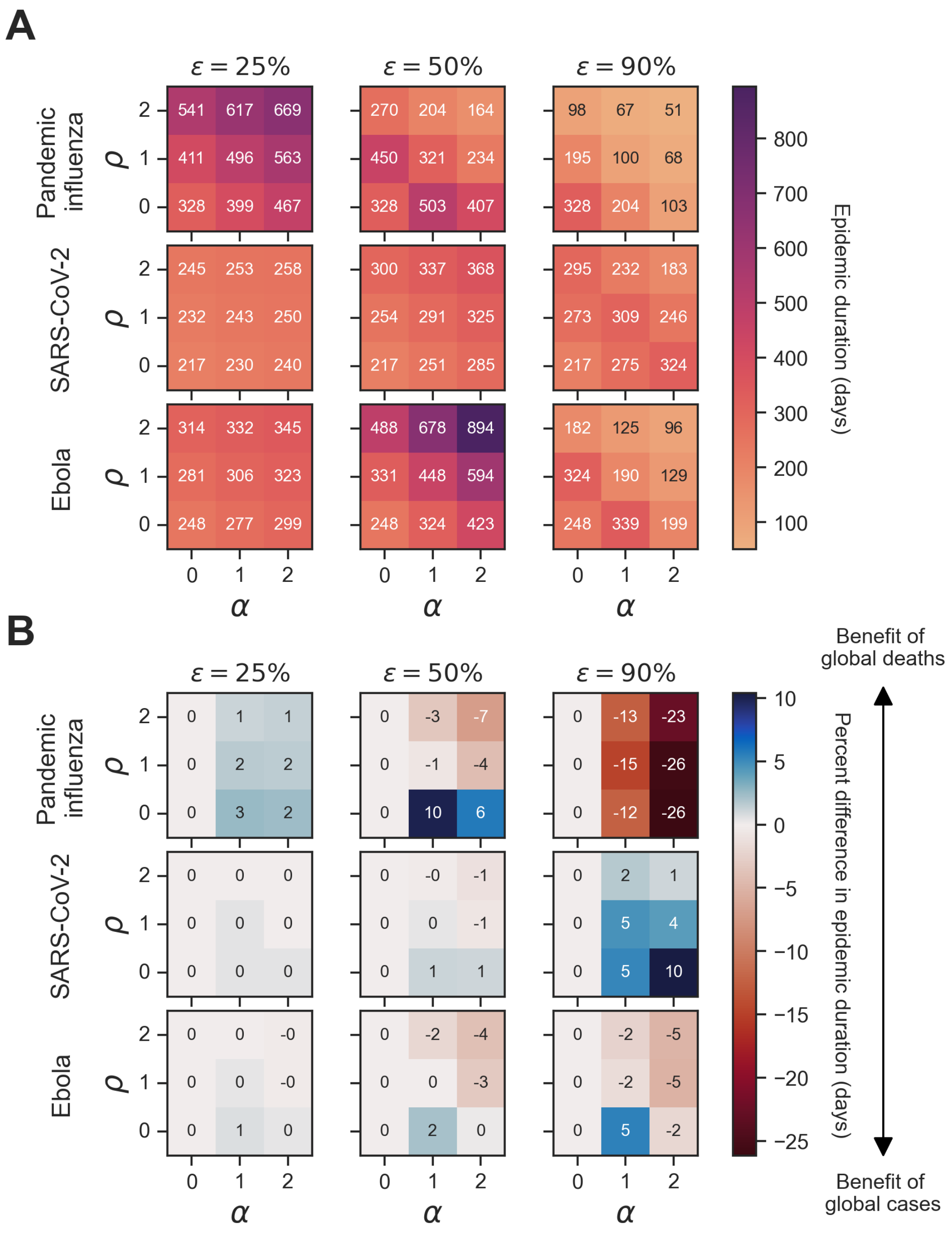

**Figure S5. Homogeneous scenario 2. Sensitivity of epidemic duration in days to vaccine efficacy across pathogens and type of outbreak information for a 1-day memory window.** For each panel, epidemic durations are presented across the three pathogens of interest (pandemic influenza, SARS-CoV-2, Ebola) alongside three vaccine efficacies ($\varepsilon \in 25\%, 50\%, 90\%$) of interest. Panel **(A)** shows simulations where the outbreak information was global cases as well as the corresponding epidemic duration for each unique combination of pathogen, vaccine efficacy ($\varepsilon$), vaccine opinion ($\rho$) and information sensitivity ($\alpha$). Dark purple hues correspond to longer epidemic durations whilst light orange hues correspond to shorter epidemic durations. Panel **(B)** shows the percent difference in epidemic duration between simulations where the outbreak information was global cases ($\theta_{GC}$) and global deaths ($\theta_{GD}$) for each unique combination of pathogen, vaccine efficacy ($\varepsilon$), vaccine opinion ($\rho$) and information sensitivity ($\alpha$). Blue hues correspond to positive percent differences in epidemic duration (representing a benefit of global deaths) whilst red hues correspond to negative percent differences in epidemic duration (representing a benefit of global cases).

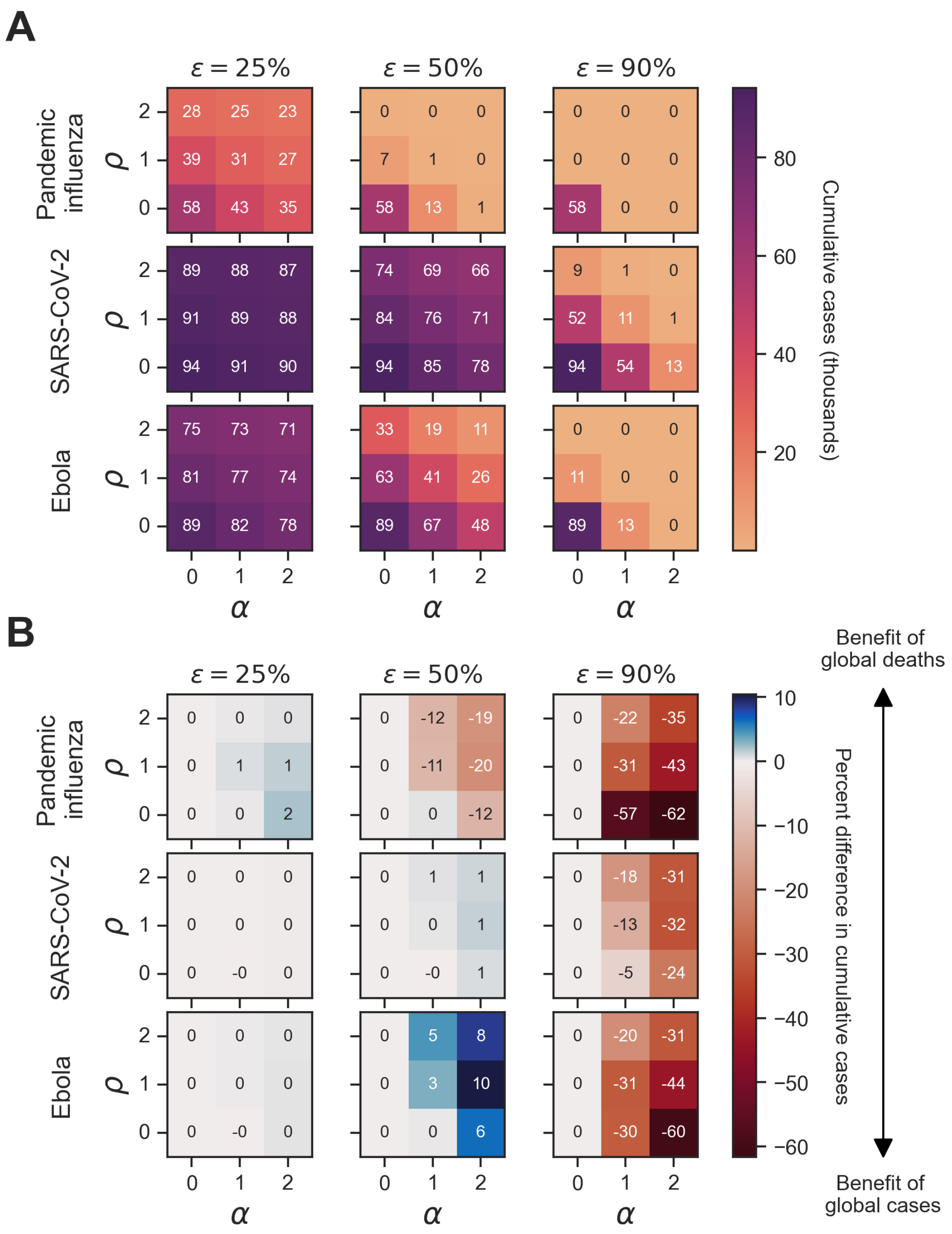

**Figure S6. Homogeneous scenario 2. Sensitivity of cumulative deaths to vaccine efficacy across pathogens and type of outbreak information for a 7-day memory window.** We present cumulative deaths across the three pathogens of interest (pandemic influenza, SARS-CoV-2, Ebola) alongside three vaccine efficacies ($\varepsilon \in 25\%, 50\%, 90\%$) of interest. Panel **(A)** shows simulations where the outbreak information was global cases as well as the corresponding cumulative cases for each unique combination of pathogen, vaccine efficacy ($\varepsilon$), vaccine opinion ($\rho$) and information sensitivity ($\alpha$). Dark purple hues correspond to more cumulative cases whilst light orange hues correspond to fewer cumulative deaths. Panel **(B)** shows the percent difference in cumulative cases between simulations where the outbreak information was global cases ($\theta_{GC}$) and global deaths ($\theta_{GD}$) for each unique combination of pathogen, vaccine efficacy ($\varepsilon$), vaccine opinion ($\rho$) and information sensitivity ($\alpha$). Blue hues correspond to positive percent differences in cumulative deaths (representing a benefit of global deaths) whilst red hues correspond to negative percent differences in cumulative deaths (representing a benefit of global cases).

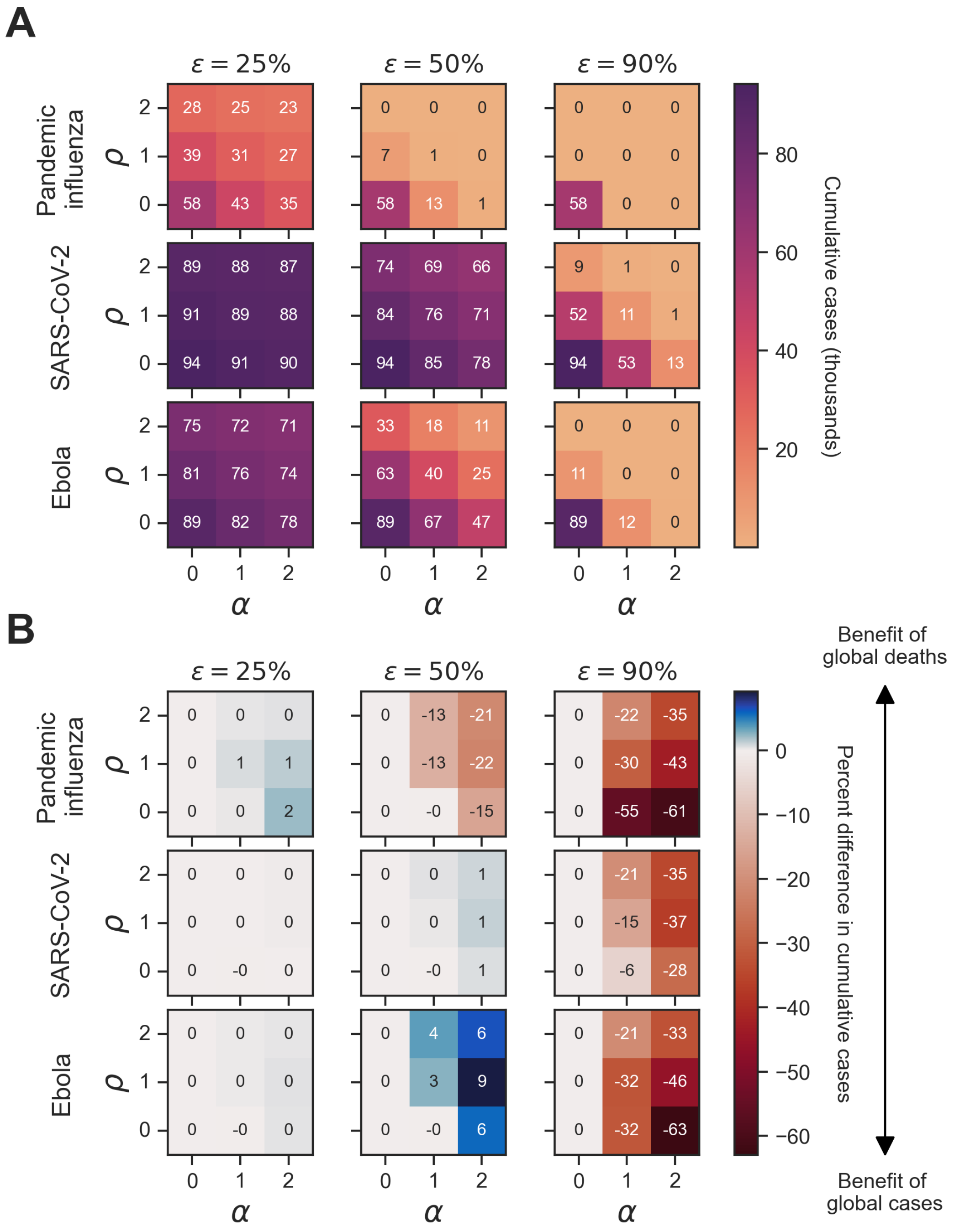

**Figure S7. Homogeneous scenario 2. Sensitivity of cumulative deaths to vaccine efficacy across pathogens and type of outbreak information for a 28-day memory window.** We present cumulative deaths across the three pathogens of interest (pandemic influenza, SARS-CoV-2, Ebola) alongside three vaccine efficacies ($\varepsilon \in 25\%, 50\%, 90\%$) of interest. Panel **(A)** shows simulations where the outbreak information was global cases as well as the corresponding cumulative cases for each unique combination of pathogen, vaccine efficacy ($\varepsilon$), vaccine opinion ($\rho$) and information sensitivity ($\alpha$). Dark purple hues correspond to more cumulative cases whilst light orange hues correspond to fewer cumulative deaths. Panel **(B)** shows the percent difference in cumulative cases between simulations where the outbreak information was global cases ($\theta_{GC}$) and global deaths ($\theta_{GD}$) for each unique combination of pathogen, vaccine efficacy ($\varepsilon$), vaccine opinion ($\rho$) and information sensitivity ($\alpha$). Blue hues correspond to positive percent differences in cumulative deaths (representing a benefit of global deaths) whilst red hues correspond to negative percent differences in cumulative deaths (representing a benefit of global cases).

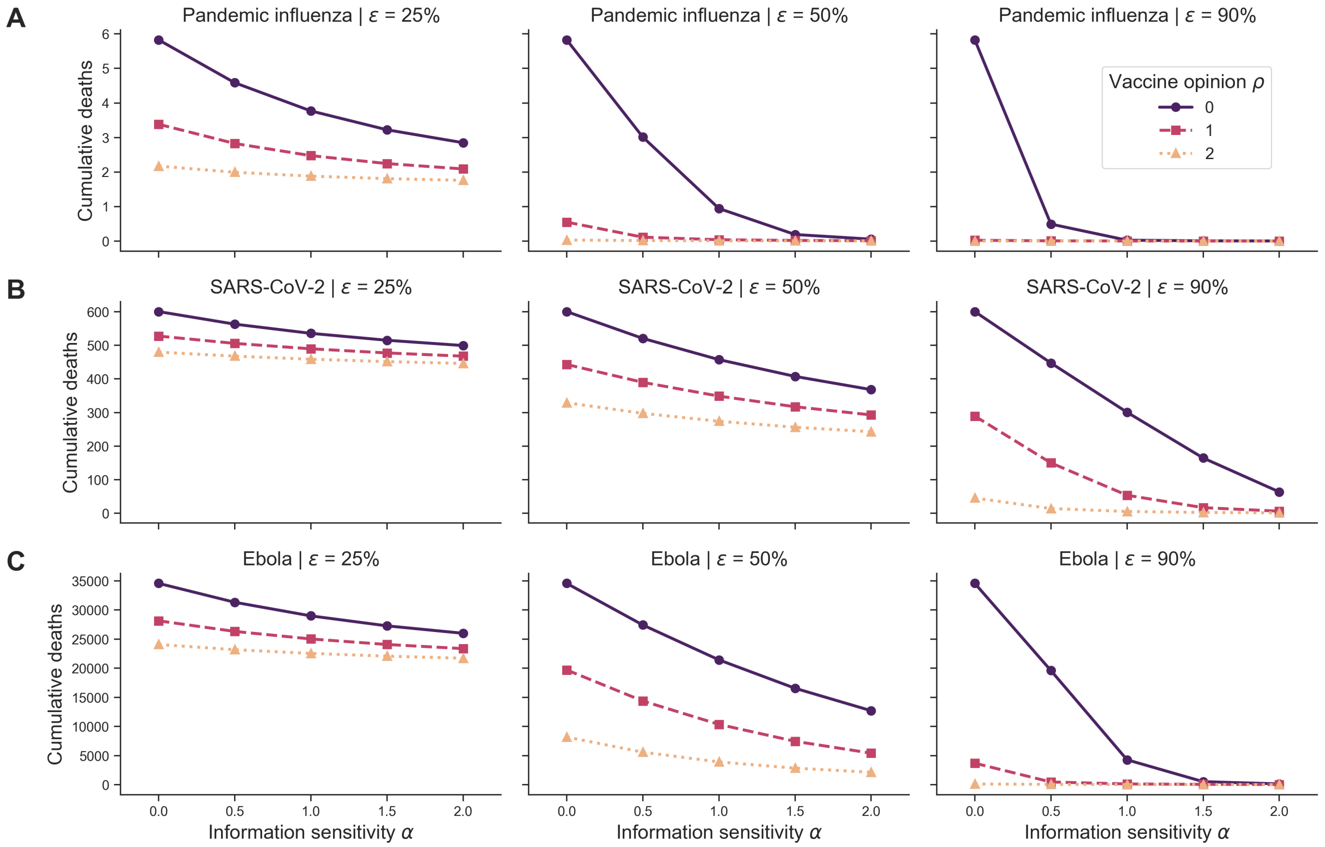

**Figure S8. Homogeneous scenario 2. Cumulative deaths across pathogen systems and vaccine efficacy for homogeneous behavioural configurations and outbreak information based on global cases.** Each row shows cumulative deaths for a pathogen system across different levels of vaccine efficacy ($\varepsilon \in 25\%, 50\%, 90\%$) in different homogeneous behavioural configurations: vaccine-resistant ($\rho = 0$, purple solid line with circle markers), vaccine-hesitant ($\rho = 1$, pink dashed line with square markers) and vaccine-accepting ($\rho = 2$, orange dotted line with triangle markers). The memory window ($\mu$) was fixed at a full history and outbreak information was based on global cases ($\theta_{GC}$).

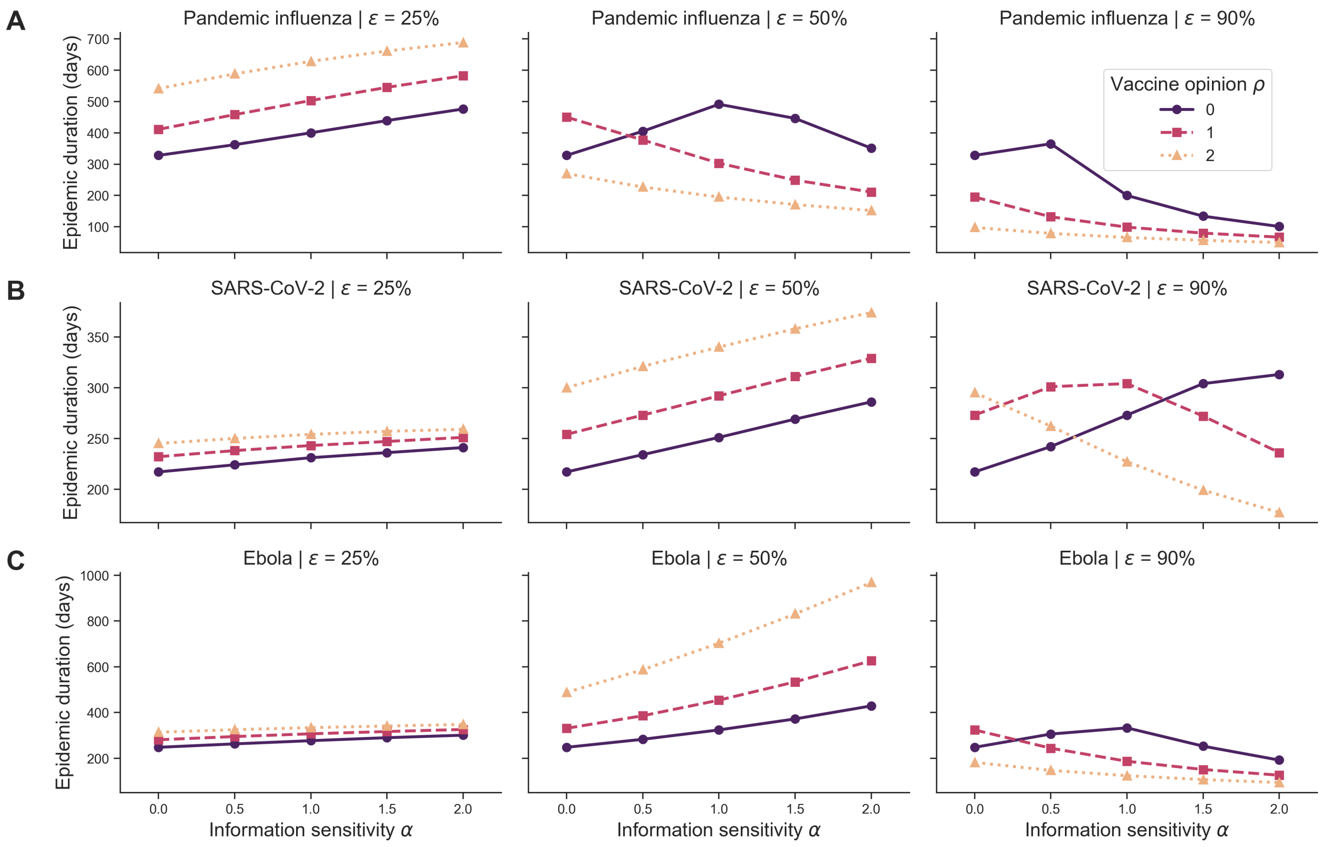

**Figure S9. Homogeneous scenario 2. Epidemic duration in days across pathogen systems and vaccine efficacy for homogeneous behavioural configurations and outbreak information based on global cases.** Each row shows epidemic duration for a pathogen system across different levels of vaccine efficacy ($\varepsilon \in 25\%, 50\%, 90\%$) in different homogeneous behavioural configurations: vaccine-resistant ($\rho = 0$, purple solid line with circle markers), vaccine-hesitant ($\rho = 1$, pink dashed line with square markers) and vaccine-accepting ($\rho = 2$, orange dotted line with triangle markers). The memory window ($\mu$) was fixed at a full history and outbreak information was based on global cases ($\theta_{GC}$).

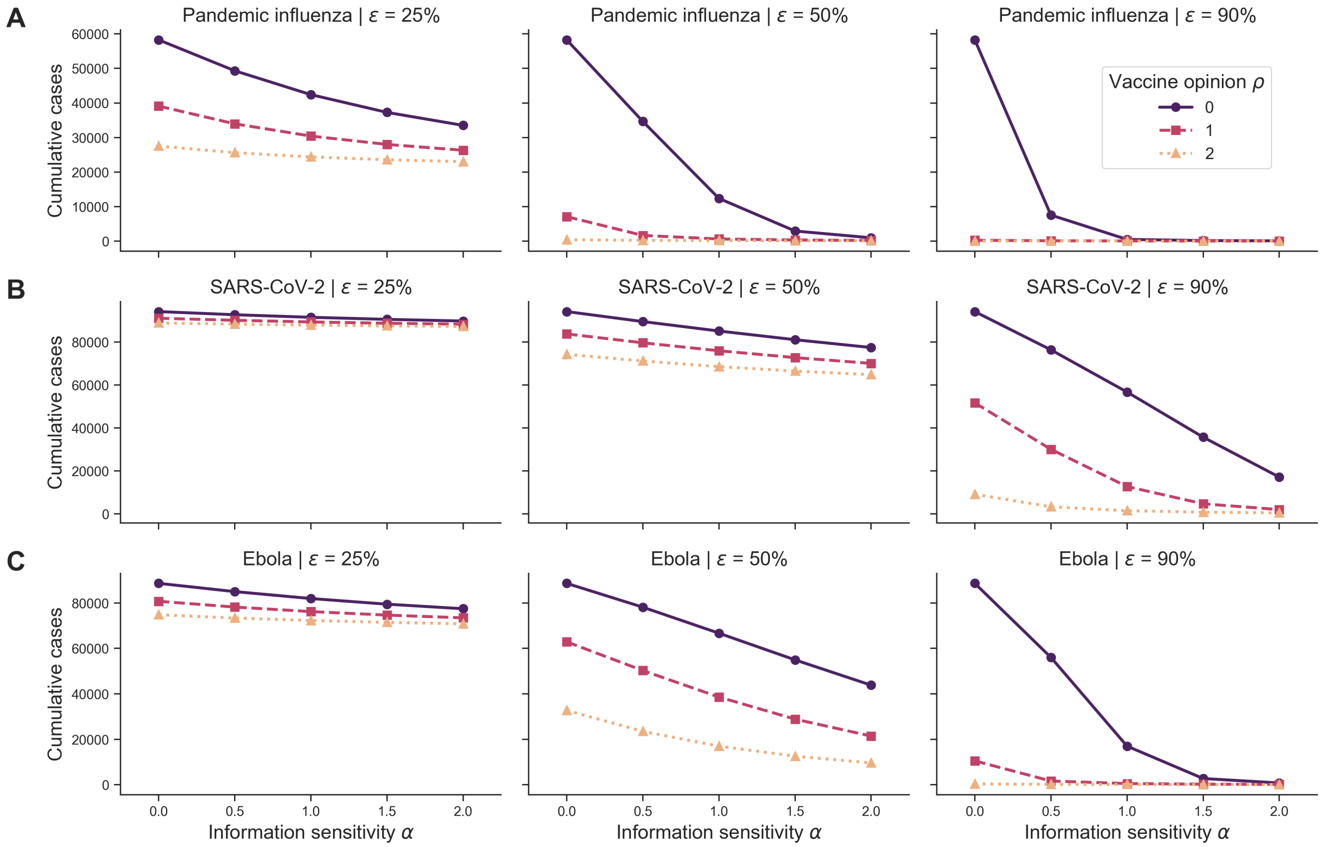

**Figure S10. Homogeneous scenario 2. Cumulative cases across pathogen systems and vaccine efficacy for homogeneous behavioural configurations and outbreak information based on global deaths.** Each row shows cumulative cases for a pathogen system across different levels of vaccine efficacy ($\varepsilon \in 25\%, 50\%, 90\%$) in different homogeneous behavioural configurations: vaccine-resistant ($\rho = 0,$ purple solid line with circle markers), vaccine-hesitant ($\rho = 1,$ pink dashed line with square markers) and vaccine-accepting ($\rho = 2,$ orange dotted line with triangle markers). The memory window ($\mu$) was fixed at a full history and outbreak information was based on global deaths ($\theta_{GD}$).

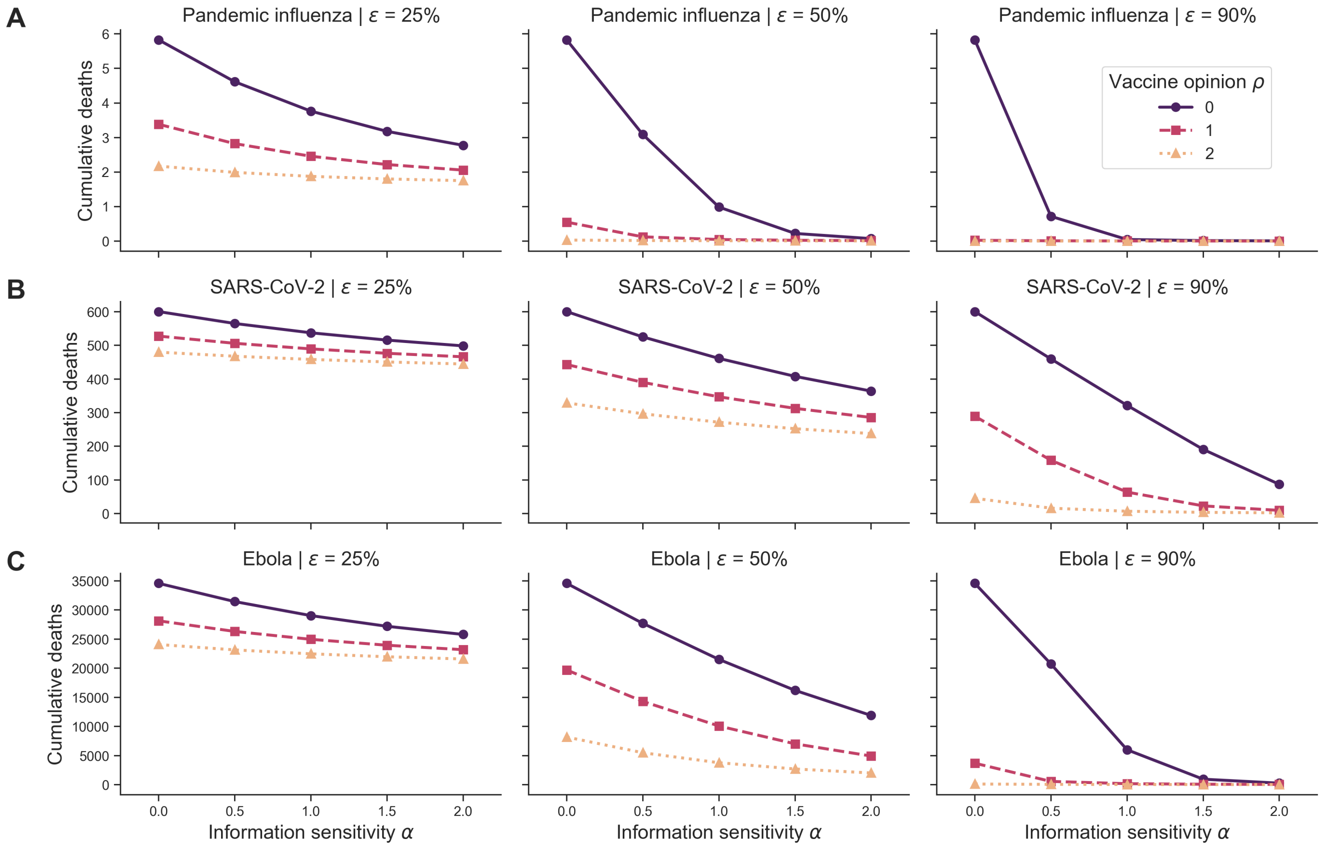

**Figure S11. Homogeneous scenario 2. Cumulative deaths across pathogen systems and vaccine efficacy for homogeneous behavioural configurations and outbreak information based on global deaths.** Each row shows cumulative deaths for a pathogen system across different levels of vaccine efficacy ($\varepsilon \in 25\%, 50\%, 90\%$) in different homogeneous behavioural configurations: vaccine-resistant ($\rho = 0$, purple solid line with circle markers), vaccine-hesitant ($\rho = 1$, pink dashed line with square markers) and vaccine-accepting ($\rho = 2$, orange dotted line with triangle markers). The memory window ($\mu$) was fixed at a full history and outbreak information was based on global deaths ($\theta_{GD}$).

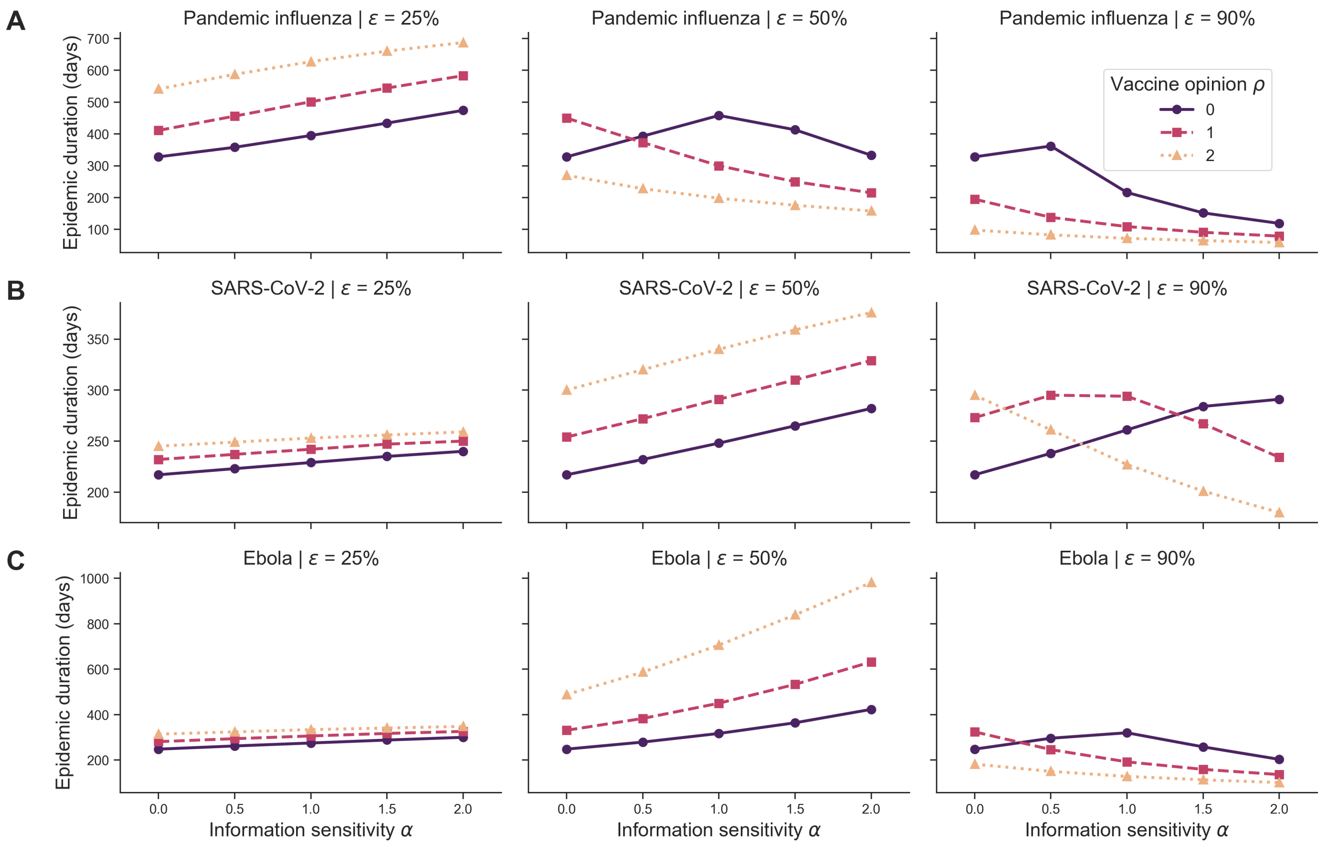

**Figure S12. Homogeneous scenario 2. Epidemic duration in days across pathogen systems and vaccine efficacy for homogeneous behavioural configurations and outbreak information based on global deaths.** Each row shows epidemic duration for a pathogen system across different levels of vaccine efficacy ($\varepsilon \in 25\%, 50\%, 90\%$) in different homogeneous behavioural configurations: vaccine-resistant ($\rho = 0,$ purple solid line with circle markers), vaccine-hesitant ($\rho = 1,$ pink dashed line with square markers) and vaccine-accepting ($\rho = 2,$ orange dotted line with triangle markers). The memory window ($\mu$) was fixed at a full history and outbreak information was based on global deaths ($\theta_{GD}$).

#### S2 Additional heterogeneous population scenario results

##### S2.1 Heterogeneous scenario 1

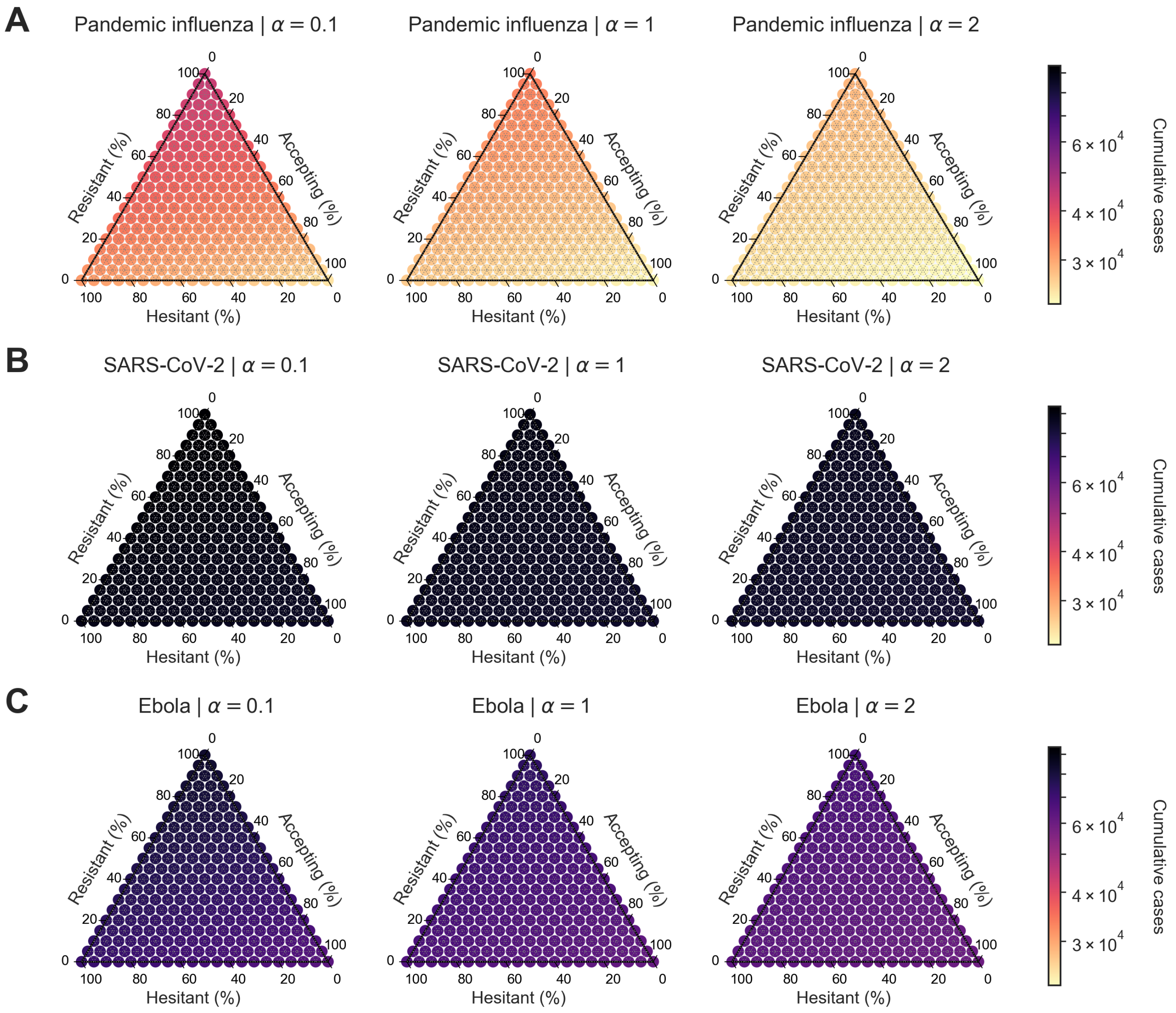

**Figure S13. Heterogeneous scenario 1. Cumulative cases across pathogen systems and information sensitivity, with outbreak information based on local cases and vaccine efficacy of 25%.** For each panel, the ternary plot axes show the percentage of the population assigned to each of three behavioural groups: vaccine-resistant ($\rho = 0$), vaccine-hesitant ($\rho = 1$) and vaccine-accepting ($\rho = 2$). Each row shows cumulative cases for a given pathogen: **(A)** pandemic influenza, **(B)** SARS-CoV-2, **(C)** Ebola. Each column corresponds to a different level of information sensitivity: $\alpha = 0.1$ (first column), $\alpha = 1$ (second column), $\alpha = 2$ (third column). Darker colour hues indicate more severe outcomes in terms of cumulative cases.

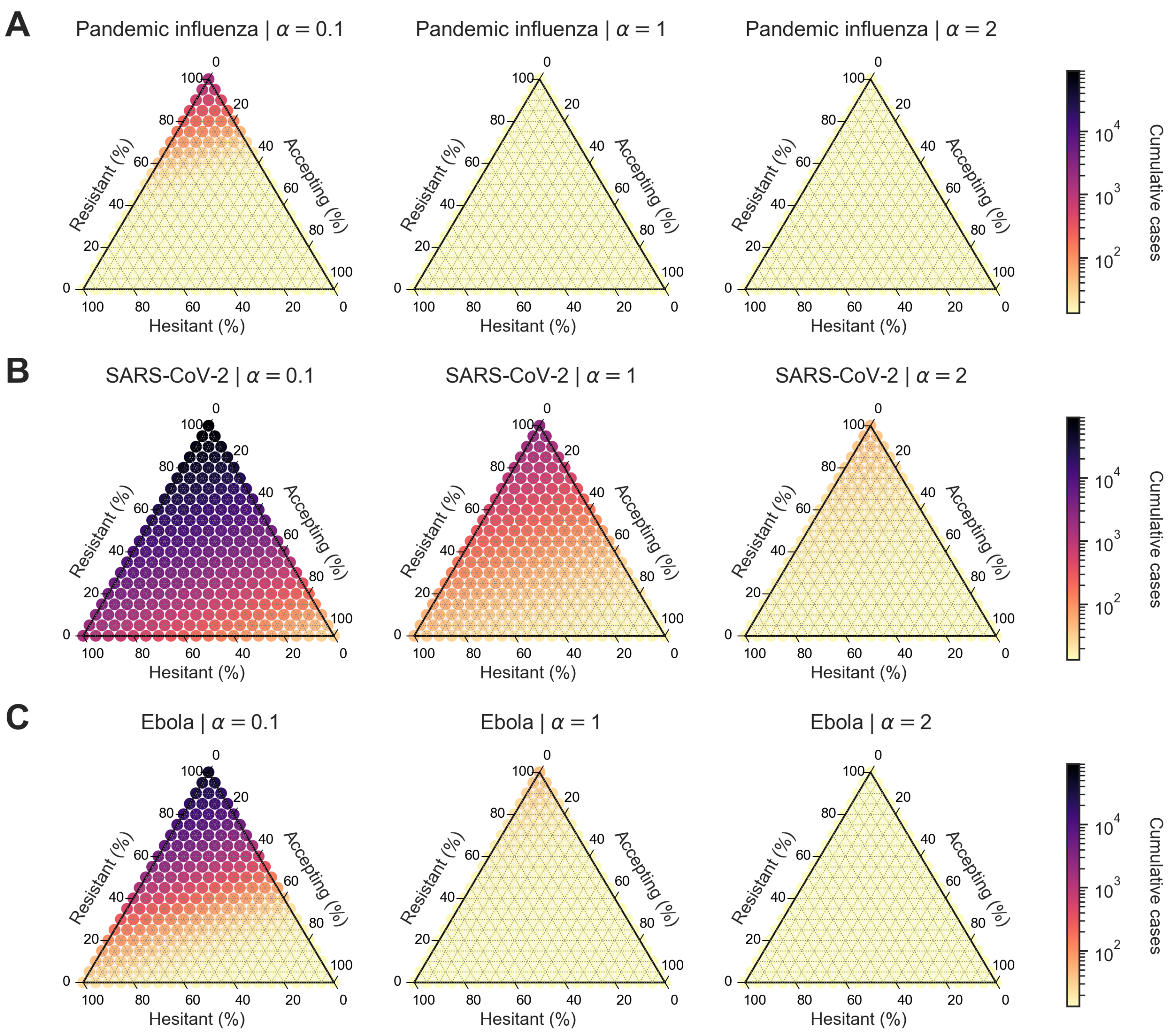

**Figure S14. Heterogeneous scenario 1. Cumulative cases across pathogen systems and information sensitivity, with outbreak information based on local cases and vaccine efficacy of 90%.** For each panel, the ternary plot axes show the percentage of the population assigned to each of three behavioural groups: vaccine-resistant ($\rho = 0$), vaccine-hesitant ($\rho = 1$) and vaccine-accepting ($\rho = 2$). Each row shows cumulative cases for a given pathogen: **(A)** pandemic influenza, **(B)** SARS-CoV-2, **(C)** Ebola. Each column corresponds to a different level of information sensitivity: $\alpha = 0.1$ (first column), $\alpha = 1$ (second column), $\alpha = 2$ (third column). Darker colour hues indicate more severe outcomes in terms of cumulative cases.

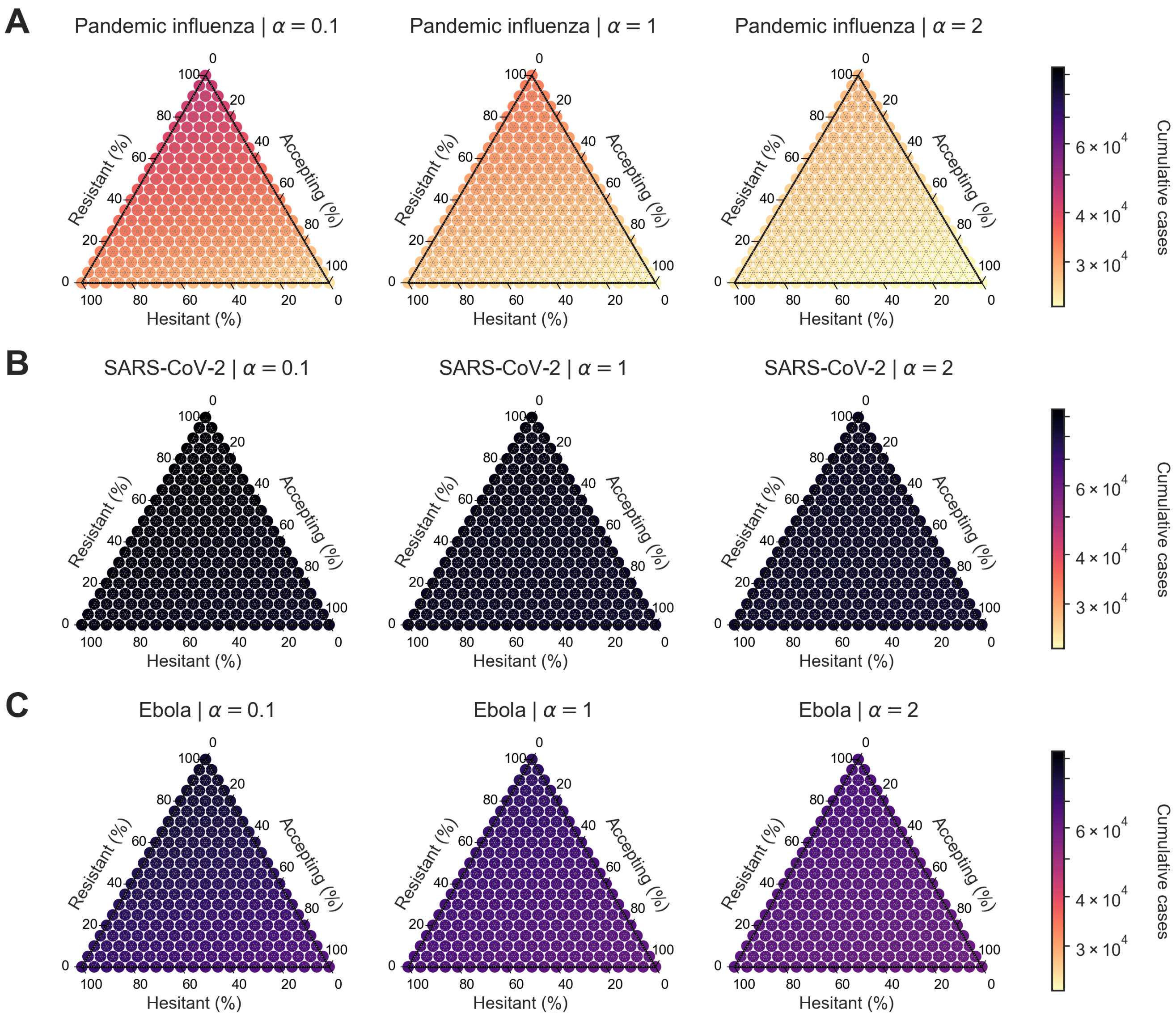

**Figure S15. Heterogeneous scenario 1. Cumulative cases across pathogen systems and information sensitivity, with outbreak information based on local deaths and vaccine efficacy of 25%.** For each panel, the ternary plot axes show the percentage of the population assigned to each of three behavioural groups: vaccine-resistant ($\rho = 0$), vaccine-hesitant ($\rho = 1$) and vaccine-accepting ($\rho = 2$). Each row shows cumulative cases for a given pathogen: **(A)** pandemic influenza, **(B)** SARS-CoV-2, **(C)** Ebola. Each column corresponds to a different level of information sensitivity: $\alpha = 0.1$ (first column), $\alpha = 1$ (second column), $\alpha = 2$ (third column). Darker colour hues indicate more severe outcomes in terms of cumulative cases.

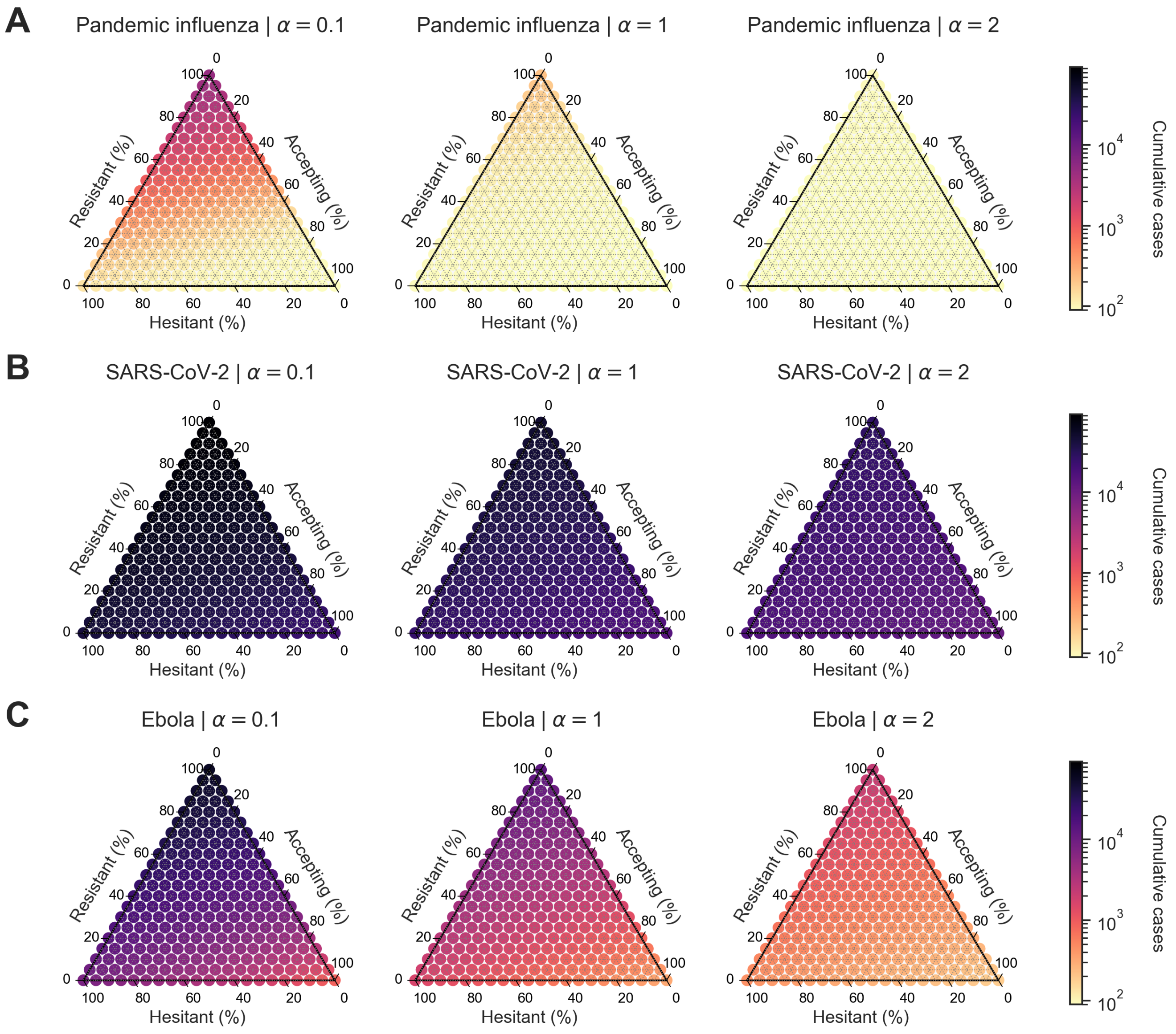

**Figure S16. Heterogeneous scenario 1. Cumulative cases across pathogen systems and information sensitivity, with outbreak information based on local deaths and vaccine efficacy of 50%.** For each panel, the ternary plot axes show the percentage of the population assigned to each of three behavioural groups: vaccine-resistant ($\rho = 0$), vaccine-hesitant ($\rho = 1$) and vaccine-accepting ($\rho = 2$). Each row shows cumulative cases for a given pathogen: **(A)** pandemic influenza, **(B)** SARS-CoV-2, **(C)** Ebola. Each column corresponds to a different level of information sensitivity: $\alpha = 0.1$ (first column), $\alpha = 1$ (second column), $\alpha = 2$ (third column). Darker colour hues indicate more severe outcomes in terms of cumulative cases.

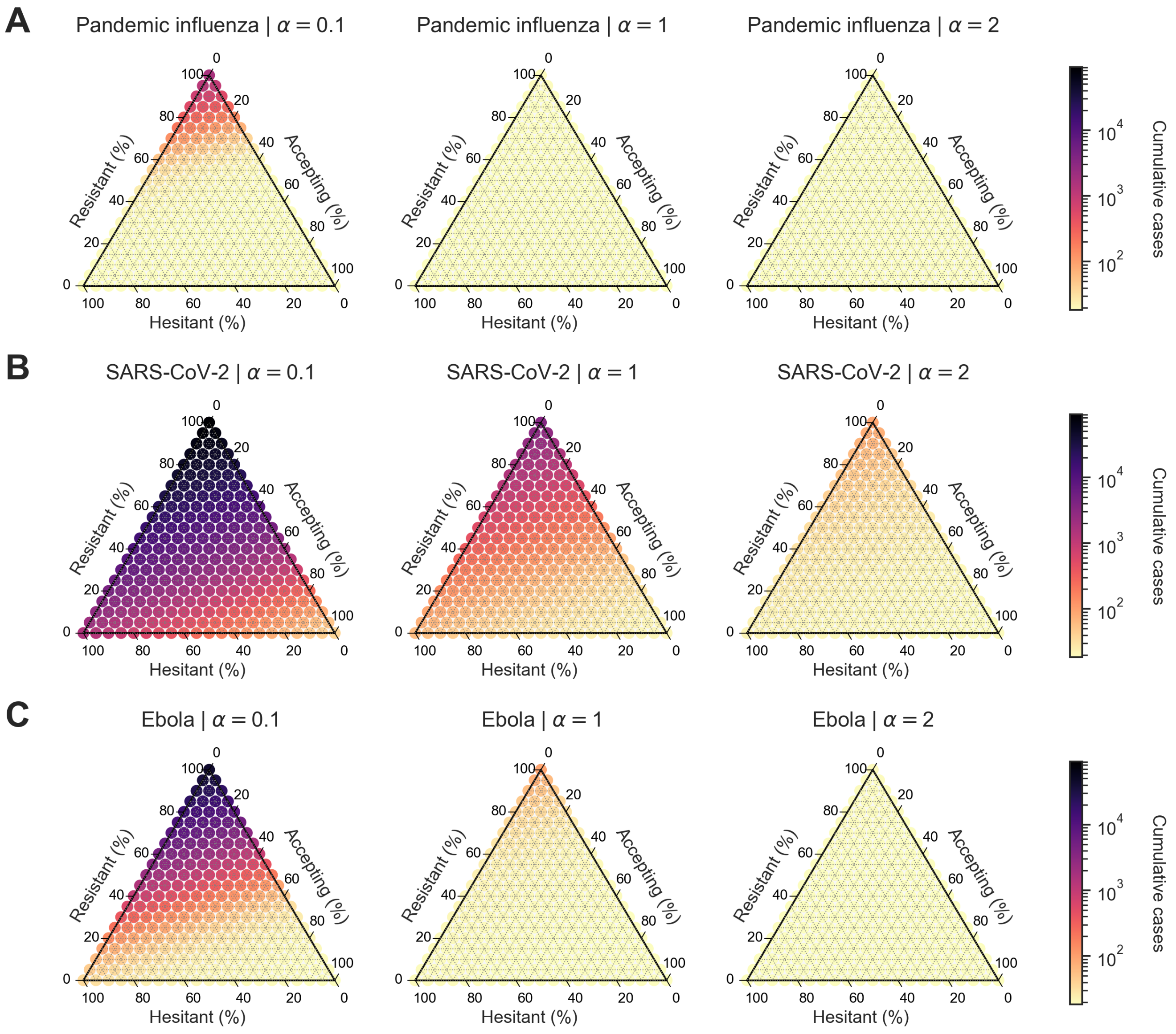

**Figure S17. Heterogeneous scenario 1. Cumulative cases across pathogen systems and information sensitivity, with outbreak information based on local deaths and vaccine efficacy of 90%.** For each panel, the ternary plot axes show the percentage of the population assigned to each of three behavioural groups: vaccine-resistant ($\rho = 0$), vaccine-hesitant ($\rho = 1$) and vaccine-accepting ($\rho = 2$). Each row shows cumulative cases for a given pathogen: **(A)** pandemic influenza, **(B)** SARS-CoV-2, **(C)** Ebola. Each column corresponds to a different level of information sensitivity: $\alpha = 0.1$ (first column), $\alpha = 1$ (second column), $\alpha = 2$ (third column). Darker colour hues indicate more severe outcomes in terms of cumulative cases.

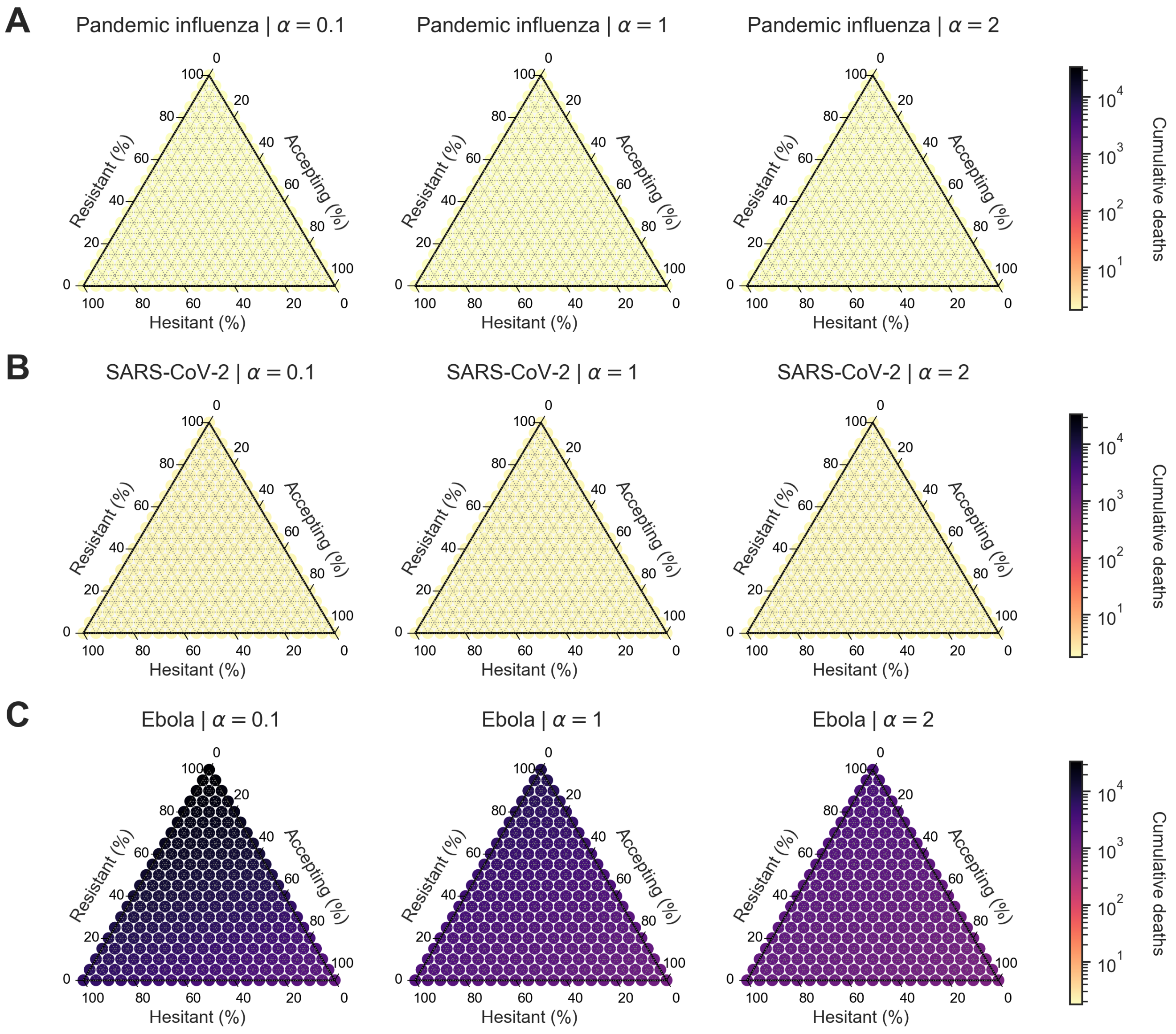

**Figure S18. Heterogeneous scenario 1. Cumulative deaths across pathogen systems and information sensitivity, with outbreak information based on local cases and vaccine efficacy of 25%.** For each panel, the ternary plot axes show the percentage of the population assigned to each of three behavioural groups: vaccine-resistant ($\rho = 0$), vaccine-hesitant ($\rho = 1$) and vaccine-accepting ($\rho = 2$). Each row shows cumulative deaths for a given pathogen: **(A)** pandemic influenza, **(B)** SARS-CoV-2, **(C)** Ebola. Each column corresponds to a different level of information sensitivity: $\alpha = 0.1$ (first column), $\alpha = 1$ (second column), $\alpha = 2$ (third column). Darker colour hues indicate more severe outcomes in terms of cumulative deaths.

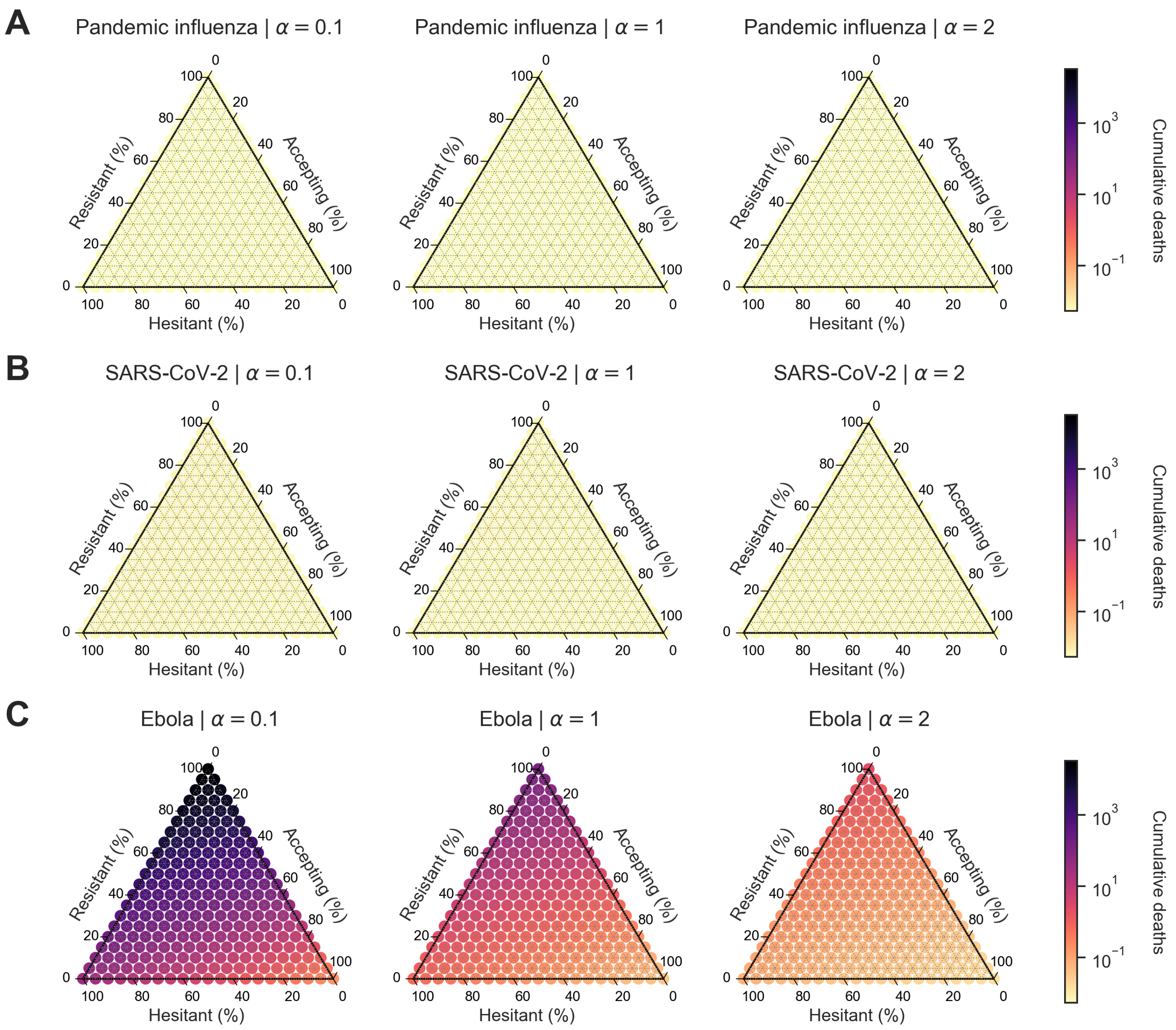

**Figure S19. Heterogeneous scenario 1. Cumulative deaths across pathogen systems and information sensitivity, with outbreak information based on local cases and vaccine efficacy of 50%.** For each panel, the ternary plot axes show the percentage of the population assigned to each of three behavioural groups: vaccine-resistant ($\rho = 0$), vaccine-hesitant ($\rho = 1$) and vaccine-accepting ($\rho = 2$). Each row shows cumulative deaths for a given pathogen: **(A)** pandemic influenza, **(B)** SARS-CoV-2, **(C)** Ebola. Each column corresponds to a different level of information sensitivity: $\alpha = 0.1$ (first column), $\alpha = 1$ (second column), $\alpha = 2$ (third column). Darker colour hues indicate more severe outcomes in terms of cumulative deaths.

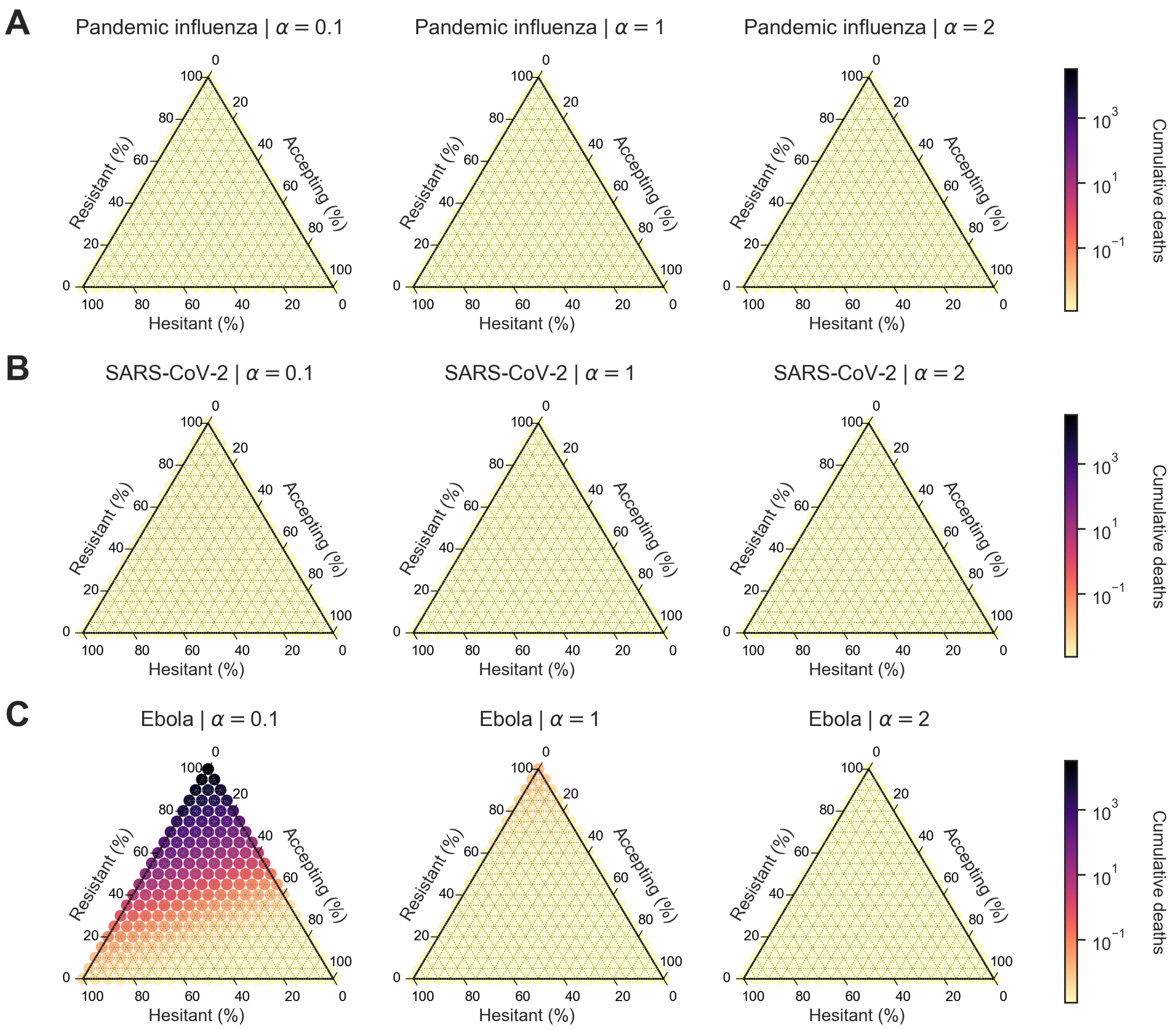

**Figure S20. Heterogeneous scenario 1. Cumulative deaths across pathogen systems and information sensitivity, with outbreak information based on local cases and vaccine efficacy of 90%.** For each panel, the ternary plot axes show the percentage of the population assigned to each of three behavioural groups: vaccine-resistant ($\rho = 0$), vaccine-hesitant ($\rho = 1$) and vaccine-accepting ($\rho = 2$). Each row shows cumulative deaths for a given pathogen: **(A)** pandemic influenza, **(B)** SARS-CoV-2, **(C)** Ebola. Each column corresponds to a different level of information sensitivity: $\alpha = 0.1$ (first column), $\alpha = 1$ (second column), $\alpha = 2$ (third column). Darker colour hues indicate more severe outcomes in terms of cumulative deaths.

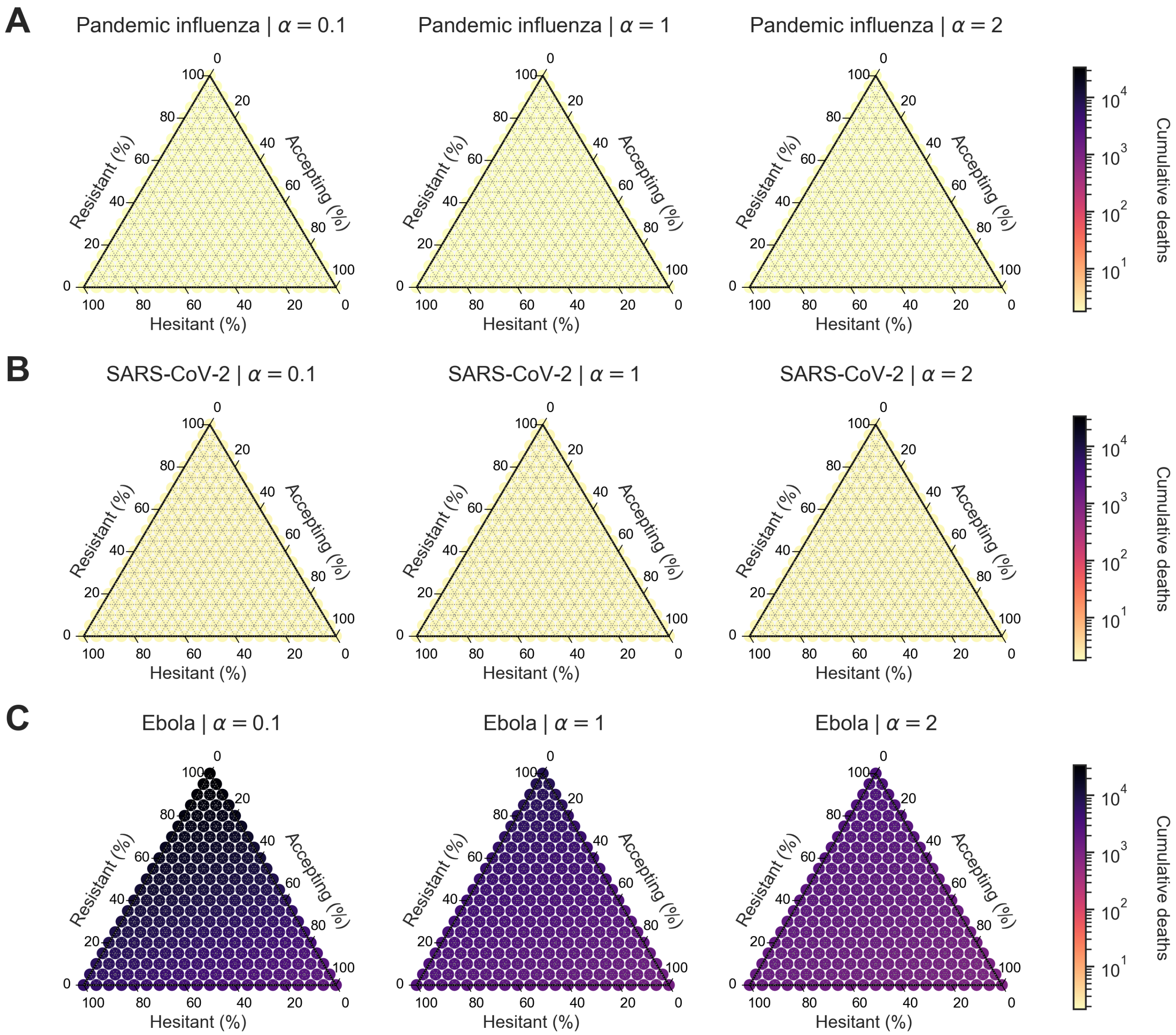

**Figure S21. Heterogeneous scenario 1. Cumulative deaths across pathogen systems and information sensitivity, with outbreak information based on local deaths and vaccine efficacy of 25%.** For each panel, the ternary plot axes show the percentage of the population assigned to each of three behavioural groups: vaccine-resistant ($\rho = 0$), vaccine-hesitant ($\rho = 1$) and vaccine-accepting ($\rho = 2$). Each row shows cumulative deaths for a given pathogen: **(A)** pandemic influenza, **(B)** SARS-CoV-2, **(C)** Ebola. Each column corresponds to a different level of information sensitivity: $\alpha = 0.1$ (first column), $\alpha = 1$ (second column), $\alpha = 2$ (third column). Darker colour hues indicate more severe outcomes in terms of cumulative deaths.

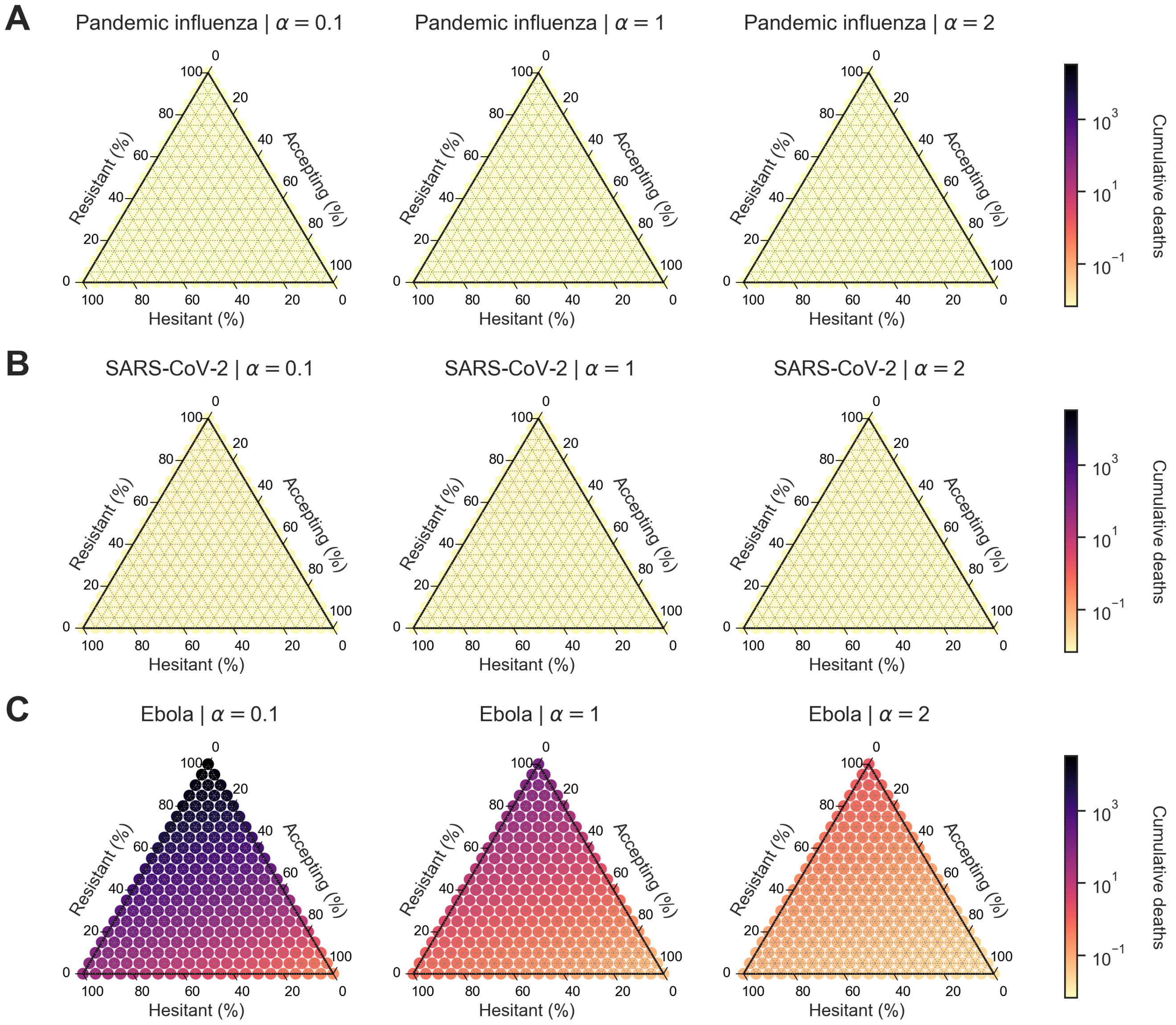

**Figure S22. Heterogeneous scenario 1. Cumulative deaths across pathogen systems and information sensitivity, with outbreak information based on local deaths and vaccine efficacy of 50%.** For each panel, the ternary plot axes show the percentage of the population assigned to each of three behavioural groups: vaccine-resistant ($\rho = 0$), vaccine-hesitant ($\rho = 1$) and vaccine-accepting ($\rho = 2$). Each row shows cumulative deaths for a given pathogen: **(A)** pandemic influenza, **(B)** SARS-CoV-2, **(C)** Ebola. Each column corresponds to a different level of information sensitivity: $\alpha = 0.1$ (first column), $\alpha = 1$ (second column), $\alpha = 2$ (third column). Darker colour hues indicate more severe outcomes in terms of cumulative deaths.

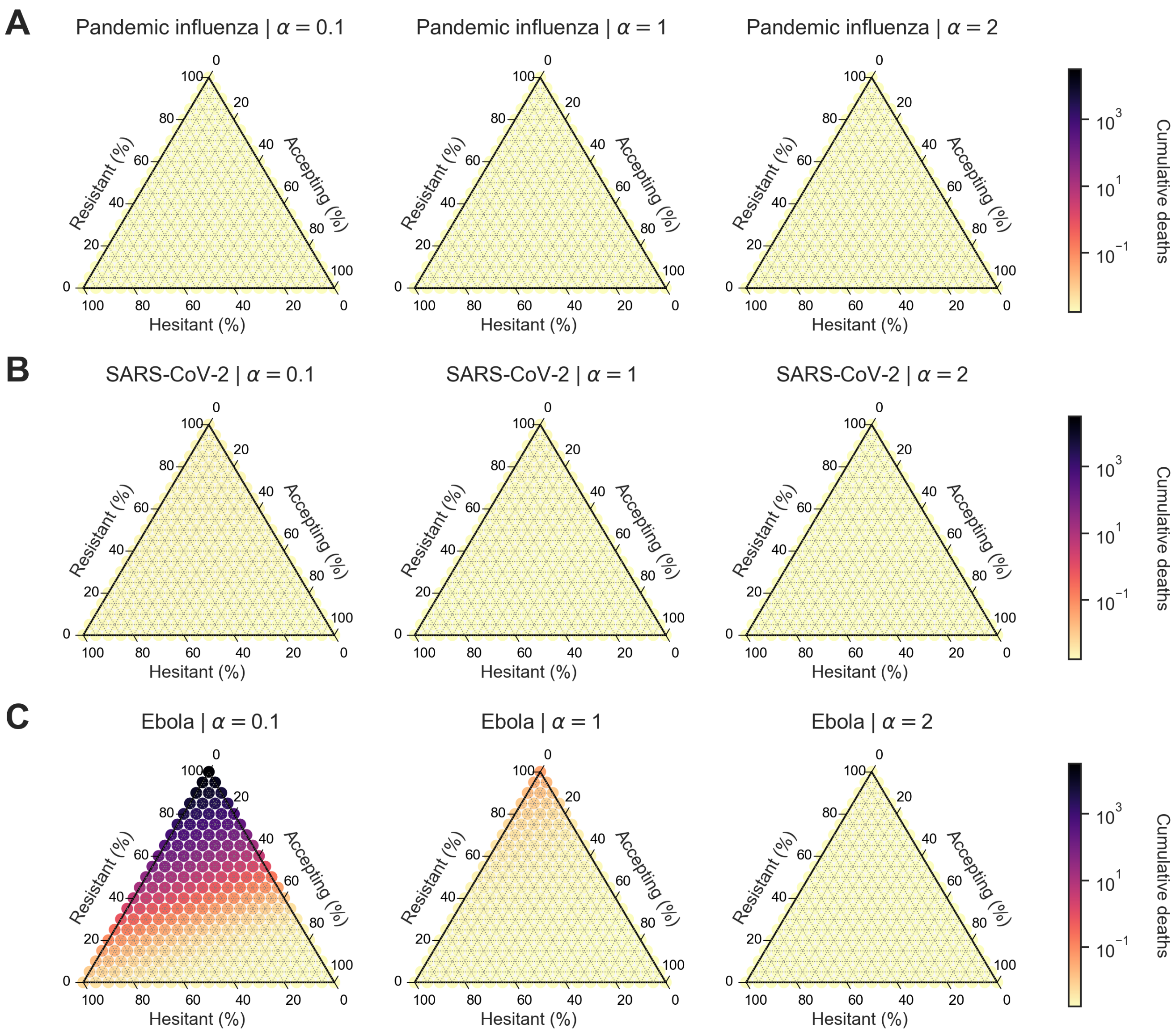

**Figure S23. Heterogeneous scenario 1. Cumulative deaths across pathogen systems and information sensitivity, with outbreak information based on local deaths and vaccine efficacy of 90%.** For each panel, the ternary plot axes show the percentage of the population assigned to each of three behavioural groups: vaccine-resistant ($\rho = 0$), vaccine-hesitant ($\rho = 1$) and vaccine-accepting ($\rho = 2$). Each row shows cumulative deaths for a given pathogen: **(A)** pandemic influenza, **(B)** SARS-CoV-2, **(C)** Ebola. Each column corresponds to a different level of information sensitivity: $\alpha = 0.1$ (first column), $\alpha = 1$ (second column), $\alpha = 2$ (third column). Darker colour hues indicate more severe outcomes in terms of cumulative deaths.

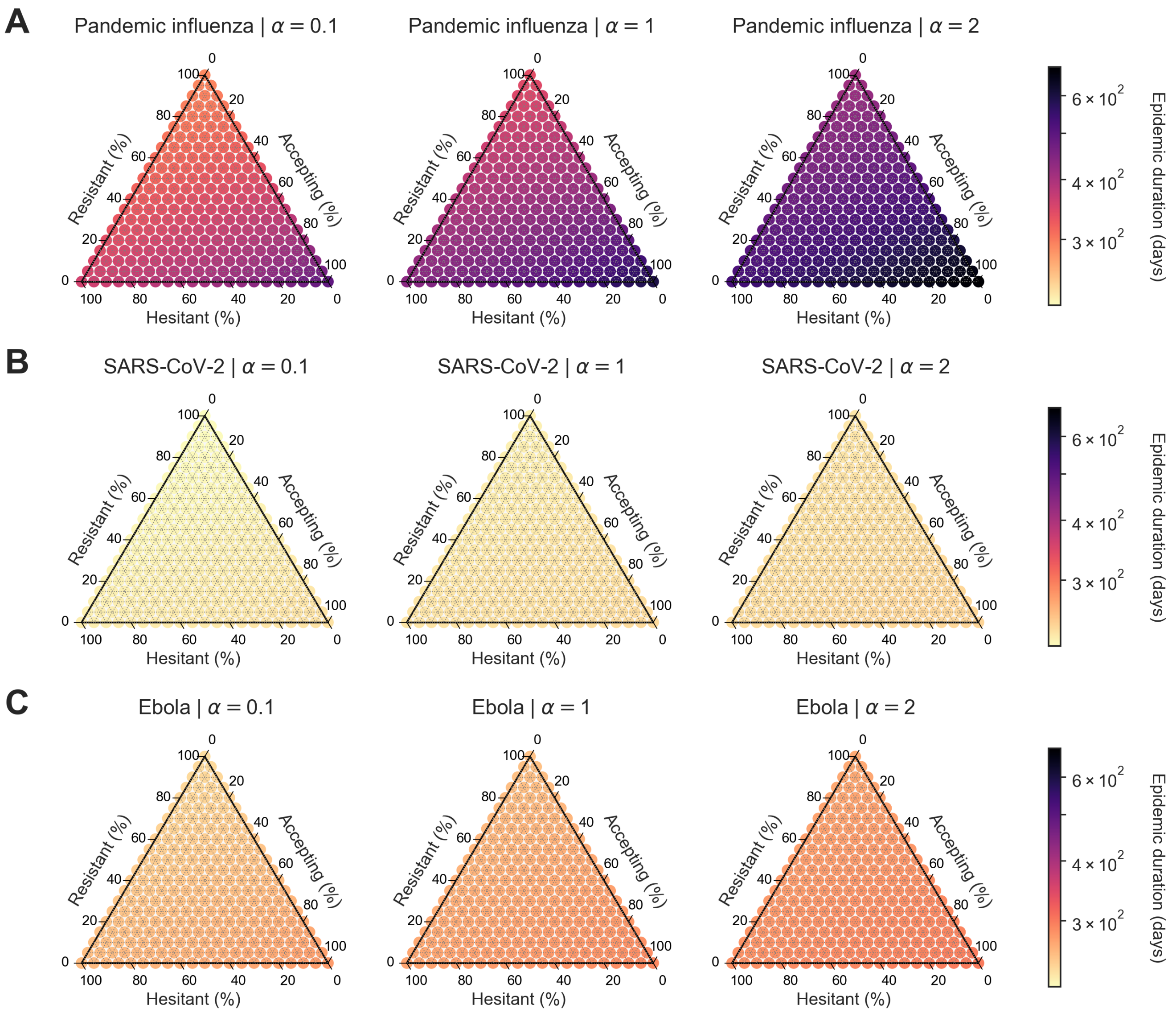

**Figure S24. Heterogeneous scenario 1. Epidemic duration in days across pathogen systems and information sensitivity, with outbreak information based on local cases and vaccine efficacy of 25%.** For each panel, the ternary plot axes show the percentage of the population assigned to each of three behavioural groups: vaccine-resistant ($\rho = 0$), vaccine-hesitant ($\rho = 1$) and vaccine-accepting ($\rho = 2$). Each row shows epidemic duration for a given pathogen: **(A)** pandemic influenza, **(B)** SARS-CoV-2, **(C)** Ebola. Each column corresponds to a different level of information sensitivity: $\alpha = 0.1$ (first column), $\alpha = 1$ (second column), $\alpha = 2$ (third column). Darker colour hues indicate more severe outcomes in terms of epidemic duration.

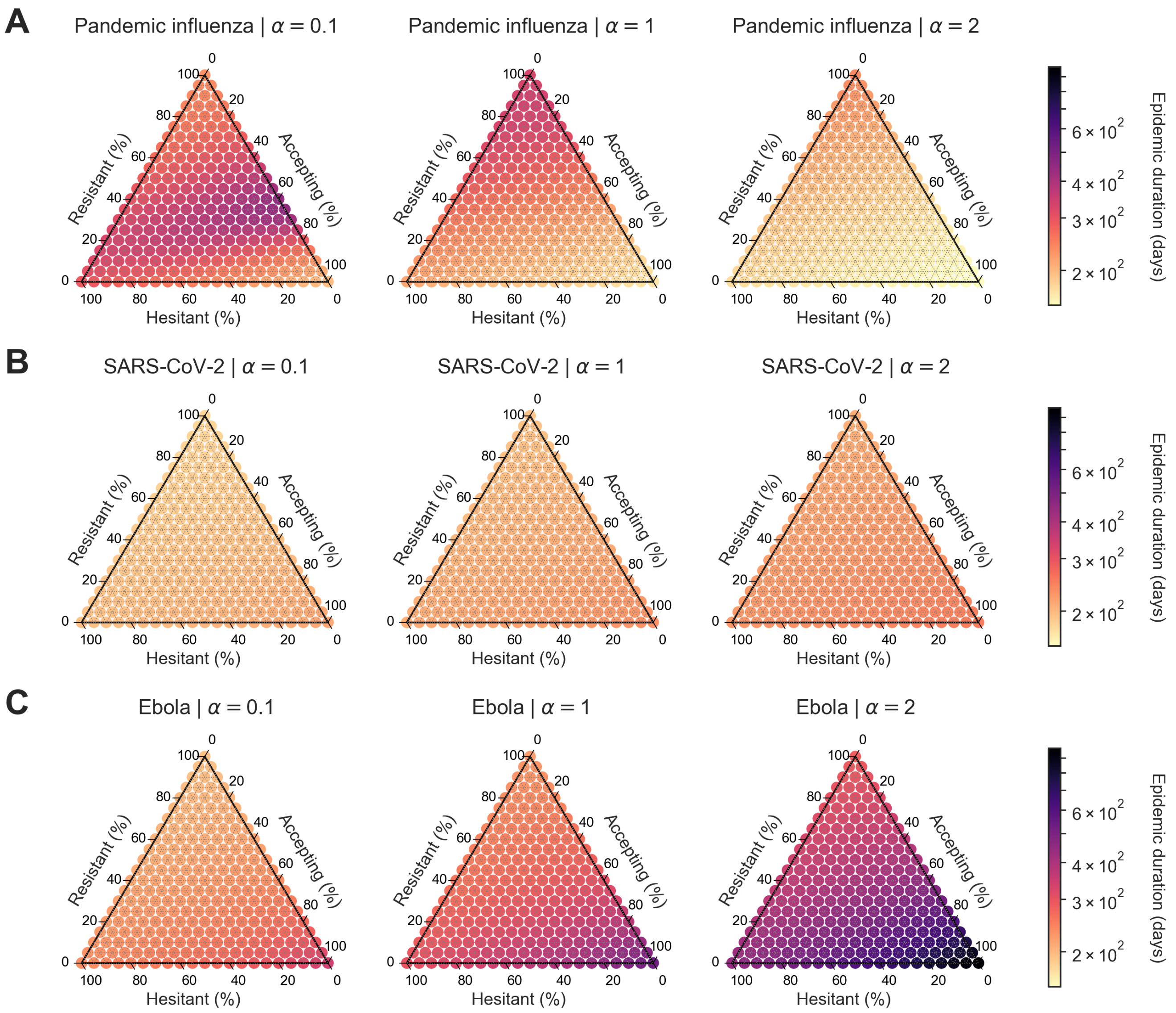

**Figure S25. Heterogeneous scenario 1. Epidemic duration in days across pathogen systems and information sensitivity, with outbreak information based on local cases and vaccine efficacy of 50%.** For each panel, the ternary plot axes show the percentage of the population assigned to each of three behavioural groups: vaccine-resistant ($\rho = 0$), vaccine-hesitant ($\rho = 1$) and vaccine-accepting ($\rho = 2$). Each row shows epidemic duration for a given pathogen: **(A)** pandemic influenza, **(B)** SARS-CoV-2, **(C)** Ebola. Each column corresponds to a different level of information sensitivity: $\alpha = 0.1$ (first column), $\alpha = 1$ (second column), $\alpha = 2$ (third column). Darker colour hues indicate more severe outcomes in terms of epidemic duration.

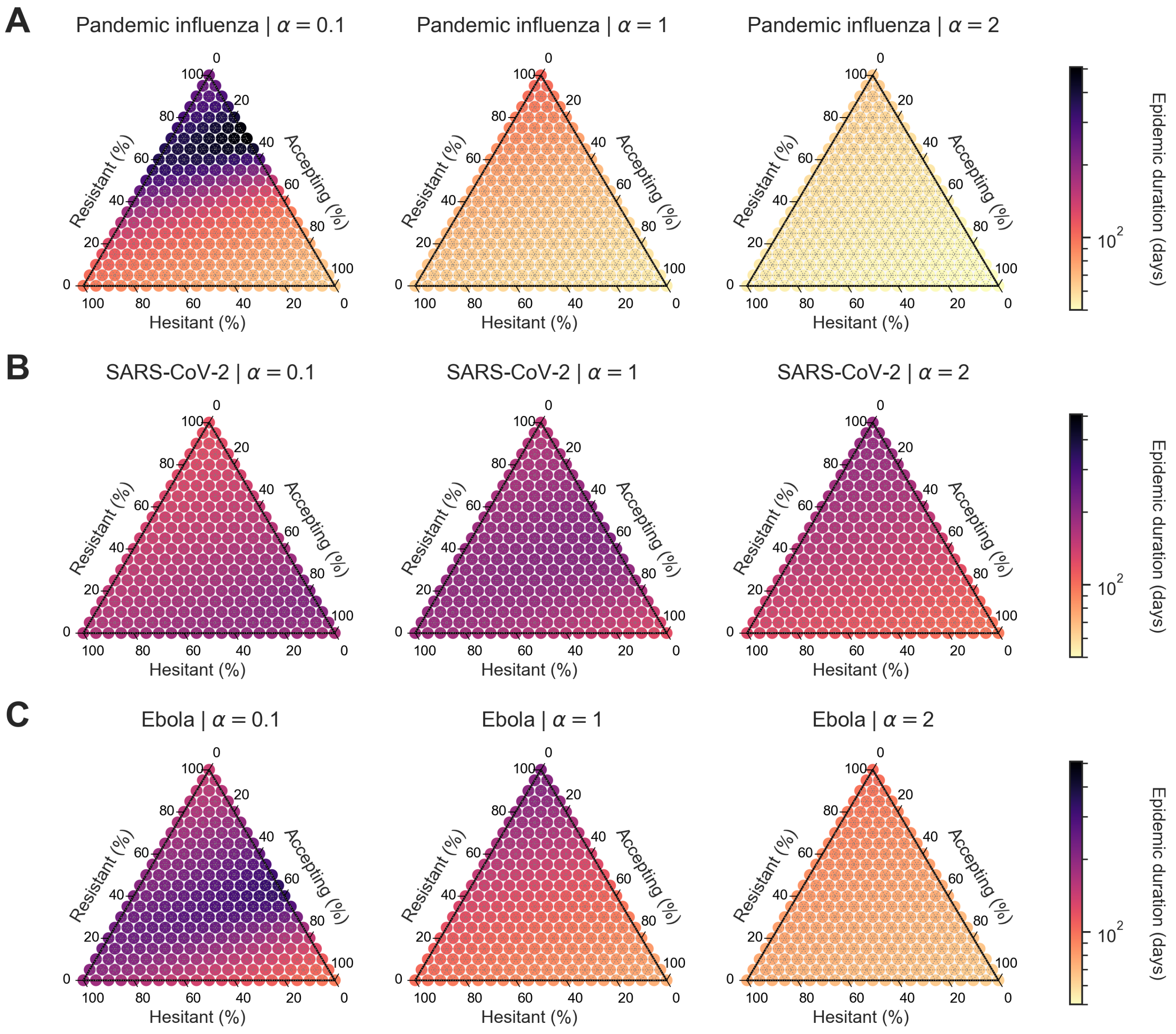

**Figure S26. Heterogeneous scenario 1. Epidemic duration in days across pathogen systems and information sensitivity, with outbreak information based on local cases and vaccine efficacy of 90%.** For each panel, the ternary plot axes show the percentage of the population assigned to each of three behavioural groups: vaccine-resistant ($\rho = 0$), vaccine-hesitant ($\rho = 1$) and vaccine-accepting ($\rho = 2$). Each row shows epidemic duration for a given pathogen: **(A)** pandemic influenza, **(B)** SARS-CoV-2, **(C)** Ebola. Each column corresponds to a different level of information sensitivity: $\alpha = 0.1$ (first column), $\alpha = 1$ (second column), $\alpha = 2$ (third column). Darker colour hues indicate more severe outcomes in terms of epidemic duration.

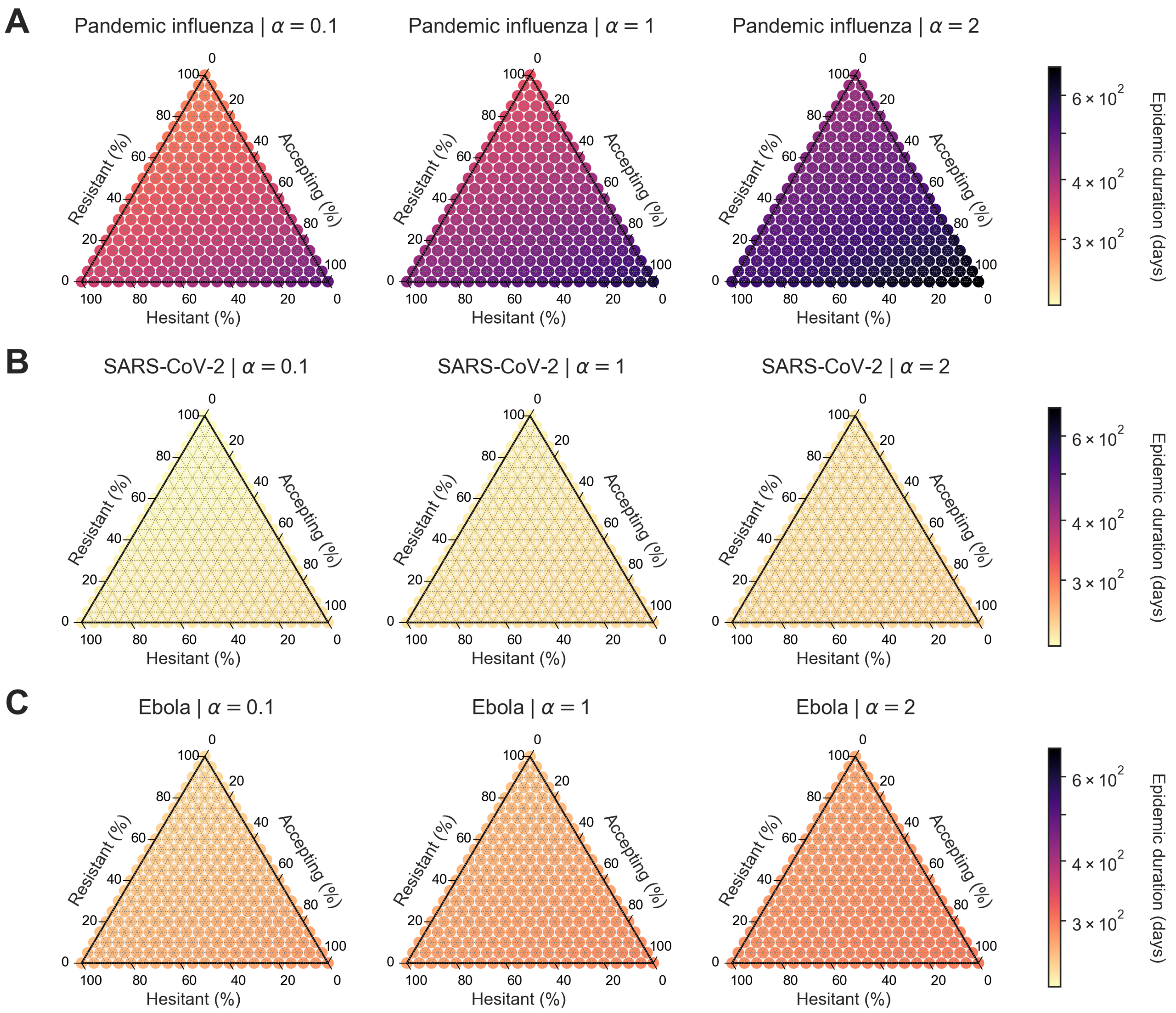

**Figure S27. Heterogeneous scenario 1. Epidemic duration in days across pathogen systems and information sensitivity, with outbreak information based on local deaths and vaccine efficacy of 25%.** For each panel, the ternary plot axes show the percentage of the population assigned to each of three behavioural groups: vaccine-resistant ($\rho = 0$), vaccine-hesitant ($\rho = 1$) and vaccine-accepting ($\rho = 2$). Each row shows epidemic duration for a given pathogen: **(A)** pandemic influenza, **(B)** SARS-CoV-2, **(C)** Ebola. Each column corresponds to a different level of information sensitivity: $\alpha = 0.1$ (first column), $\alpha = 1$ (second column), $\alpha = 2$ (third column). Darker colour hues indicate more severe outcomes in terms of epidemic duration.

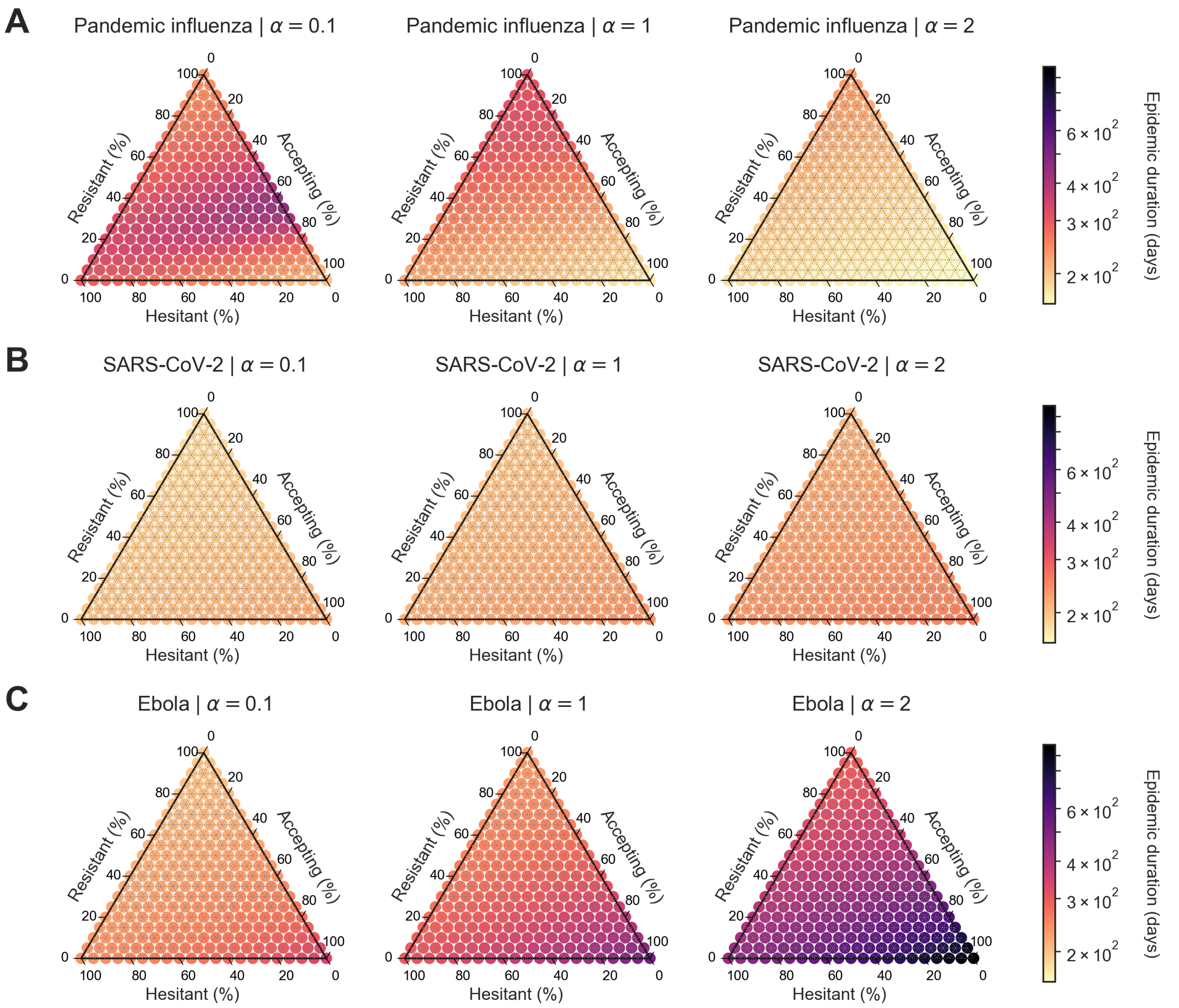

**Figure S28. Heterogeneous scenario 1. Epidemic duration in days across pathogen systems and information sensitivity, with outbreak information based on local deaths and vaccine efficacy of 50%.** For each panel, the ternary plot axes show the percentage of the population assigned to each of three behavioural groups: vaccine-resistant ($\rho = 0$), vaccine-hesitant ($\rho = 1$) and vaccine-accepting ($\rho = 2$). Each row shows epidemic duration for a given pathogen: **(A)** pandemic influenza, **(B)** SARS-CoV-2, **(C)** Ebola. Each column corresponds to a different level of information sensitivity: $\alpha = 0.1$ (first column), $\alpha = 1$ (second column), $\alpha = 2$ (third column). Darker colour hues indicate more severe outcomes in terms of epidemic duration.

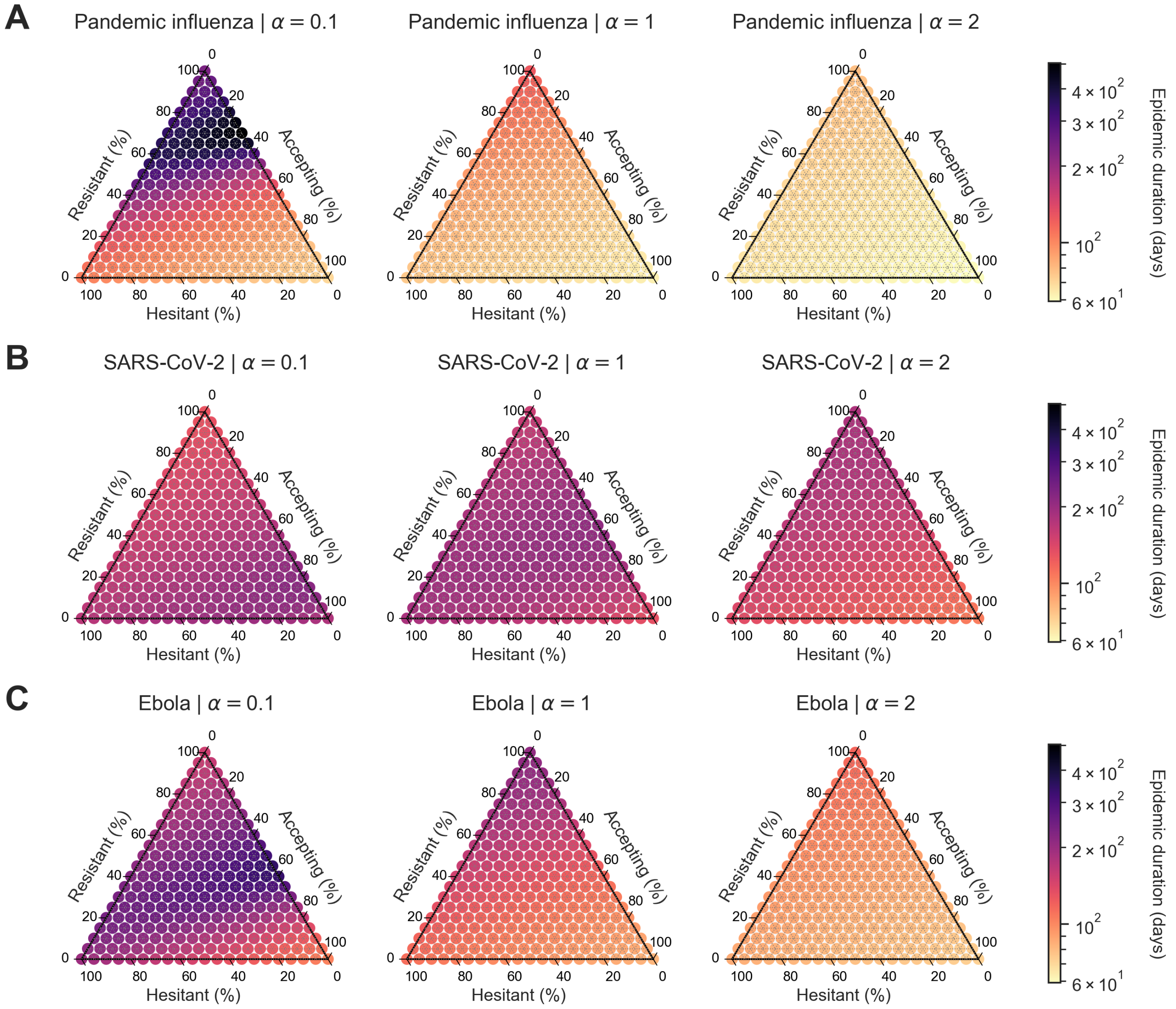

**Figure S29. Heterogeneous scenario 1. Epidemic duration in days across pathogen systems and information sensitivity, with outbreak information based on local deaths and vaccine efficacy of 90%.** For each panel, the ternary plot axes show the percentage of the population assigned to each of three behavioural groups: vaccine-resistant ($\rho = 0$), vaccine-hesitant ($\rho = 1$) and vaccine-accepting ($\rho = 2$). Each row shows epidemic duration for a given pathogen: **(A)** pandemic influenza, **(B)** SARS-CoV-2, **(C)** Ebola. Each column corresponds to a different level of information sensitivity: $\alpha = 0.1$ (first column), $\alpha = 1$ (second column), $\alpha = 2$ (third column). Darker colour hues indicate more severe outcomes in terms of epidemic duration.

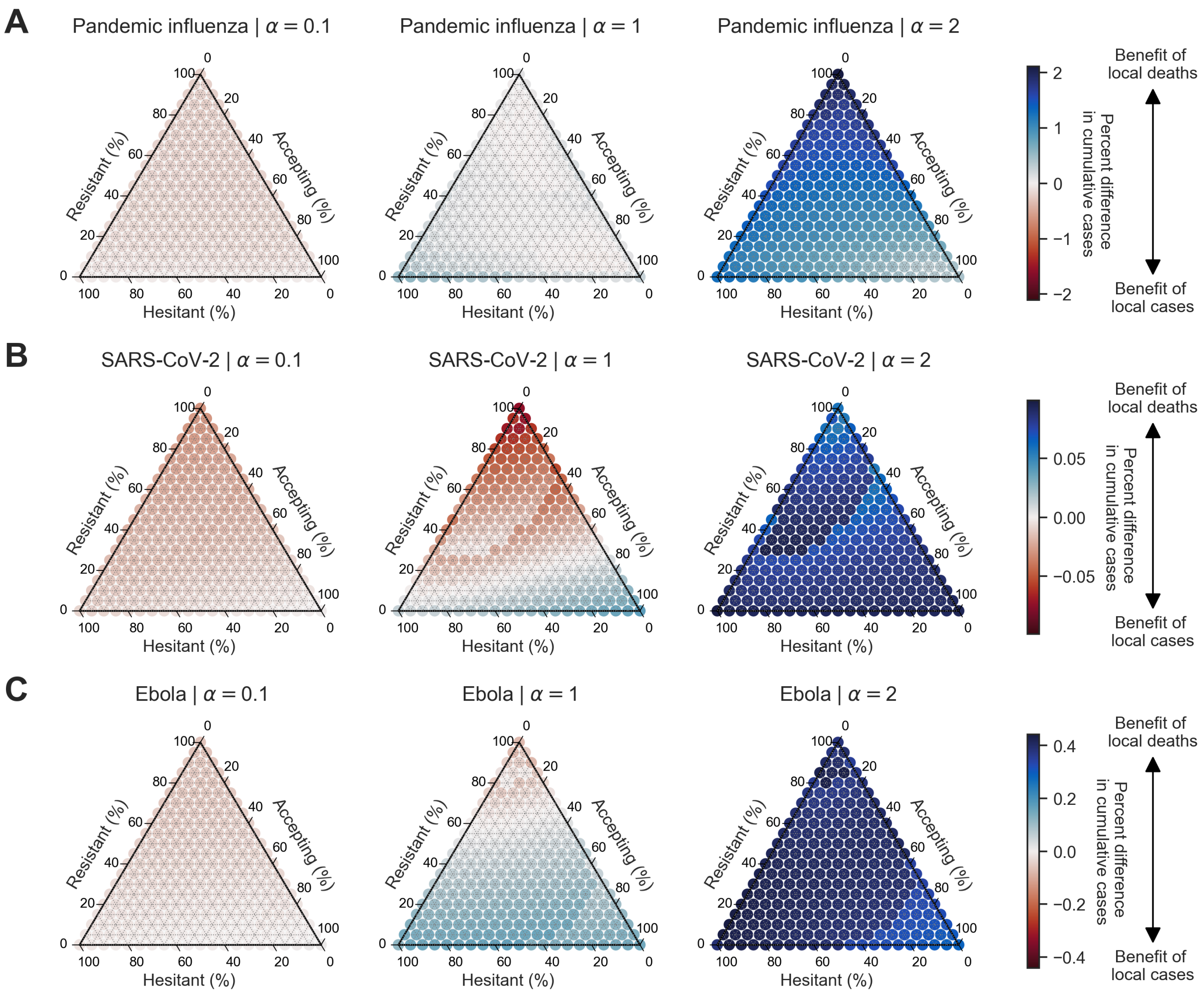

**Figure S30. Heterogeneous scenario 1. Percent difference in cumulative cases between local cases and local deaths across pathogen systems and information sensitivity with a vaccine efficacy of 25%.** For each panel, the ternary plot axes show the percentage of the population assigned to each of three behavioural groups: vaccine-resistant ($\rho = 0$), vaccine-hesitant ($\rho = 1$) and vaccine-accepting ($\rho = 2$). Each row shows percent difference in cumulative cases between outbreak information scenarios based on local cases and local deaths for a given pathogen: **(A)** pandemic influenza, **(B)** SARS-CoV-2, **(C)** Ebola. Each column corresponds to a different level of information sensitivity: $\alpha = 0.1$ (first column), $\alpha = 1$ (second column), $\alpha = 2$ (third column). Darker colour hues indicate more severe outcomes in terms of percent difference in cumulative cases.

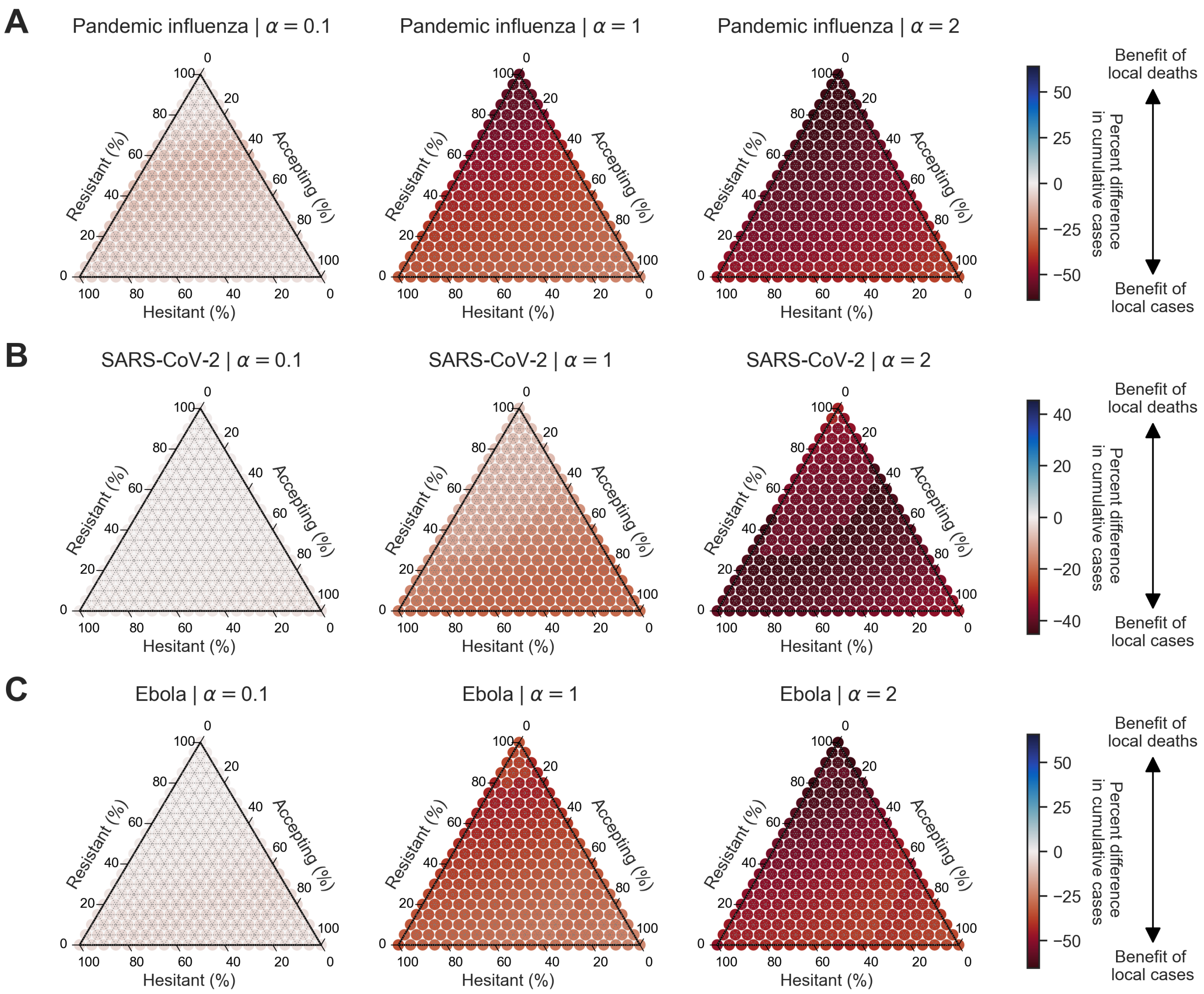

**Figure S31. Heterogeneous scenario 1. Percent difference in cumulative cases between local cases and local deaths across pathogen systems and information sensitivity with a vaccine efficacy of 90%.** For each panel, the ternary plot axes show the percentage of the population assigned to each of three behavioural groups: vaccine-resistant ($\rho = 0$), vaccine-hesitant ($\rho = 1$) and vaccine-accepting ($\rho = 2$). Each row shows percent difference in cumulative cases between outbreak information scenarios based on local cases and local deaths for a given pathogen: **(A)** pandemic influenza, **(B)** SARS-CoV-2, **(C)** Ebola. Each column corresponds to a different level of information sensitivity: $\alpha = 0.1$ (first column), $\alpha = 1$ (second column), $\alpha = 2$ (third column). Darker colour hues indicate more severe outcomes in terms of percent difference in cumulative cases.

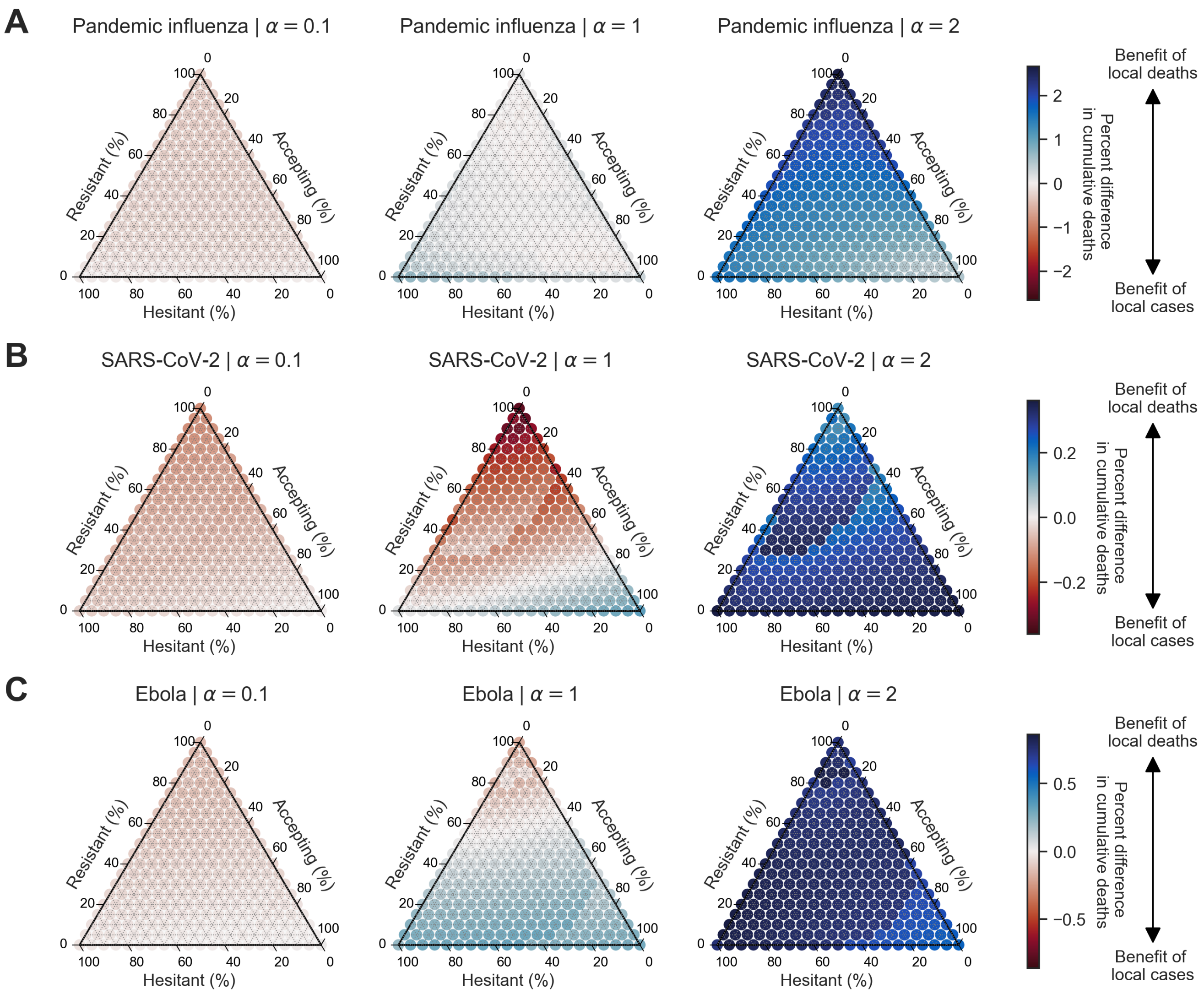

**Figure S32. Heterogeneous scenario 1. Percent difference in cumulative deaths between local cases and local deaths across pathogen systems and information sensitivity with a vaccine efficacy of 25%.** For each panel, the ternary plot axes show the percentage of the population assigned to each of three behavioural groups: vaccine-resistant ($\rho = 0$), vaccine-hesitant ($\rho = 1$) and vaccine-accepting ($\rho = 2$). Each row shows percent difference in cumulative deaths between outbreak information scenarios based on local cases and local deaths for a given pathogen: **(A)** pandemic influenza, **(B)** SARS-CoV-2, **(C)** Ebola. Each column corresponds to a different level of information sensitivity: $\alpha = 0.1$ (first column), $\alpha = 1$ (second column), $\alpha = 2$ (third column). Darker colour hues indicate more severe outcomes in terms of percent difference in cumulative deaths.

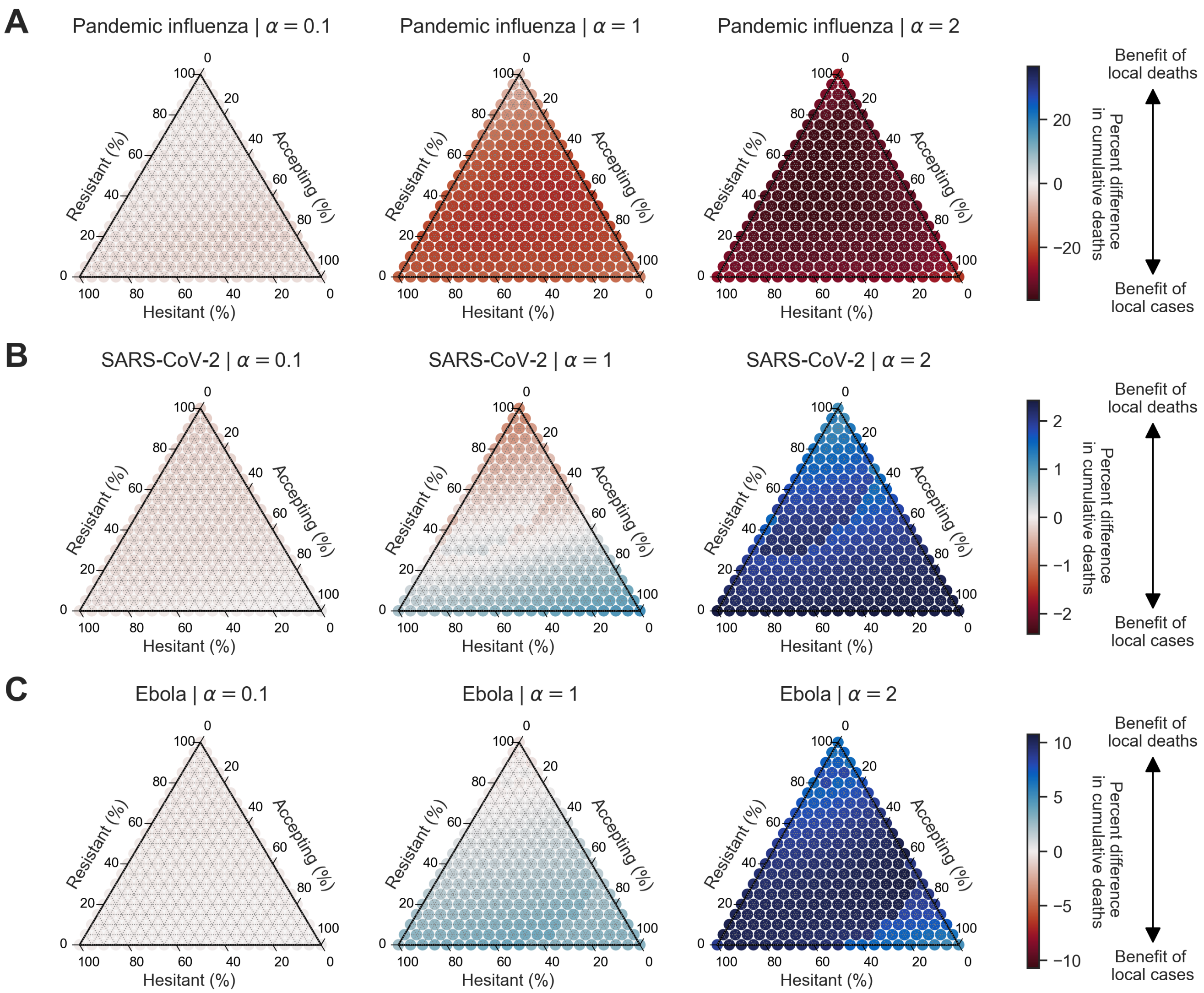

**Figure S33. Heterogeneous scenario 1. Percent difference in cumulative deaths between local cases and local deaths across pathogen systems and information sensitivity with a vaccine efficacy of 50%.** For each panel, the ternary plot axes show the percentage of the population assigned to each of three behavioural groups: vaccine-resistant ($\rho = 0$), vaccine-hesitant ($\rho = 1$) and vaccine-accepting ($\rho = 2$). Each row shows percent difference in cumulative deaths between outbreak information scenarios based on local cases and local deaths for a given pathogen: **(A)** pandemic influenza, **(B)** SARS-CoV-2, **(C)** Ebola. Each column corresponds to a different level of information sensitivity: $\alpha = 0.1$ (first column), $\alpha = 1$ (second column), $\alpha = 2$ (third column). Darker colour hues indicate more severe outcomes in terms of percent difference in cumulative deaths.

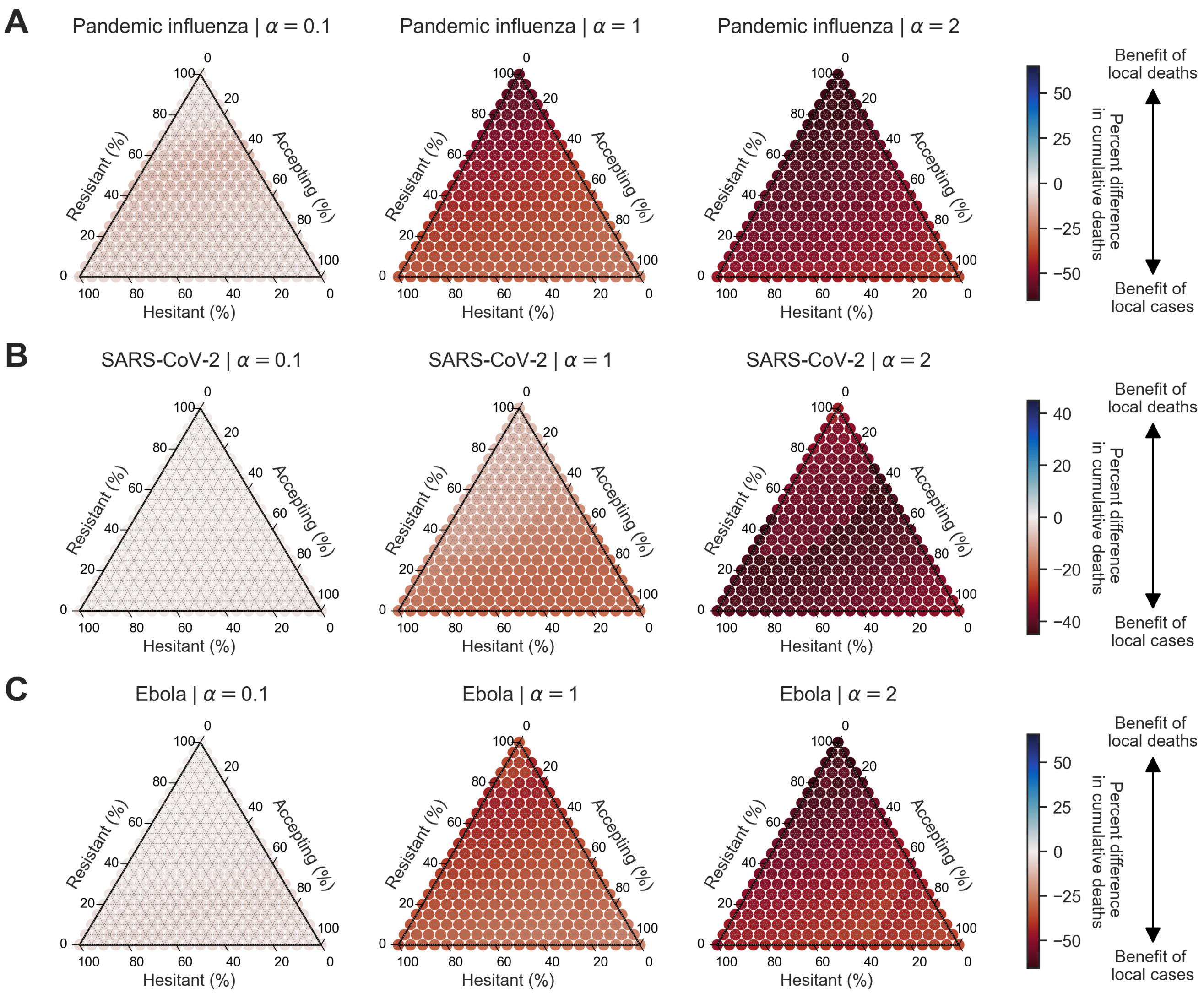

**Figure S34. Heterogeneous scenario 1. Percent difference in cumulative deaths between local cases and local deaths across pathogen systems and information sensitivity with a vaccine efficacy of 90%.** For each panel, the ternary plot axes show the percentage of the population assigned to each of three behavioural groups: vaccine-resistant ($\rho = 0$), vaccine-hesitant ($\rho = 1$) and vaccine-accepting ($\rho = 2$). Each row shows percent difference in cumulative deaths between outbreak information scenarios based on local cases and local deaths for a given pathogen: **(A)** pandemic influenza, **(B)** SARS-CoV-2, **(C)** Ebola. Each column corresponds to a different level of information sensitivity: $\alpha = 0.1$ (first column), $\alpha = 1$ (second column), $\alpha = 2$ (third column). Darker colour hues indicate more severe outcomes in terms of percent difference in cumulative deaths.

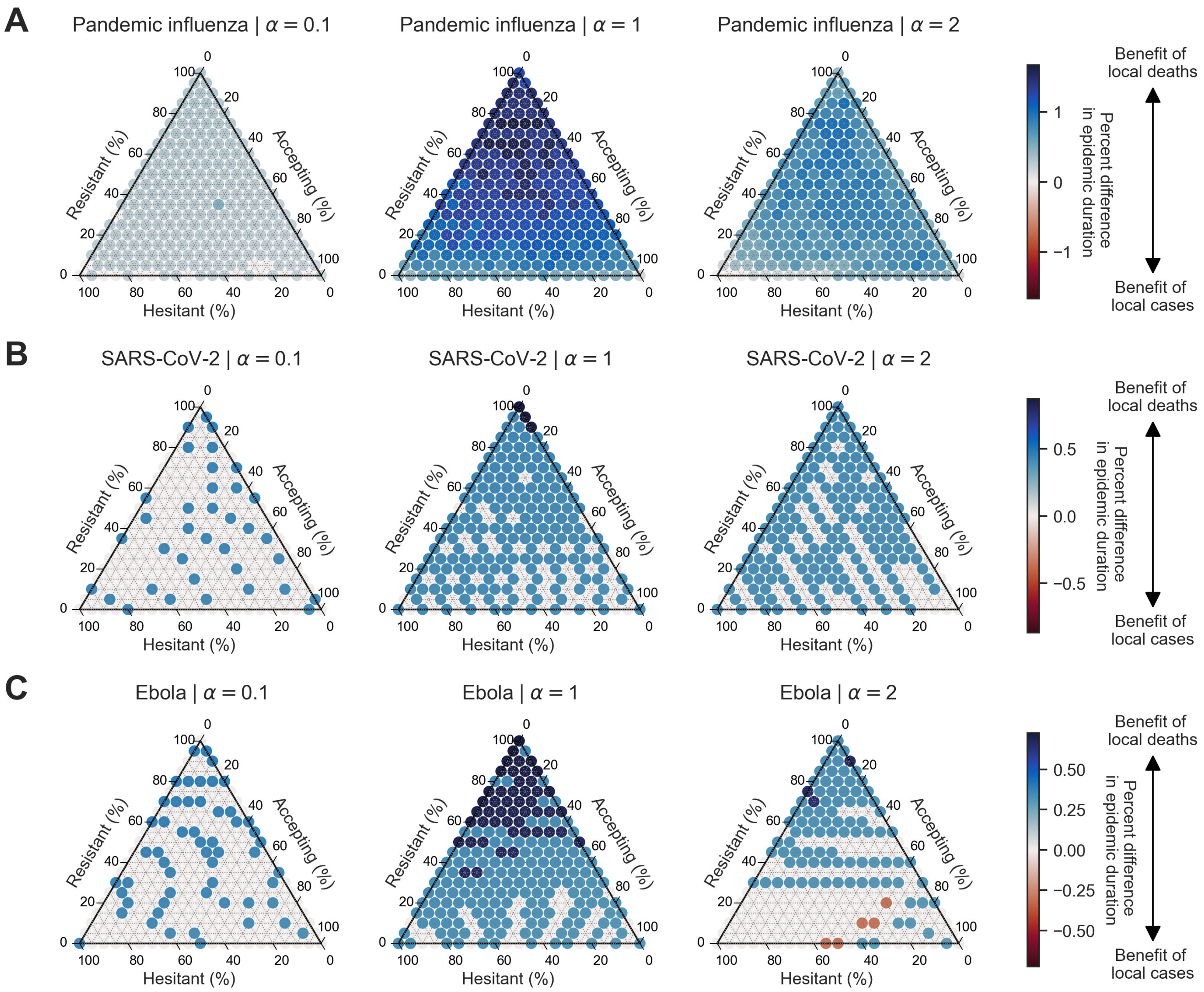

**Figure S35. Heterogeneous scenario 1. Percent difference in epidemic duration in days between local cases and local deaths across pathogen systems and information sensitivity with a vaccine efficacy of 25%.** For each panel, the ternary plot axes show the percentage of the population assigned to each of three behavioural groups: vaccine-resistant ($\rho = 0$), vaccine-hesitant ($\rho = 1$) and vaccine-accepting ($\rho = 2$). Each row shows percent difference in epidemic duration between outbreak information scenarios based on local cases and local deaths for a given pathogen: **(A)** pandemic influenza, **(B)** SARS-CoV-2, **(C)** Ebola. Each column corresponds to a different level of information sensitivity: $\alpha = 0.1$ (first column), $\alpha = 1$ (second column), $\alpha = 2$ (third column). Darker colour hues indicate more severe outcomes in terms of percent difference in epidemic duration.

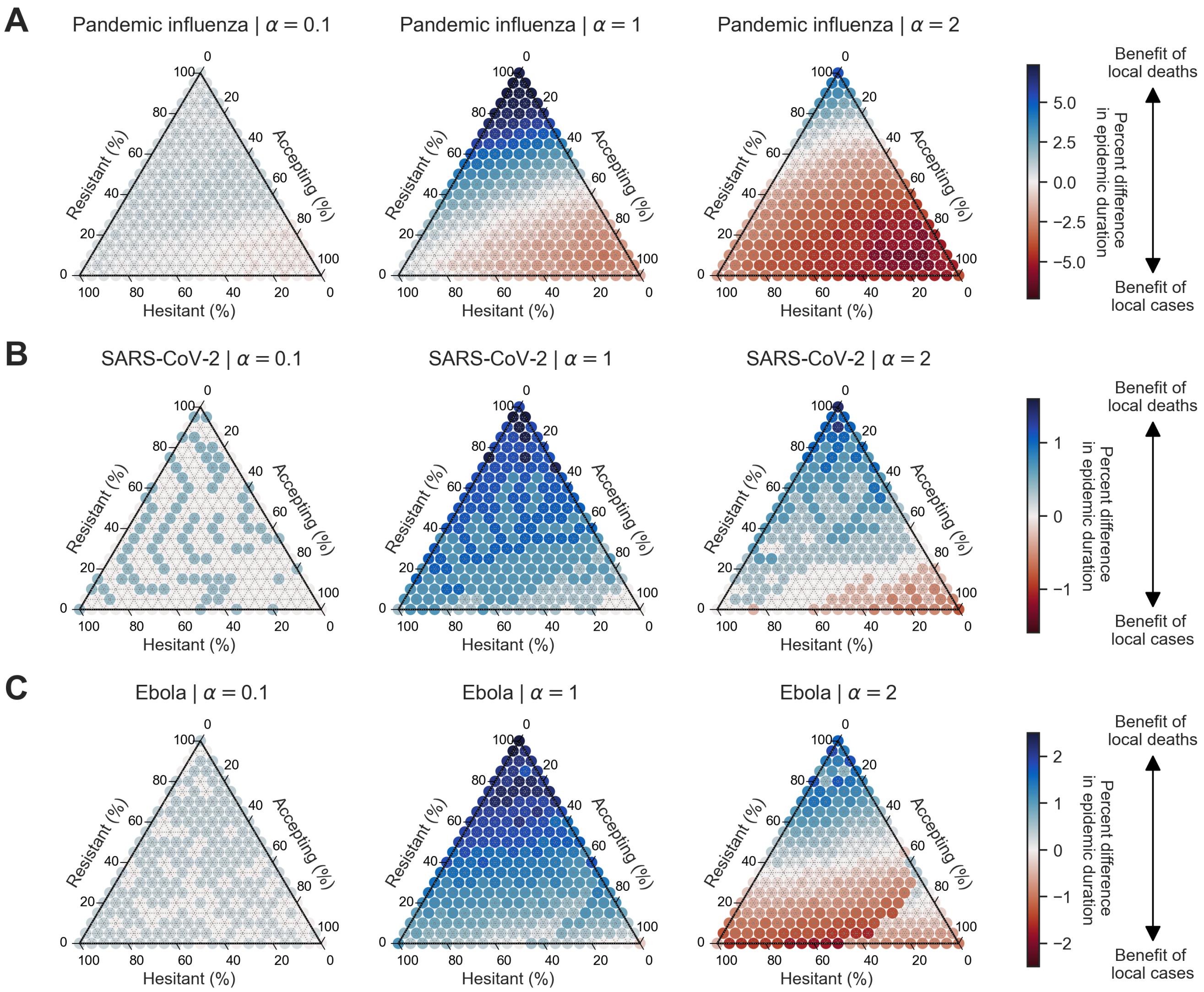

**Figure S36. Heterogeneous scenario 1. Percent difference in epidemic duration in days between local cases and local deaths across pathogen systems and information sensitivity with a vaccine efficacy of 50%.** For each panel, the ternary plot axes show the percentage of the population assigned to each of three behavioural groups: vaccine-resistant ($\rho = 0$), vaccine-hesitant ($\rho = 1$) and vaccine-accepting ($\rho = 2$). Each row shows percent difference in epidemic duration between outbreak information scenarios based on local cases and local deaths for a given pathogen: **(A)** pandemic influenza, **(B)** SARS-CoV-2, **(C)** Ebola. Each column corresponds to a different level of information sensitivity: $\alpha = 0.1$ (first column), $\alpha = 1$ (second column), $\alpha = 2$ (third column). Darker colour hues indicate more severe outcomes in terms of percent difference in epidemic duration.

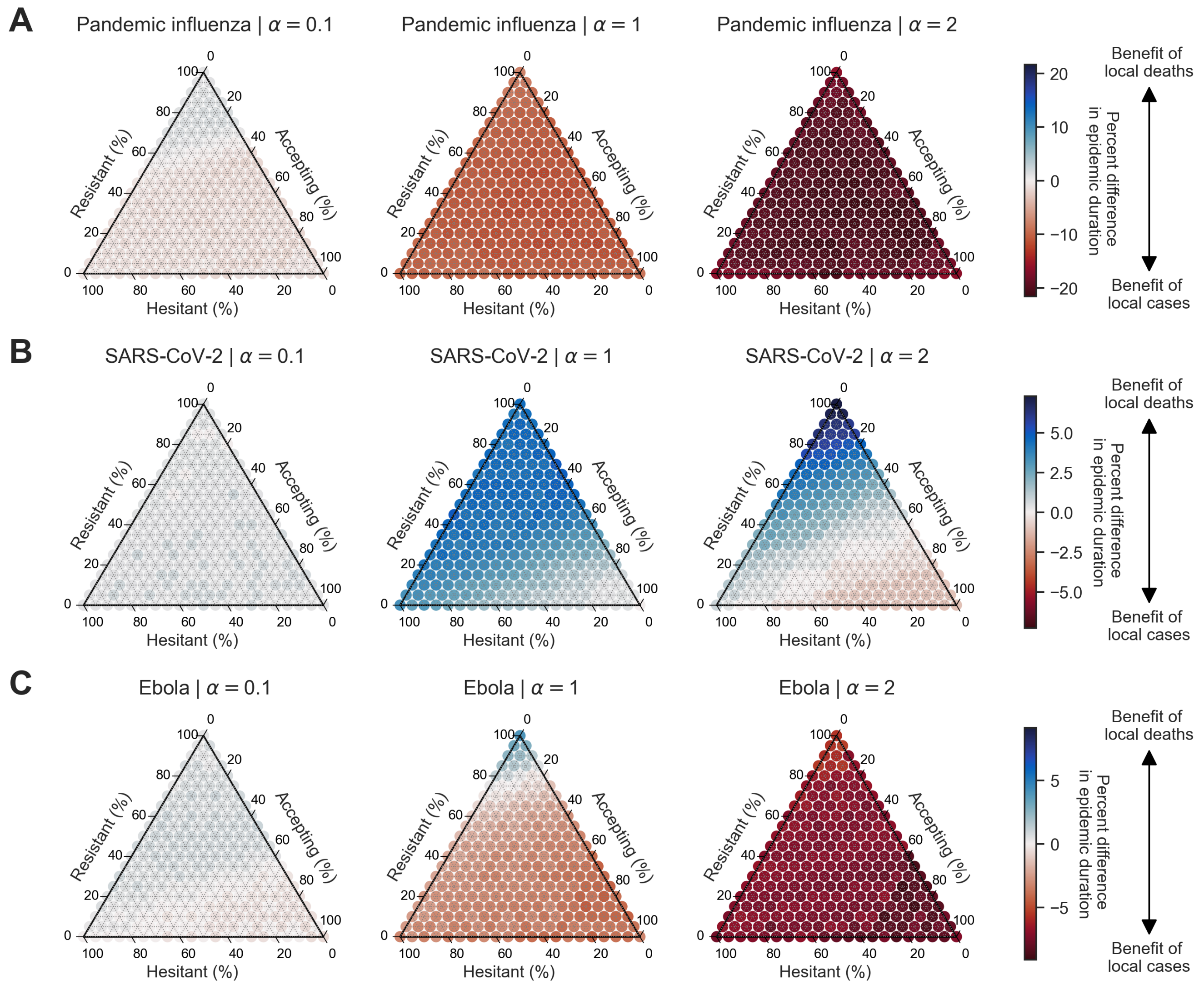

**Figure S37. Heterogeneous scenario 1. Percent difference in epidemic duration in days between local cases and local deaths across pathogen systems and information sensitivity with a vaccine efficacy of 90%.** For each panel, the ternary plot axes show the percentage of the population assigned to each of three behavioural groups: vaccine-resistant ($\rho = 0$), vaccine-hesitant ($\rho = 1$) and vaccine-accepting ($\rho = 2$). Each row shows percent difference in epidemic duration between outbreak information scenarios based on local cases and local deaths for a given pathogen: **(A)** pandemic influenza, **(B)** SARS-CoV-2, **(C)** Ebola. Each column corresponds to a different level of information sensitivity: $\alpha = 0.1$ (first column), $\alpha = 1$ (second column), $\alpha = 2$ (third column). Darker colour hues indicate more severe outcomes in terms of percent difference in epidemic duration.

##### S2.2 Heterogeneous scenario 2

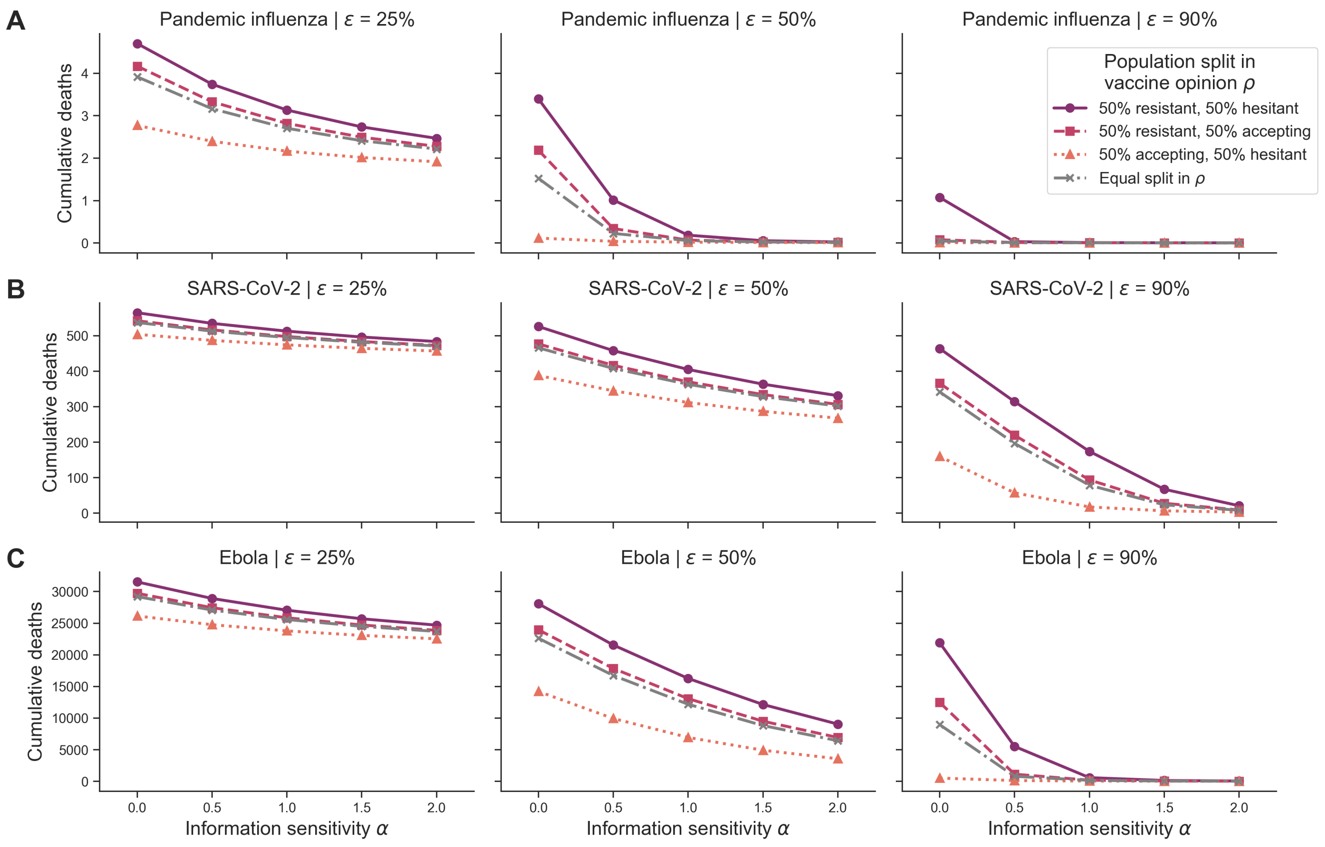

**Figure S38. Heterogeneous scenario 2. Cumulative deaths across pathogen systems, vaccine efficacy for heterogeneous behavioural configurations with local cases as outbreak information.** Each row shows cumulative deaths for a given pathogen system: **(A)** pandemic influenza, **(B)** SARS-CoV-2 and **(C)** Ebola. Each column shows a different vaccine efficacy ($\varepsilon$): 25% (first column), 50% (second column) and 90% (third column). Different line types, colours and markers indicate different mixed behavioural configurations: 50% resistant ($\rho = 0$), 50% hesitant ($\rho = 1$) (solid purple line with circle markers), 50% resistant ($\rho = 0$), 50% accepting ($\rho = 2$) (dashed pink line with square markers), 50% accepting ($\rho = 2$), 50% hesitant ($\rho = 1$) (orange dotted line with triangle markers) and equal split in vaccine opinion ($\rho \in 0, 1, 2$, grey dashed-dotted line with x markers). The memory window ($\mu$) was fixed at the full history.

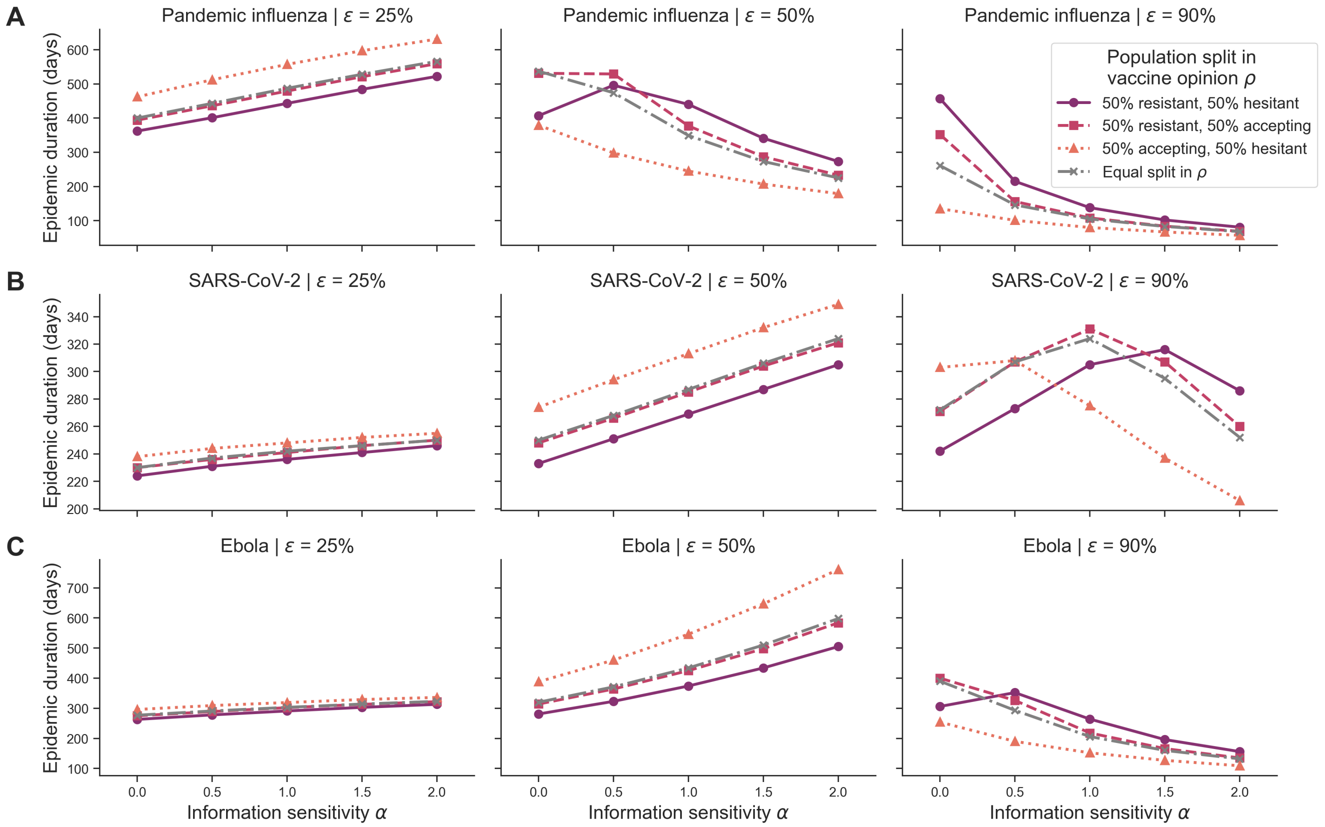

**Figure S39. Heterogeneous scenario 2. Epidemic duration in days across pathogen systems, vaccine efficacy for heterogeneous behavioural configurations with local cases as outbreak information.** Each row shows epidemic duration for a given pathogen system: **(A)** pandemic influenza, **(B)** SARS-CoV-2 and **(C)** Ebola. Each column shows a different vaccine efficacy ($\varepsilon$): 25% (first column), 50% (second column) and 90% (third column). Different line types, colours and markers indicate different mixed behavioural configurations: 50% resistant ($\rho = 0$), 50% hesitant ($\rho = 1$) (solid purple line with circle markers), 50% resistant ($\rho = 0$), 50% accepting ($\rho = 2$) (dashed pink line with square markers), 50% accepting ($\rho = 2$), 50% hesitant ($\rho = 1$) (orange dotted line with triangle markers) and equal split in vaccine opinion ($\rho \in 0, 1, 2$, grey dashed-dotted line with x markers). The memory window ($\mu$) was fixed at the full history.

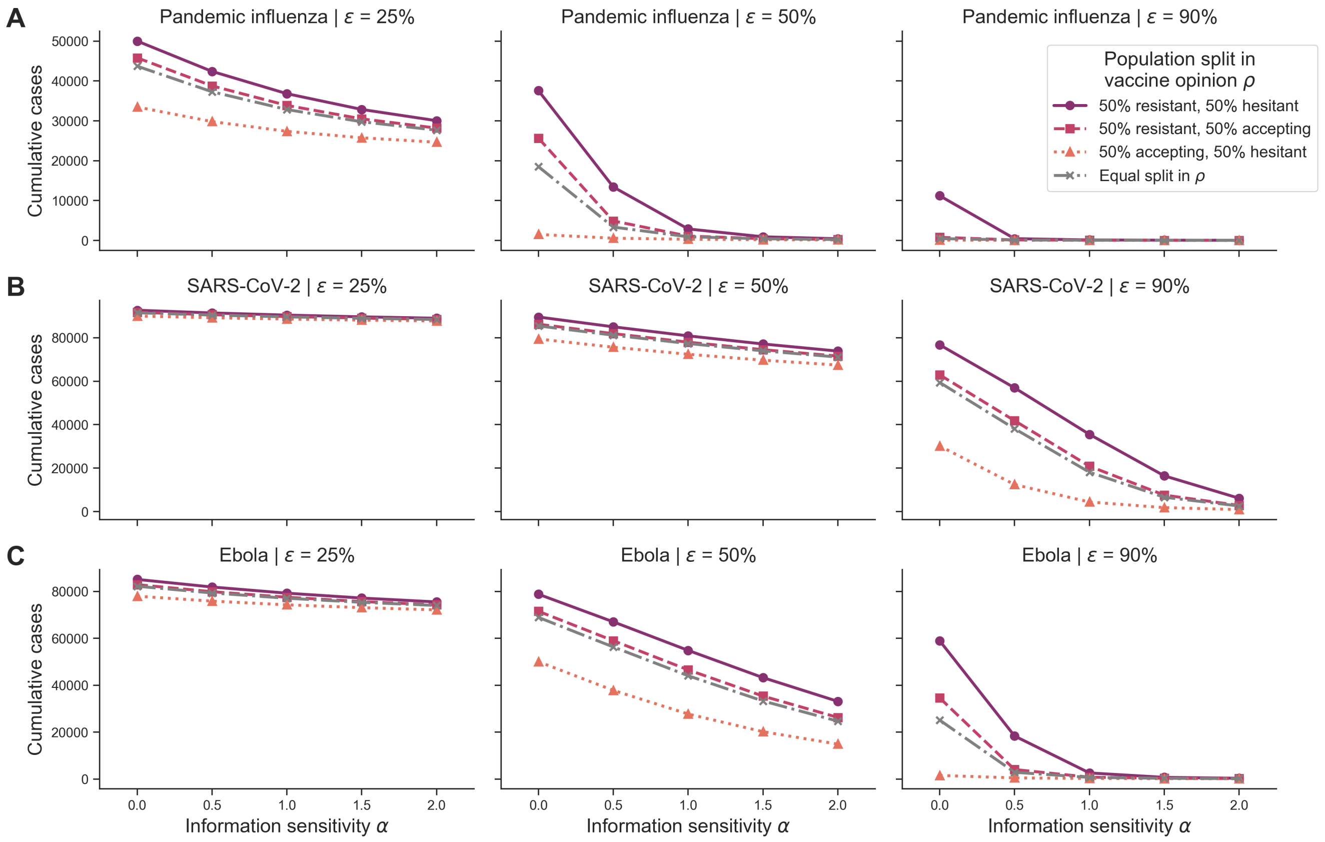

**Figure S40. Heterogeneous scenario 2. Cumulative cases across pathogen systems, vaccine efficacy for heterogeneous behavioural configurations with local deaths as outbreak information.** Each row shows cumulative cases for a given pathogen system: **(A)** pandemic influenza, **(B)** SARS-CoV-2 and **(C)** Ebola. Each column shows a different vaccine efficacy ($\varepsilon$): 25% (first column), 50% (second column) and 90% (third column). Different line types, colours and markers indicate different mixed behavioural configurations: 50% resistant ($\rho = 0$), 50% hesitant ($\rho = 1$) (solid purple line with circle markers), 50% resistant ($\rho = 0$), 50% accepting ($\rho = 2$) (dashed pink line with square markers), 50% accepting ($\rho = 2$), 50% hesitant ($\rho = 1$) (orange dotted line with triangle markers) and equal split in vaccine opinion ($\rho \in 0, 1, 2$, grey dashed-dotted line with x markers). The memory window ($\mu$) was fixed at the full history.

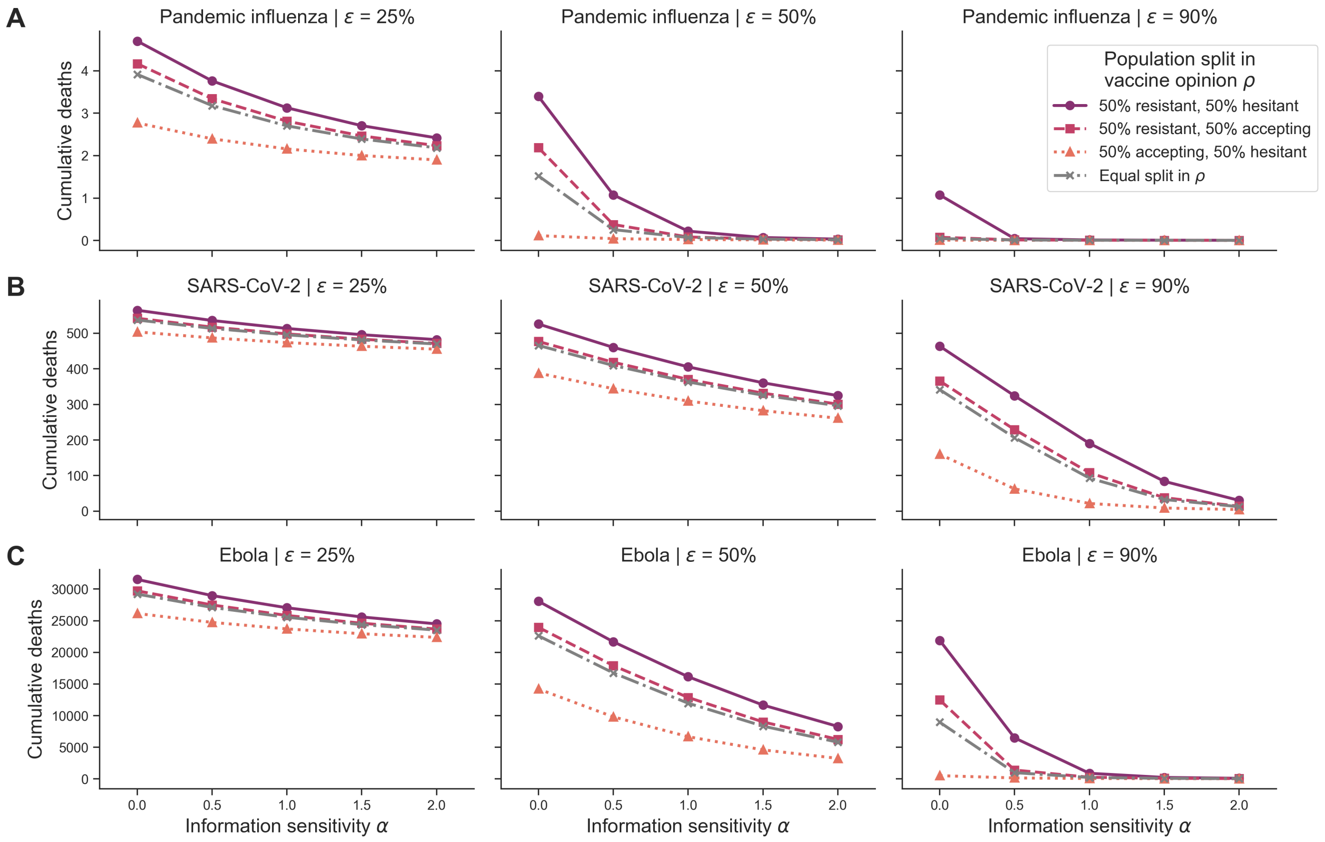

**Figure S41. Heterogeneous scenario 2. Cumulative deaths across pathogen systems, vaccine efficacy for heterogeneous behavioural configurations with local deaths as outbreak information.** Each row shows cumulative deaths for a given pathogen system: **(A)** pandemic influenza, **(B)** SARS-CoV-2 and **(C)** Ebola. Each column shows a different vaccine efficacy ($\varepsilon$): 25% (first column), 50% (second column) and 90% (third column). Different line types, colours and markers indicate different mixed behavioural configurations: 50% resistant ($\rho = 0$), 50% hesitant ($\rho = 1$) (solid purple line with circle markers), 50% resistant ($\rho = 0$), 50% accepting ($\rho = 2$) (dashed pink line with square markers), 50% accepting ($\rho = 2$), 50% hesitant ($\rho = 1$) (orange dotted line with triangle markers) and equal split in vaccine opinion ($\rho \in 0, 1, 2$, grey dashed-dotted line with x markers). The memory window ($\mu$) was fixed at the full history.

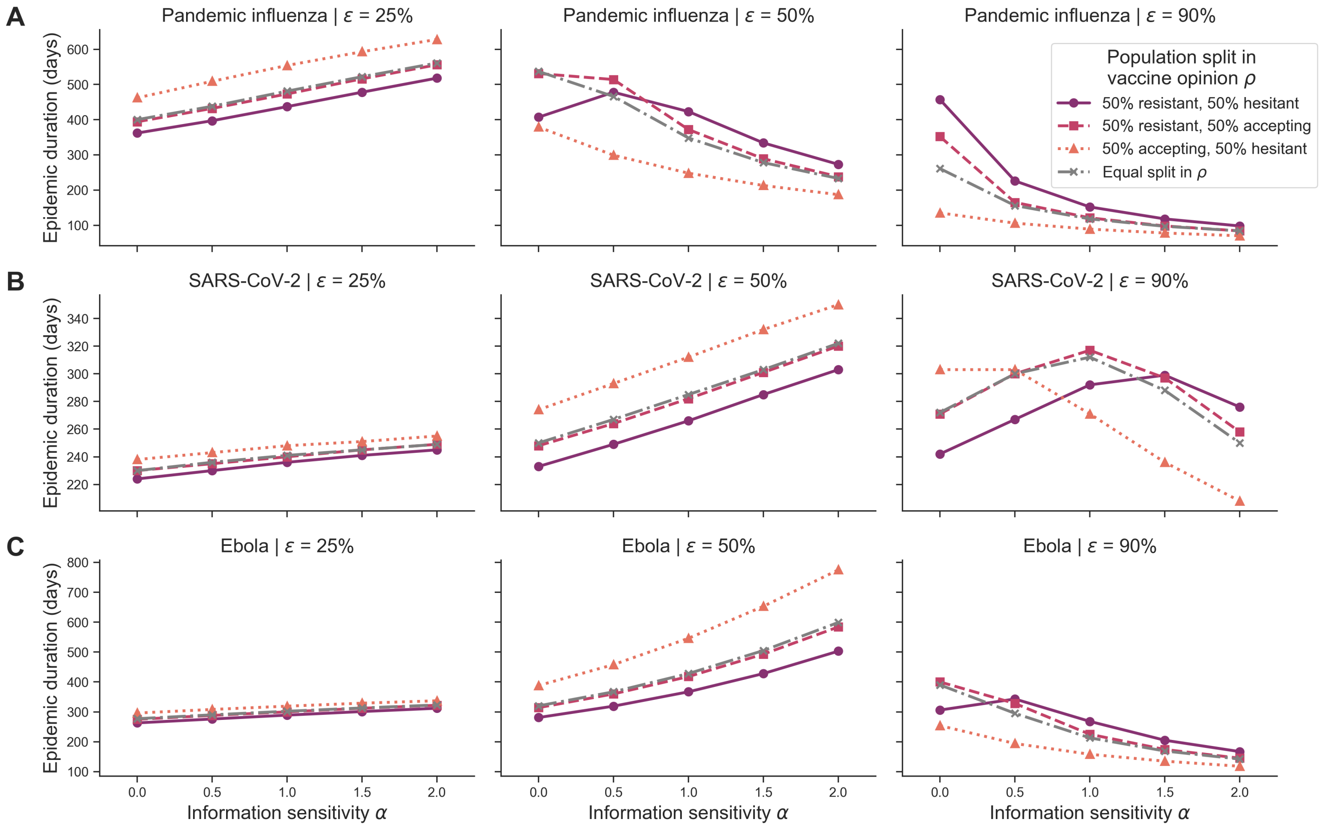

**Figure S42. Heterogeneous scenario 2. Epidemic duration in days across pathogen systems, vaccine efficacy for heterogeneous behavioural configurations with local deaths as outbreak information.** Each row shows epidemic duration for a given pathogen system: **(A)** pandemic influenza, **(B)** SARS-CoV-2 and **(C)** Ebola. Each column shows a different vaccine efficacy ($\varepsilon$): 25% (first column), 50% (second column) and 90% (third column). Different line types, colours and markers indicate different mixed behavioural configurations: 50% resistant ($\rho = 0$), 50% hesitant ($\rho = 1$) (solid purple line with circle markers), 50% resistant ($\rho = 0$), 50% accepting ($\rho = 2$) (dashed pink line with square markers), 50% accepting ($\rho = 2$), 50% hesitant ($\rho = 1$) (orange dotted line with triangle markers) and equal split in vaccine opinion ($\rho \in 0, 1, 2$, grey dashed-dotted line with x markers). The memory window ($\mu$) was fixed at the full history.

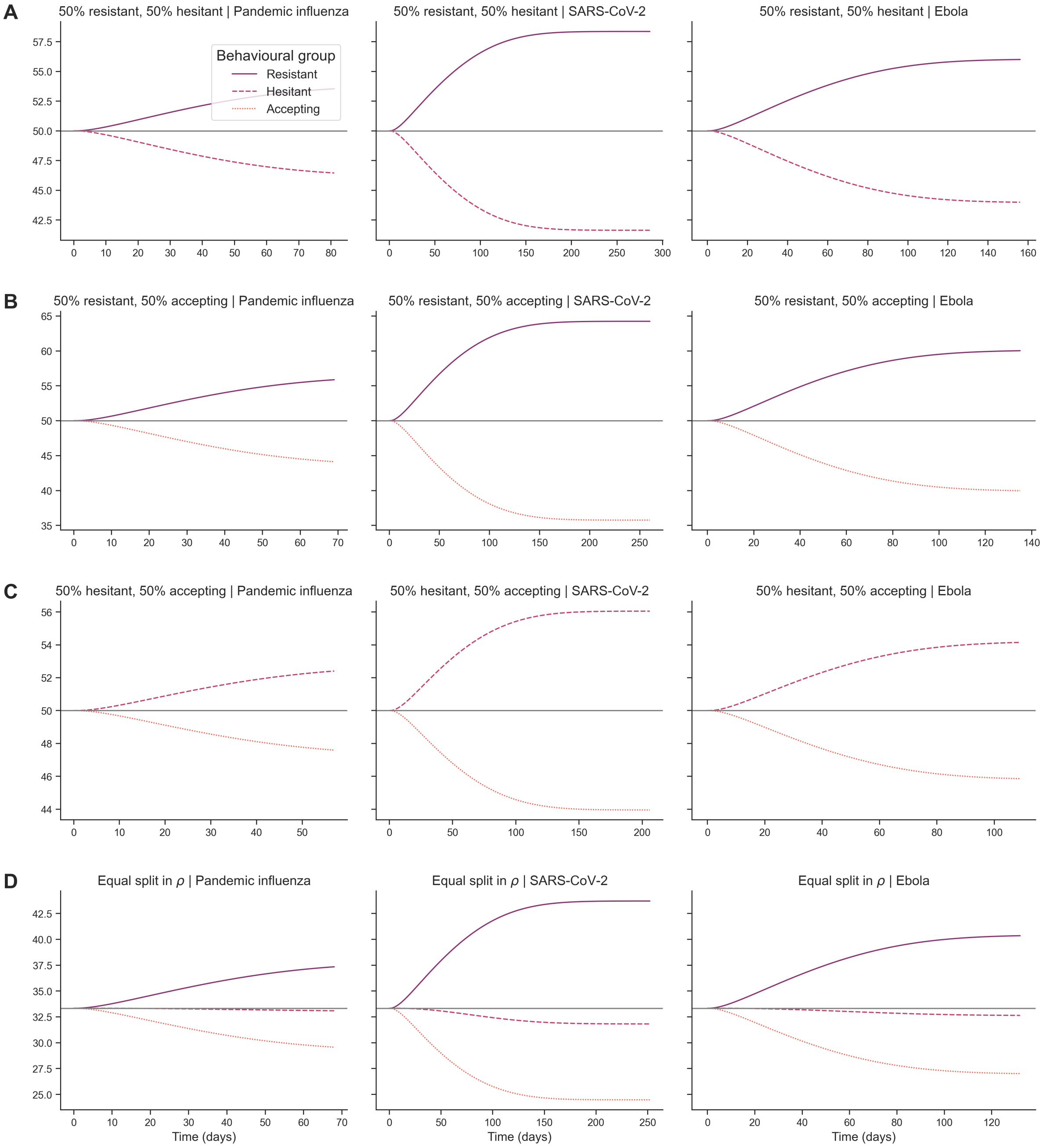

**Figure S43. Heterogeneous scenario 2. Subpopulation-level percent contribution to cumulative deaths over time across pathogen systems for heterogeneous behavioural configurations and outbreak information based on local cases.** Each row shows percent contribution for each subpopulation towards cumulative deaths across time in days for a unique behavioural configuration: **(A)** 50% resistant ($\rho = 0$), 50% accepting ($\rho = 2$); **(B)** 50% resistant ($\rho = 0$), 50% hesitant ($\rho = 1$); **(C)** 50% accepting ($\rho = 2$), 50% hesitant ($\rho = 1$); and **(D)** equal split in vaccine opinion ($\rho \in 0, 1, 2$). Each column represents a different pathogen system: pandemic influenza (first column), SARS-CoV-2 (second column) and Ebola (third column). For each behavioural configuration, lines are shown for each behavioural subpopulation: vaccine-resistant ($\rho = 0$), vaccine-hesitant ($\rho = 1$) and vaccine-accepting ($\rho = 2$). The grey colours represent subpopulations which are not represented in the given configuration. Information sensitivity ($\alpha$) was fixed at 2, memory window ($\mu$) was fixed at the full history and vaccine efficacy ($\varepsilon$) was fixed at 90%.
